# Self-reported knowledge attitude and practice of healthcare professionals in the management of infection and antimicrobial stewardship: a systematic review

**DOI:** 10.1101/2025.04.28.25324348

**Authors:** Iman Ghosh, Adebisi Adedunmola, Erkan Alkan, Victoria Adetunji, Charlotte Web, Philip Anyanwu, Samantha Johnson, Ellie Gilham, Diane Ashiru-Oredope, Abimbola Ayorinde

**Affiliations:** University of Warwick; Centre of Evidence and Implementation Science, University of Birmingham.; UK Health Security Agency; School of Pharmacy, University College London.

## Abstract

**Objectives:** This review aims to synthesise studies on health and social care professionals (HCPs) knowledge, attitudes, and practices (KAP) regarding infection management, infection prevention and control, antimicrobial use, stewardship, and resistance to inform future research and policy.

**Method:** In January 2024, we conducted a comprehensive search in Medline, Embase, Web of Science, and CINAHL to identify studies on health and social care professionals’ KAP regarding infection management, prevention and control, antimicrobial use, stewardship, and resistance. After deduplication, the initial screening was conducted in Rayyan, with 10% checked for accuracy. Two reviewers independently assessed full texts. Data extraction was performed by one and verified by another reviewer. Quality assessment was completed by one reviewer, with 20% checked for accuracy. We included relevant studies published from 2016 onwards focusing on those conducted in the UK and in countries with comparable settings. Finally, a narrative synthesis was carried out due to significant differences between studies.

**Results:** Out of 10,990 unique records identified, 113 studies with diverse participants and settings were included. The findings showed substantial variation in KAP measures, complicating direct comparisons between studies. Some studies assessed objective knowledge(N=40) while most measured perceived knowledge(N=41), revelling discrepancies between the two. Attitude assessments revealed widespread consensus on the harms of inappropriate antimicrobial use, though willingness to participate in antimicrobial stewardship (AMS) activities varied by profession. Practice behaviour assessment indicated varying hand-hygiene compliance and AMS implementation, along with significant concerns about inappropriate antibiotic prescribing.

**Conclusion:** The review highlights significant gaps in healthcare professionals’ KAP regarding infection prevention and antimicrobial stewardship, with variations across professions. This underscores the need for targeted interventions. Additionally, standardised KAP assessment measures are essential to enhance comparability across different contexts. These findings provide a foundation for future research and policy initiatives aimed at combating AMR.

## 1. Introduction

Antimicrobial Resistance (AMR) happens when bacteria, viruses, fungi, and parasites become resistant to antimicrobial drugs, making these treatments ineffective.[1] AMR is one of the top ten threats to global health.[1] Since the introduction of the first antimicrobial agent, Salvarsan, in 1910, these drugs have been crucial in treating various infections.[2] However, factors like the misuse or overuse of antibiotics, and patients not completing their prescribed courses have contributed to the development of resistance in many pathogens.[3] This resistance leads to infections becoming harder to treat, which has significant clinical and economic impact.[4, 5] In 2019, the World Health Organization (WHO) estimated that AMR directly caused 1.27 million deaths globally. Economically, the Organisation for Economic Co-operation and Development (OECD) estimated in 2018 that AMR could cost up to 3.5 billion US dollars annually across Europe, North America, and Australia.[6]

Alongside antibiotic prudency, infection prevention, control, and management (IPCM) measures are also crucial in managing AMR. Research has shown that improving IPCM practices can help reduce AMR rates, while inappropriate IPCM measures and excessive antimicrobial prescribing can increase resistance.[6, 7] Efforts to combat AMR have been initiated both in the UK and globally. On an international scale, the ‘Global Action Plan on AMR’ was adopted during the World Health Assembly in 2015 which had countries commit to 5 AMR-related goals.[8] Furthermore, the WHO has established a ‘quadripartite joint secretariat’ which has facilitated the creation of a ‘Global Leaders Group on AMR’. Since 2016 the United Nations General Assembly have had a key role in organising international High-Level Meetings to plan action against AMR, the latest of which occurred in September 2024.[9] In the UK, the government has set a goal to contain and control AMR by 2040 and has implemented rolling 5-year action plans to combat AMR, the latest of which began in 2024 and will run until 2029.[10]

Antimicrobial Stewardship (AMS) and IPCM programmes aim to address AMR by increasing knowledge and improving practices related to this topic (e.g. stopping over-prescription of antibiotics, promoting correct hand hygiene techniques).[11] Current and future health and social care professionals (including workers, students and leaders) are a key target of AMS and IPCM programmes, as many of them are directly involved in the practices of infection management and antibiotics prescribing. To support the UK National Action Plan (NAP) on AMR, which aims to equip the health, animal, and agriculture sectors with the necessary skills and governance to tackle AMR by 2029, a key target is to increase public and healthcare professionals’ knowledge of AMR by 10% based on 2018–2019 baselines.[12] Whilst many primary studies have investigated the knowledge, attitudes and practices (KAP) of HCPs towards AMR, their findings tend to vary.[13–15]. Additionally, existing review often fail to encompass the full range of HCPs. They typically focus on healthcare professionals (such as doctors, nurses and pharmacists) but overlook other important groups, such as students enrolled in health and social care programmes. This underscores the urgent need for a comprehensive review to consolidate current studies on KAP among health and social care professionals, in order to identify gaps and inform future interventions to combat AMR.

### 1.1. Aims and objectives

We systematically reviewed the literature to assess knowledge, attitudes, and practices among health and social care professionals (including students, workers, and leaders) concerning the management of infections, infection prevention and control, antimicrobial use, stewardship, and resistance. A secondary objective of this study was to collate survey instruments and describe the types of questions used in KAP surveys among health and social care professionals.

## 2. Materials and Method

### 2.1. Information source

We conducted a comprehensive search of four databases: MEDLINE All (via Ovid), EMBASE (via Ovid), Web of Science, and the Cumulative Index to Nursing and Allied Health Literature (CINAHL) to identify relevant literature. An information specialist (SJ) developed the search strategy and ran the searches in January 2024. The search strategy included terms associated with health and social care students, workers, and professionals, as well as keywords related to knowledge, attitudes, or practices in the context of infection prevention and control, antimicrobial use, stewardship, and resistance. We also incorporated search terms such as surveys, questionnaires, tools, and instruments to narrow down the results. Additionally, we restricted the search to English-language publications only, since the year 2016. Given the overwhelming number of relevant records identified in the initial search and the time constraints, we did not perform any additional searches. We limited our focus to Health and social care in the UK and countries with similar settings (including European countries, United States of America, Canada, Australia and New Zealand)

The search strategy and terms used are presented in the Appendix 1.

### 2.2. Study selection

We imported the records into an Endnote library, and deduplication was performed. The unique records were then exported to an online platform, Rayyan (https://www.rayyan.ai/), to facilitate screening. A predefined list of inclusion and exclusion criteria (Table 1) was used to perform title and abstract screening. Titles and abstracts of all citations were screened by one reviewer (IG, EA, AA, VA, AAy, or PA), and 10% were double screened by a second reviewer. Any disagreements were resolved through discussion or with the involvement of a third reviewer.

**Table 1:** Study eligibility criteria.

| Item | Inclusion criteria | Exclusion criteria |
| --- | --- | --- |
| Participants/<br>population | Any health and social care professional or student expected to have prescribing, dispensing or administration responsibilities. | A mixed population of Health and social care professionals, patients and the general public, where data specific to health and social care professionals' not reported separately |
| Context | Health and social care in the UK and countries with similar settings (including European countries, United States of America, Canada, Australia and New Zealand) | Studies from low- and middle-income countries and other settings where the healthcare systems are not similar to the UK settings. |
| Outcome | Knowledge related to infection management, infection prevention control, antimicrobial use, stewardship, and resistance (such as, proportion of respondents who correctly answer key knowledge questions, as reported in the primary study).<br>Attitude towards management of infections, infection prevention control, antimicrobial use, stewardship, and resistance (such as, the proportion of agreement/disagreement regarding | Any other outcomes |
|  | personal role in addressing antibiotic resistance).<br>Practice regarding management of infections, infection prevention control, antimicrobial use, stewardship, and resistance (such as, the number of antibiotics prescribed) |  |
| Study type | Quantitative, cross-sectional observation study | We will exclude non-quantitative studies and articles that do not stem from original research, including secondary data analysis, topical reviews, essays, expert opinions, comments, letters, etc. Systematic reviews will be excluded.<br>Studies published before the year 2016 and those not published in the English language. |

Studies that qualified for full-text screening were then transferred to an Excel spreadsheet and double-screened by IG, EA, and VA. Any conflicts in the decision were resolved through discussion between the reviewers and with the involvement of a third reviewer (AAy or PA).

### 2.3. Data extraction

We designed and piloted the data extraction form prepared in Excel to extract information on the first author, study location, study settings, population characteristics, information on the survey instrument, and the outcomes of interest: knowledge, attitude, and practice. Data extraction was performed by an independent reviewer (AA, or VA) and checked by another reviewer (CW, AA or EA).

### 2.4. Quality assessment

The quality of the included studies was assessed using the risk-of-bias assessment tool developed by Hoy et al.[16] This tool assesses 10 criteria, covering external validity (items one to four) and internal validity (items five to ten). Each criterion was assigned a score of “0” (indicating the absence of bias) or “1” (indicating the presence of bias). Studies were categorised into low (0–3), moderate (4–6), and high (7–10) risk of bias based on the cumulative scores. We modified the definition for three criteria in the original 10-item scale according to our study requirements. We defined ‘case definition’ as ‘a definition for knowledge, attitude and practice’, we measured ‘shortest prevalence period’ as ‘recall period of four weeks and measured appropriate numerator and denominator’ when the study has reported ‘actual numbers, not the proportions’ A single reviewer (VA or AA) evaluated the external and internal validity of the included studies, using the 0-1 scoring system and made an overall judgment using the cumulative score for each study An independent second reviewer (IG) randomly cross-verified 20% of the scoring for accuracy and completeness. Any disagreements were resolved through discussion.

### 2.5. Data synthesis

The included studies examined various healthcare professionals using diverse questionnaires and assessed different aspects of infection management, prevention, control, antimicrobial use, stewardship, and resistance. Due to variations in study focus and outcome reporting (e.g., percentages vs. mean scores), conducting a meta-analysis was impractical. Consequently, we employed a narrative synthesis approach. We first categorised the included studies into four groups based on their aims and objectives, such as infection prevention and control measures (IPCM), antimicrobial stewardship (AMS), antimicrobial resistance (AMR), and antimicrobial use. We included studies focusing on hand hygiene (HH), infectious disease prevention strategies, and vaccination within the broader scope of IPCM. Following this, we analysed the reported outcome into three categories— knowledge, attitude and practice. It is important to note that we grouped objective knowledge (measured by yes/no or true/false questions) and perceived knowledge (measured by agree/disagree statements or Likert scales) together under knowledge. Similarly, we grouped all reported practices, whether measured by the number of antibiotics prescribed or using a Likert scale, under practice. We grouped awareness, perception, and capability as knowledge. Attitude included motivation, belief, opportunity, and confidence. Practice encompassed behaviour, use, and compliance.

The protocol for this study was registered online with the PROSPERO database: CRD42024510775.[17]

## 3. Results

### 3.1. Study selection

Figure 1 provides the PRISMA flow diagram for study selection and the reason for exclusion. The searches identified 13,410 records. After removing 2420 duplicate records, 10,990 unique records were screened for their title and abstracts. We identified 295 studies eligible for full-text assessment and 113 of which met the inclusion criteria.

**Figure 1:**
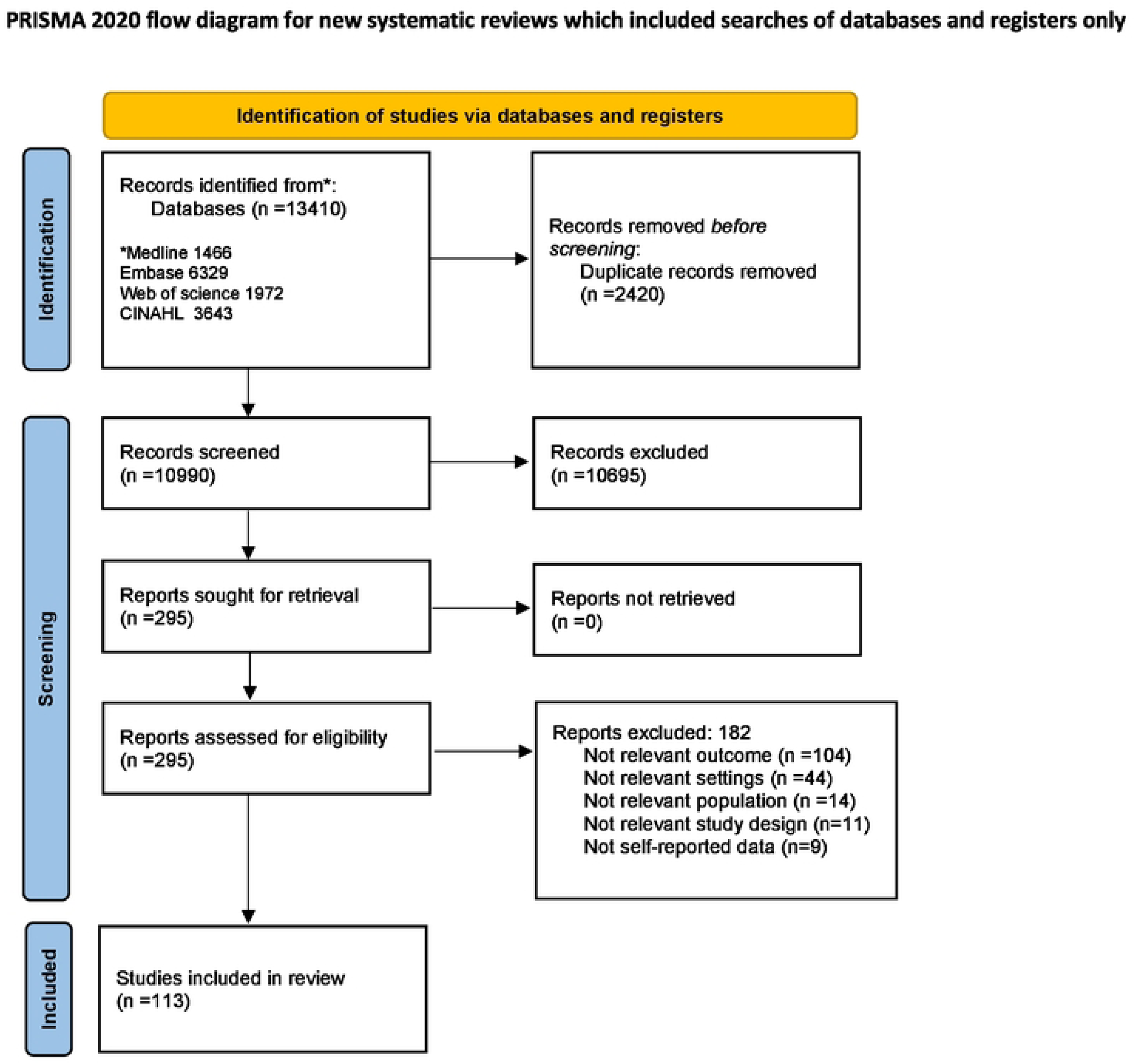
PRISMA flow diagram.

### 3.2. Study characteristics

The studies included in this systematic review are summarised in Table 2. Of the included 113 studies, twenty-seven (n=27) were conducted in the USA,[18–44] eleven each from Australia[45–54] and Italy,[55–65] nine from the United Kingdom,[66–74] seven each from Germany[75–81] and France,[82–88] six each from Poland [89–94]and Canada,[95–100] five from Greece,[101–105] four from Portugal,[106–109] two each from Switzerland,[110, 111] Spain,[112, 113] and New Zealand,[114, 115] and one each from Sweden,[116]

**Table 2:**
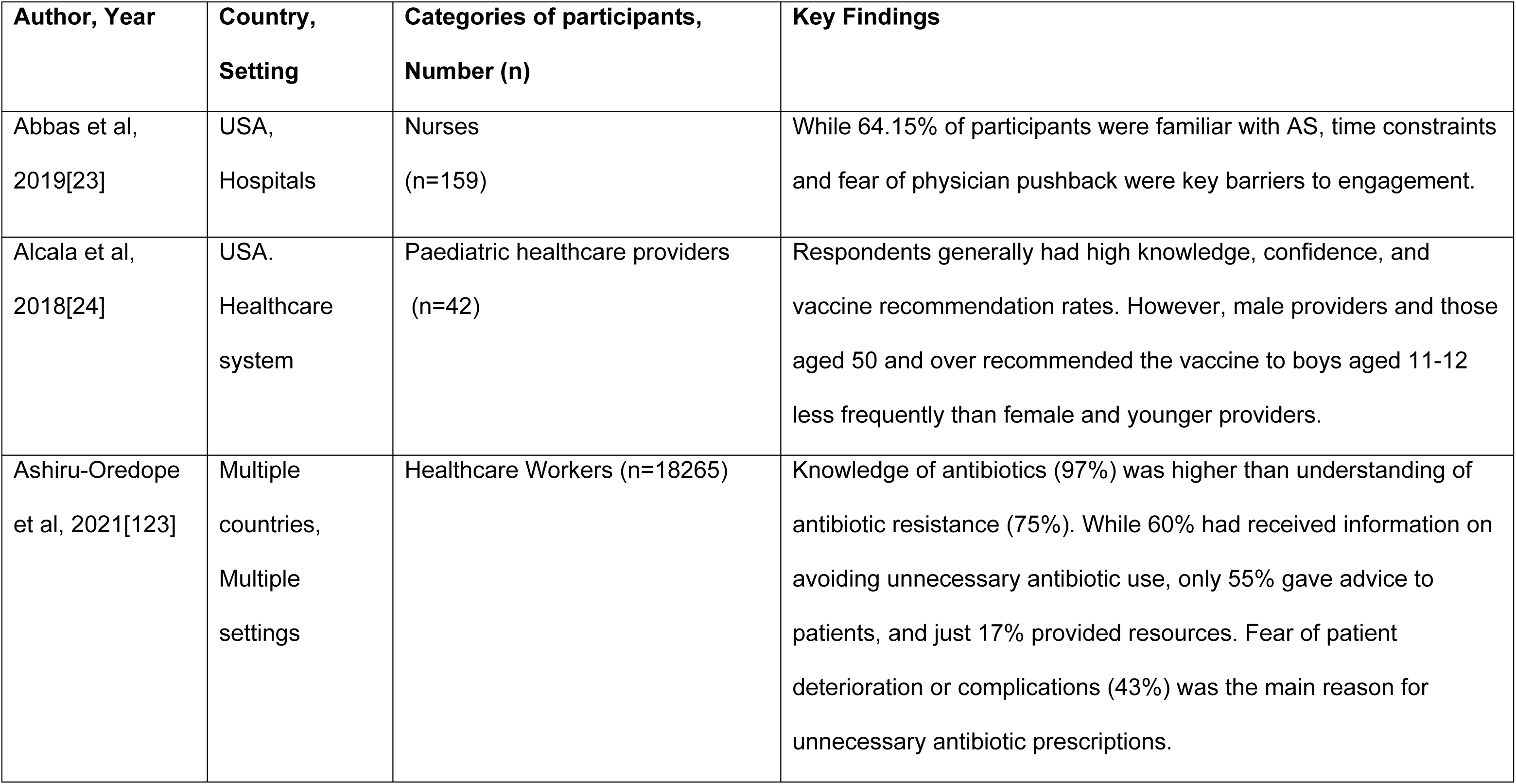
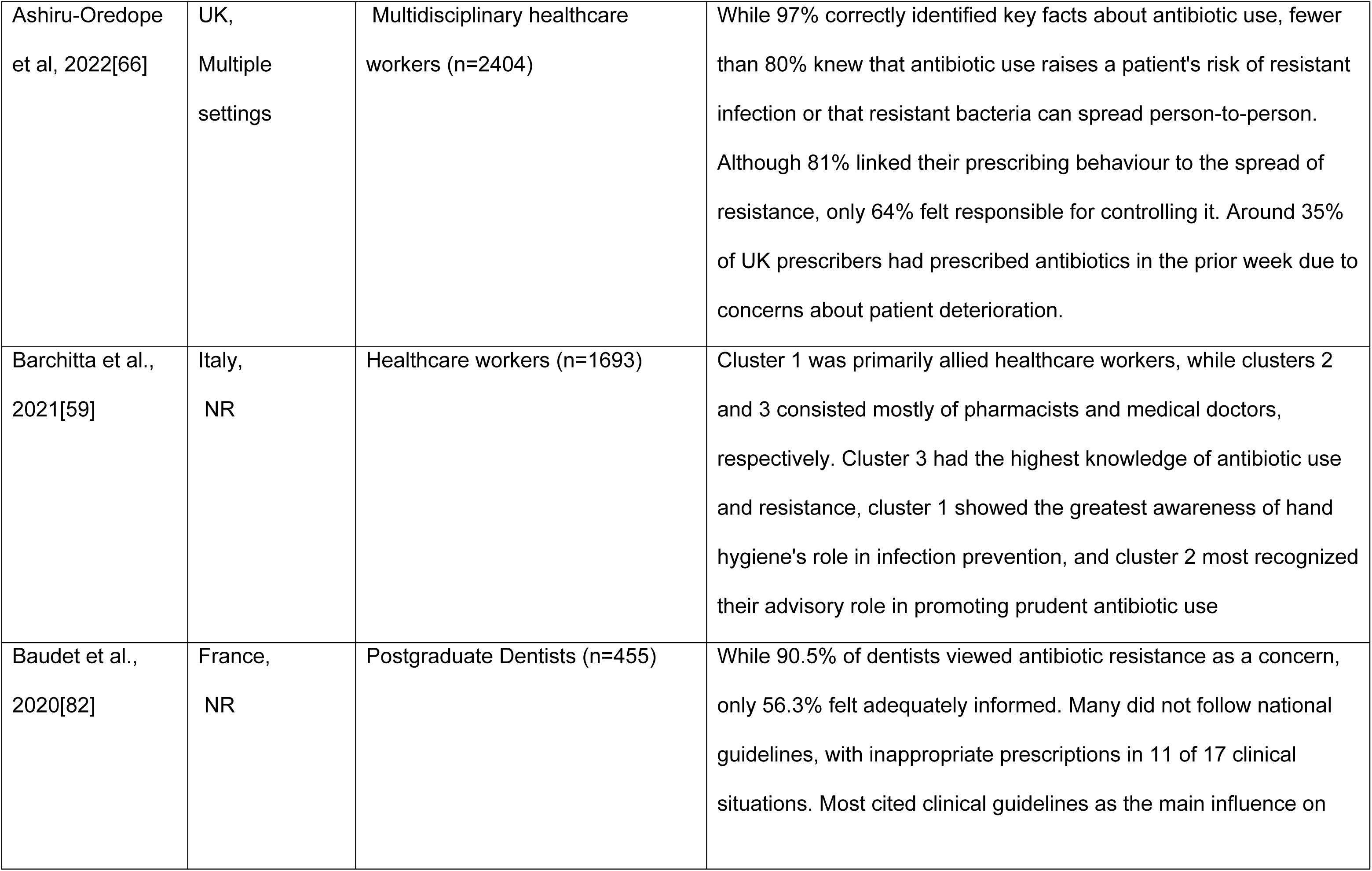
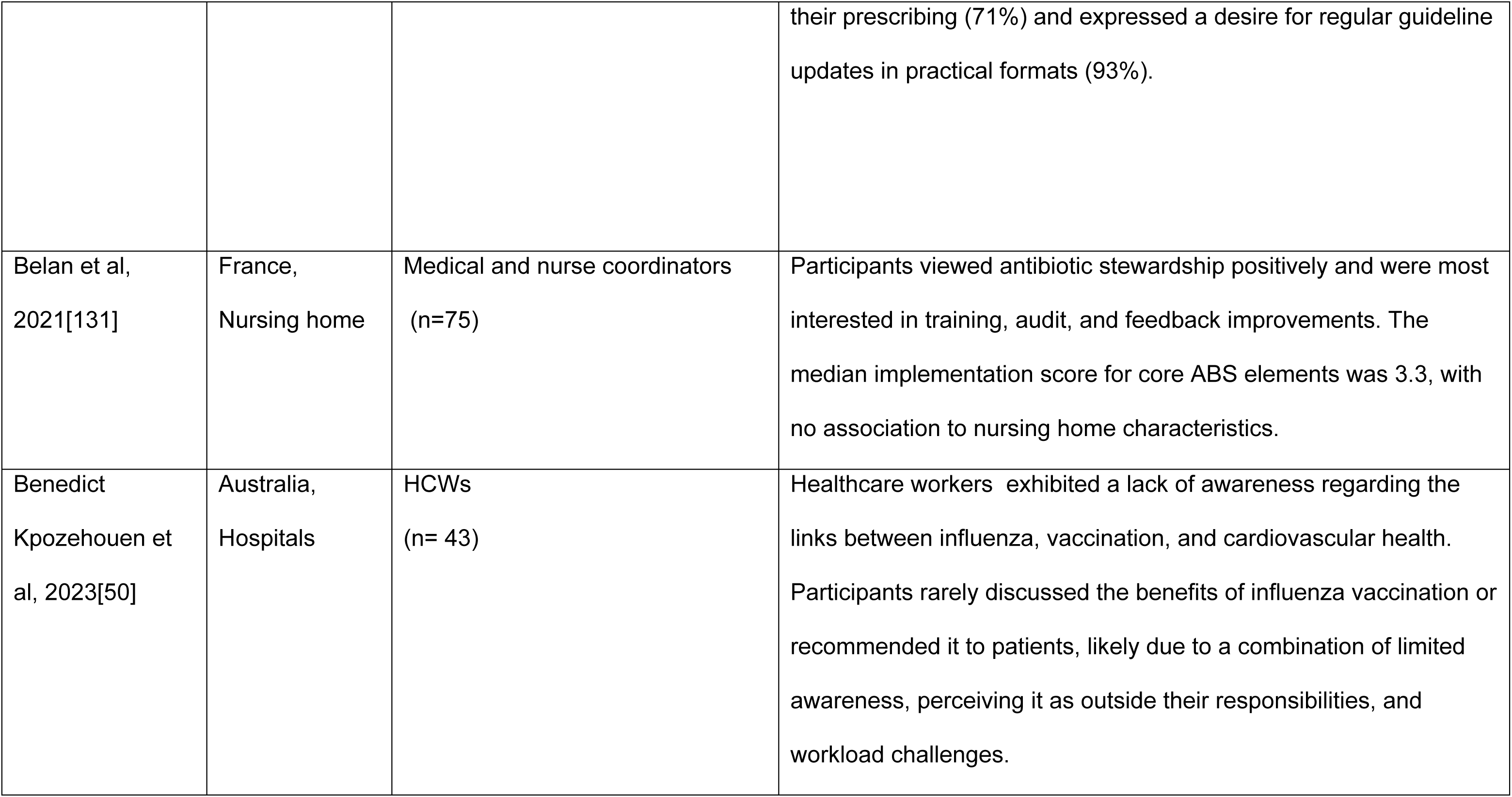
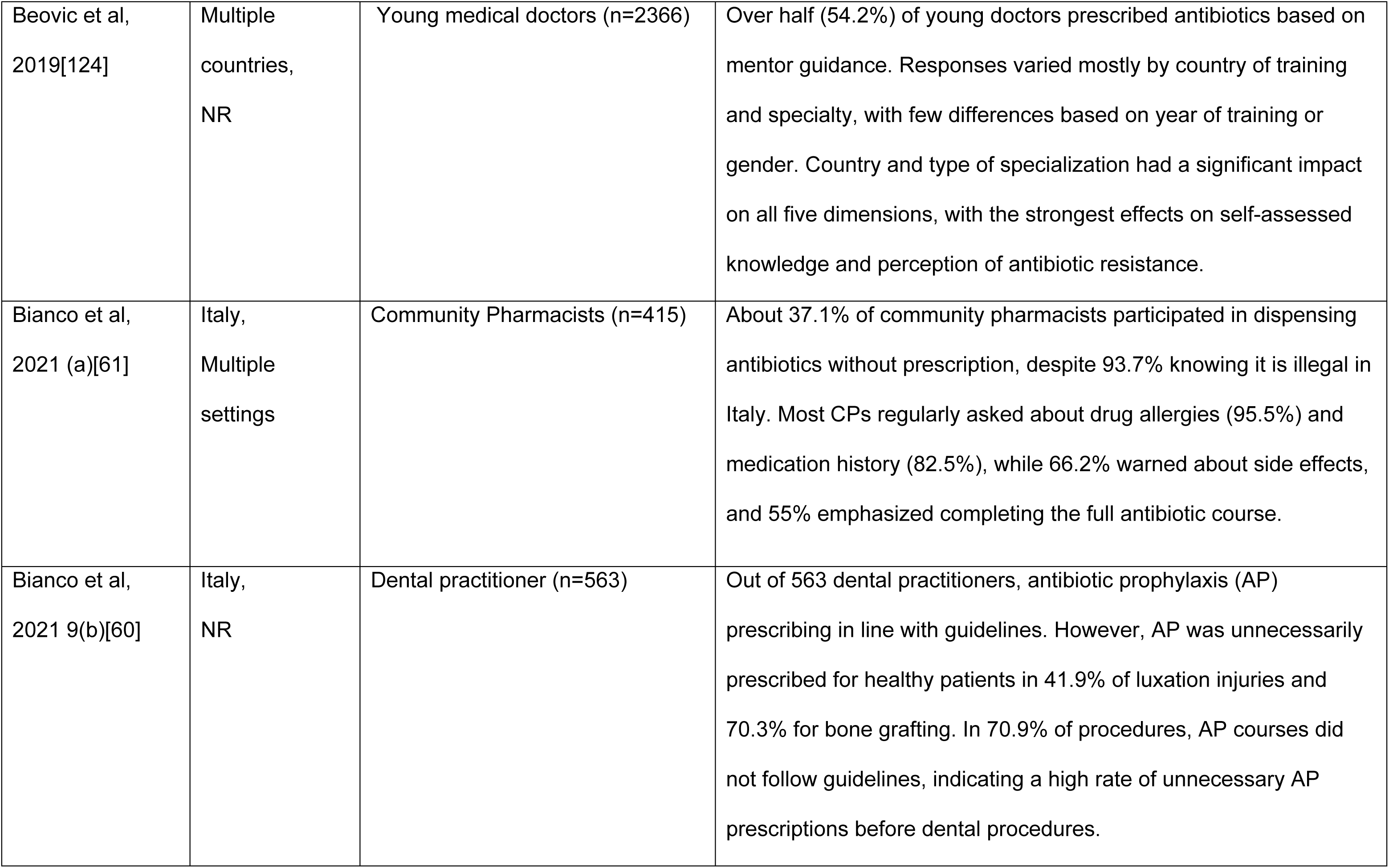
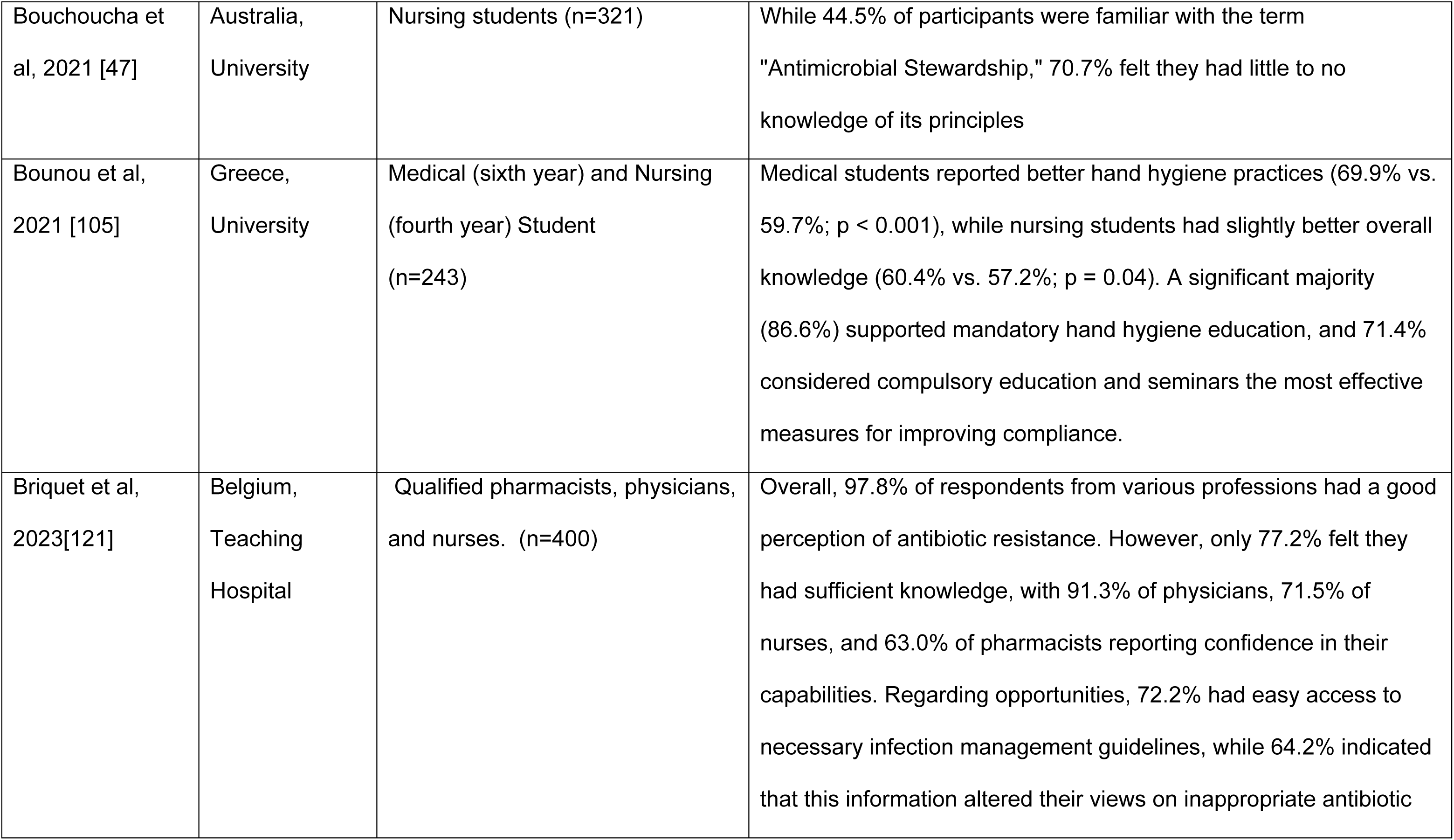
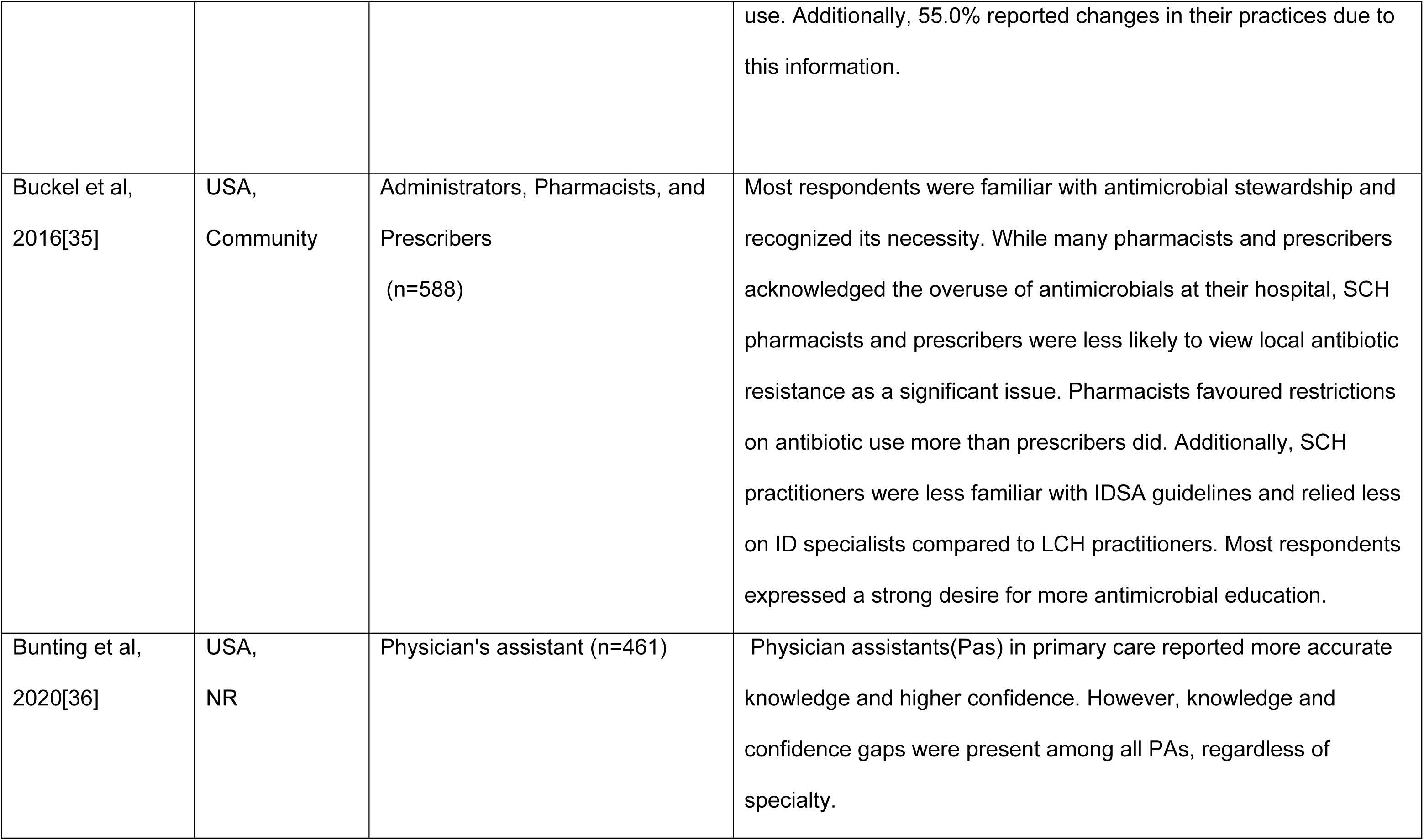
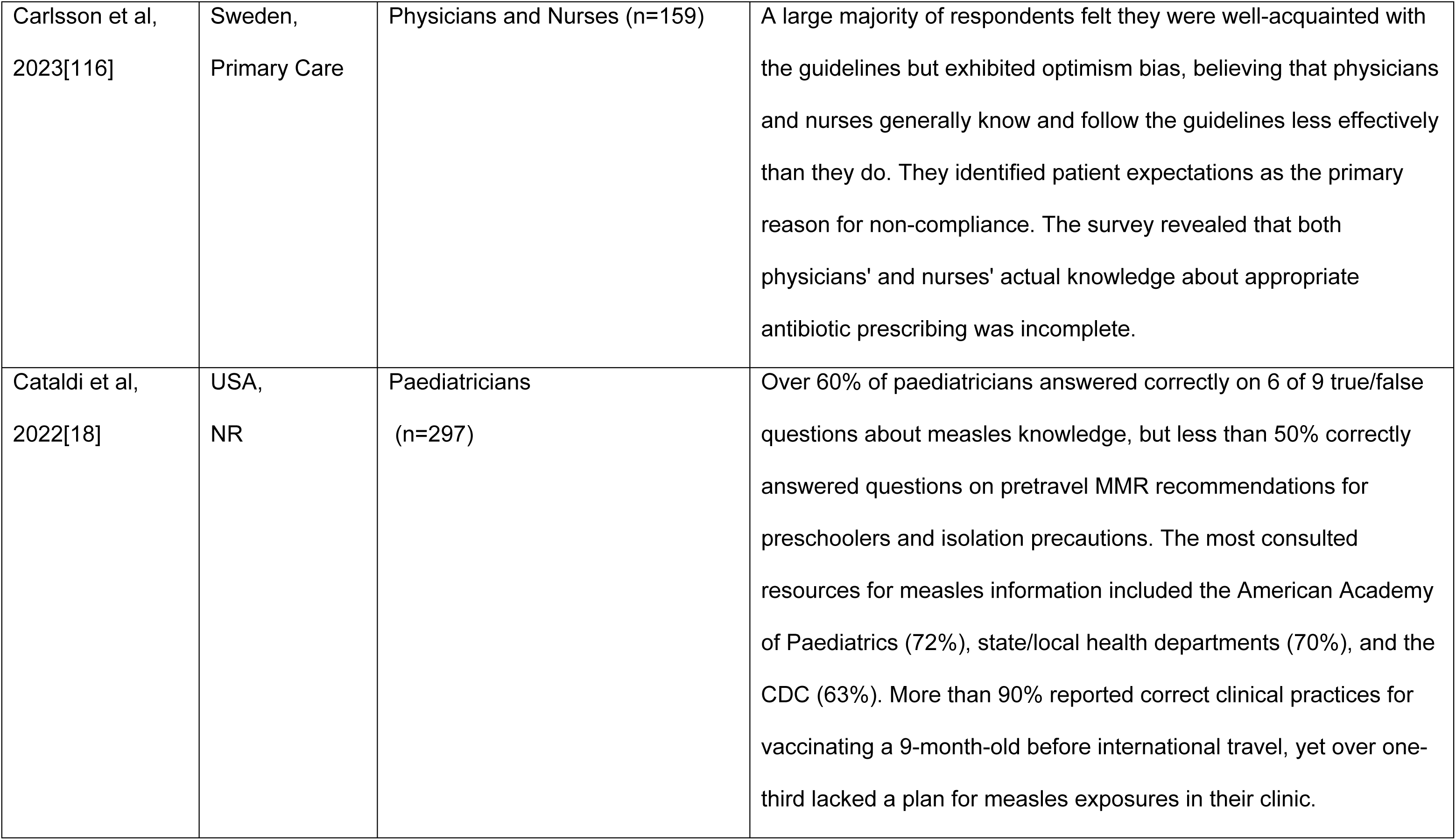
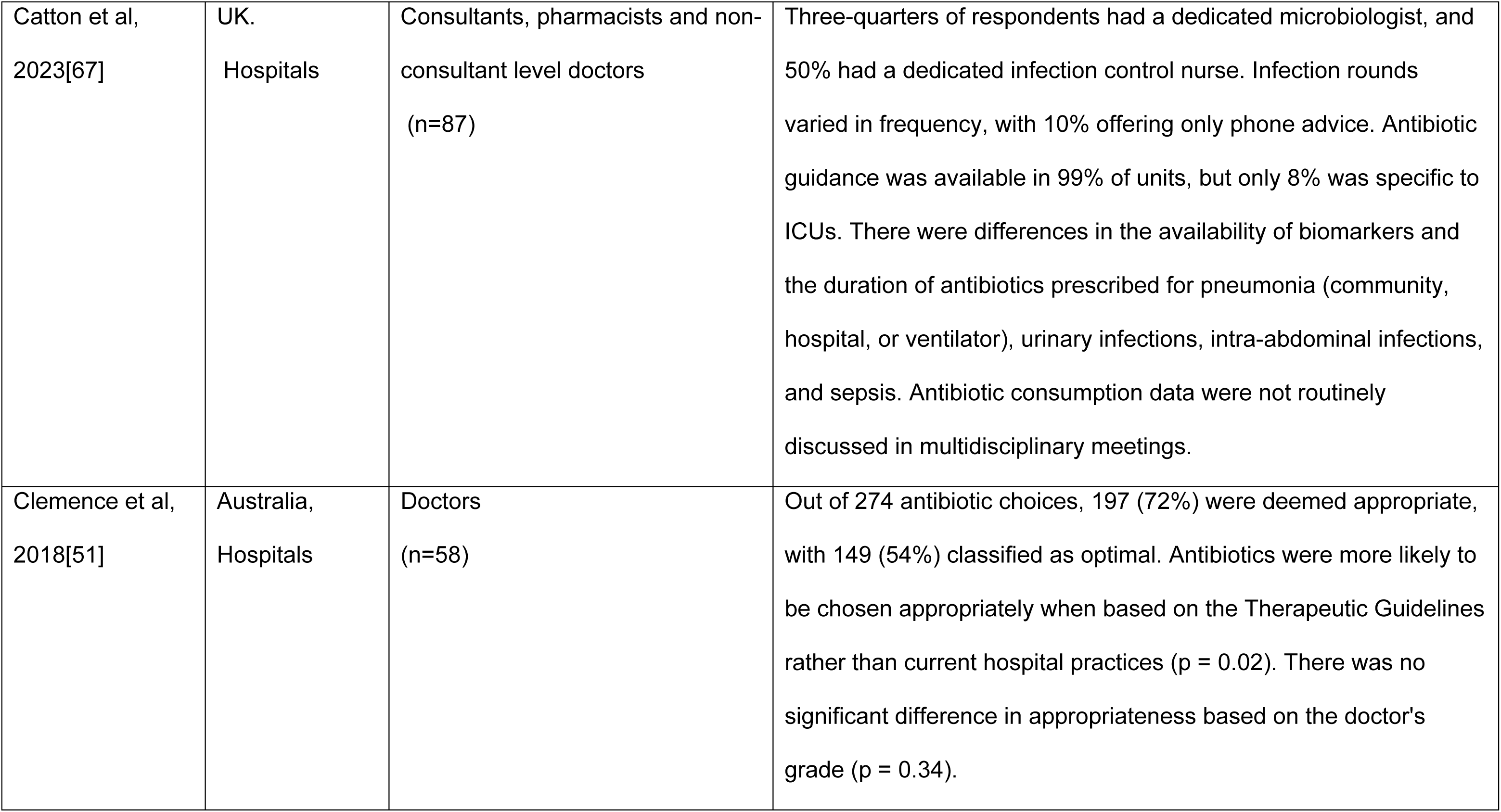
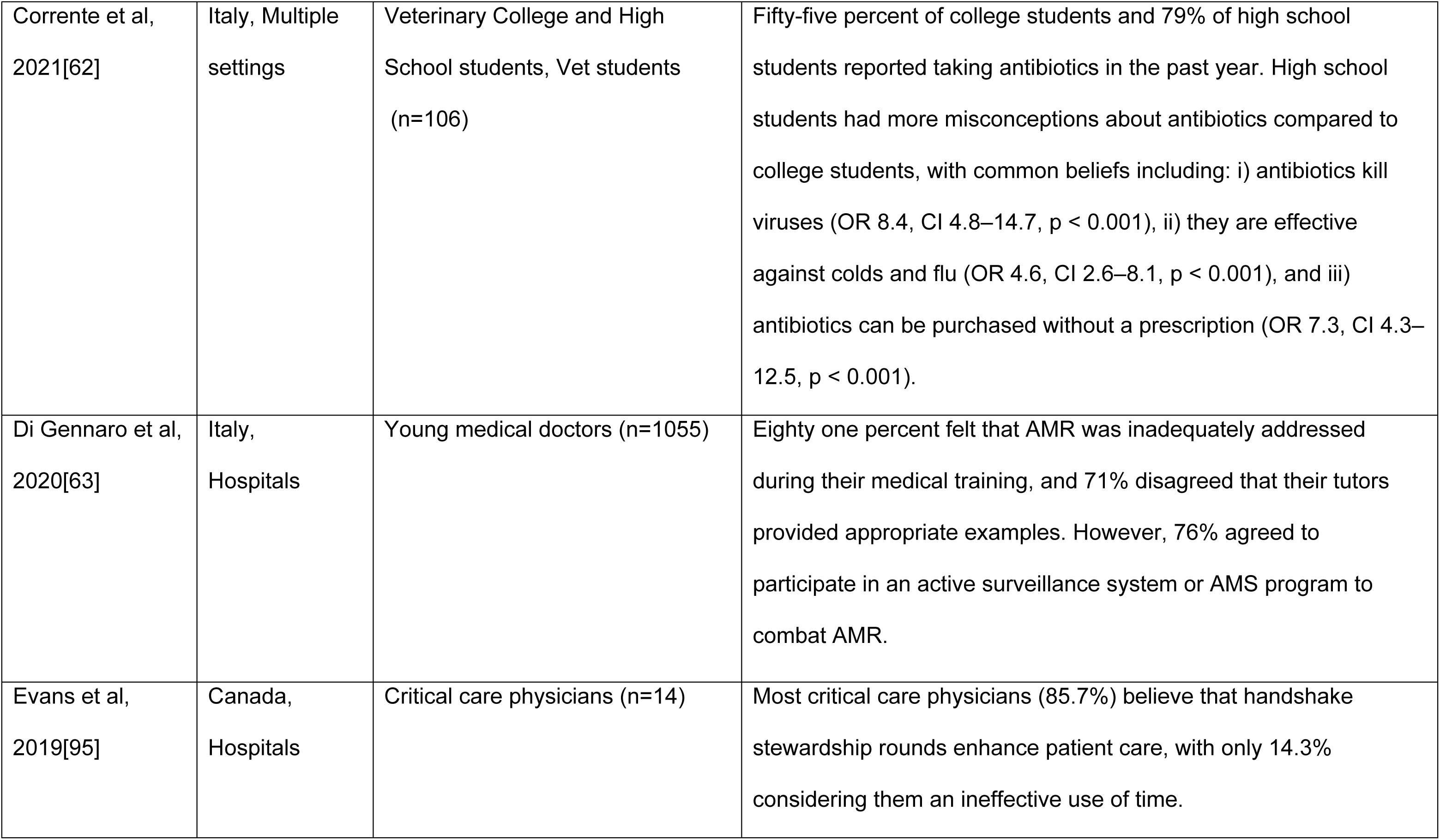
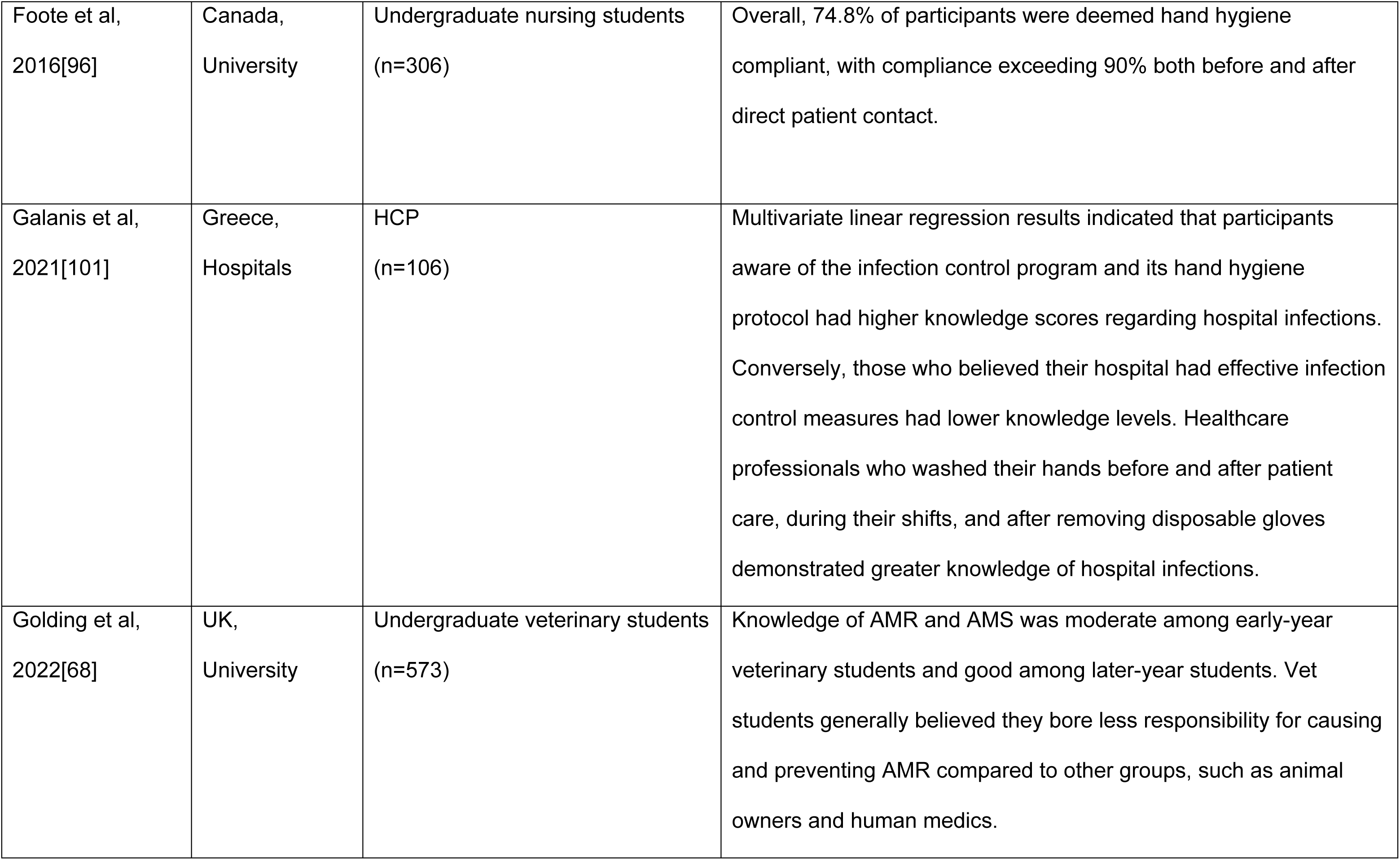
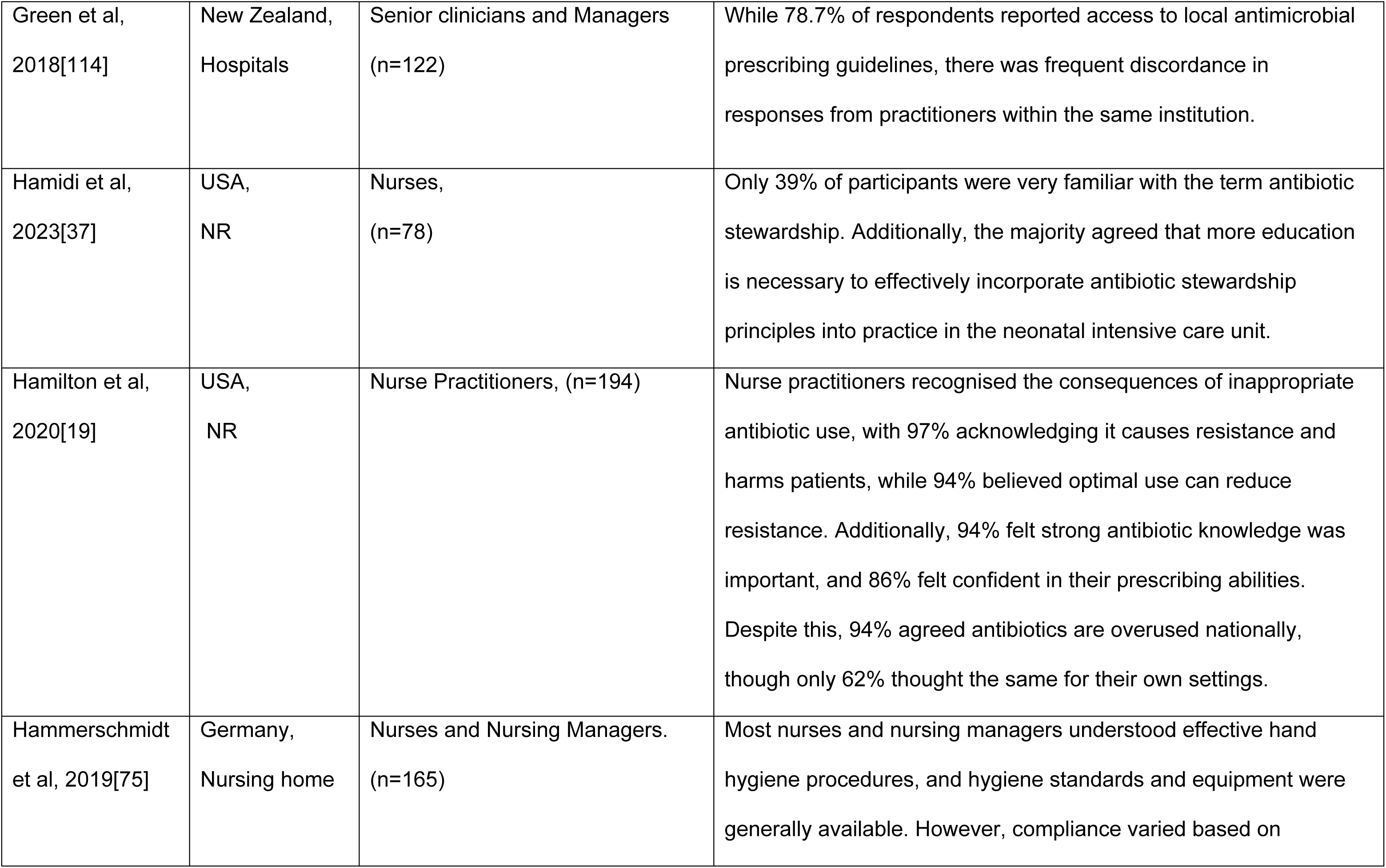
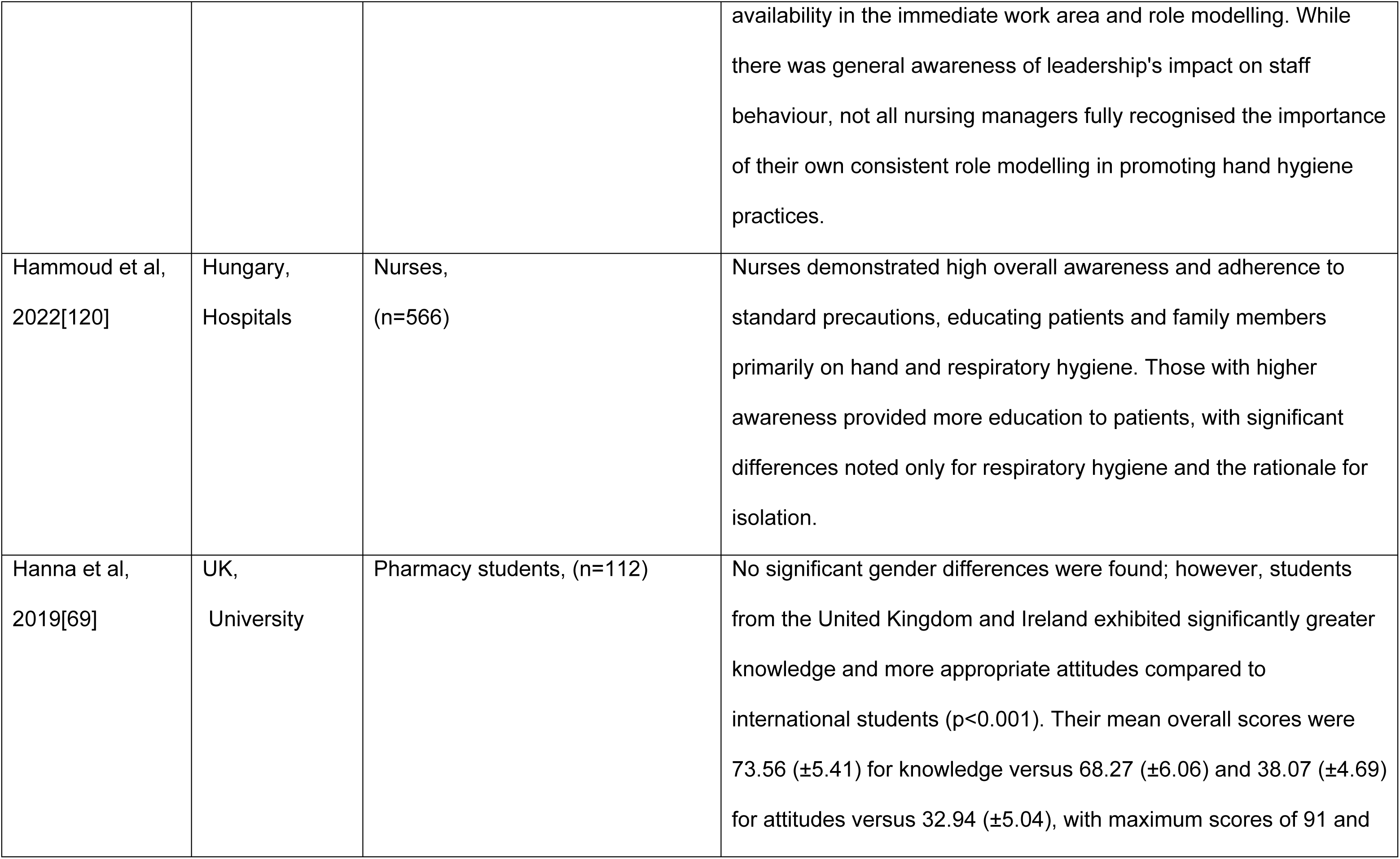
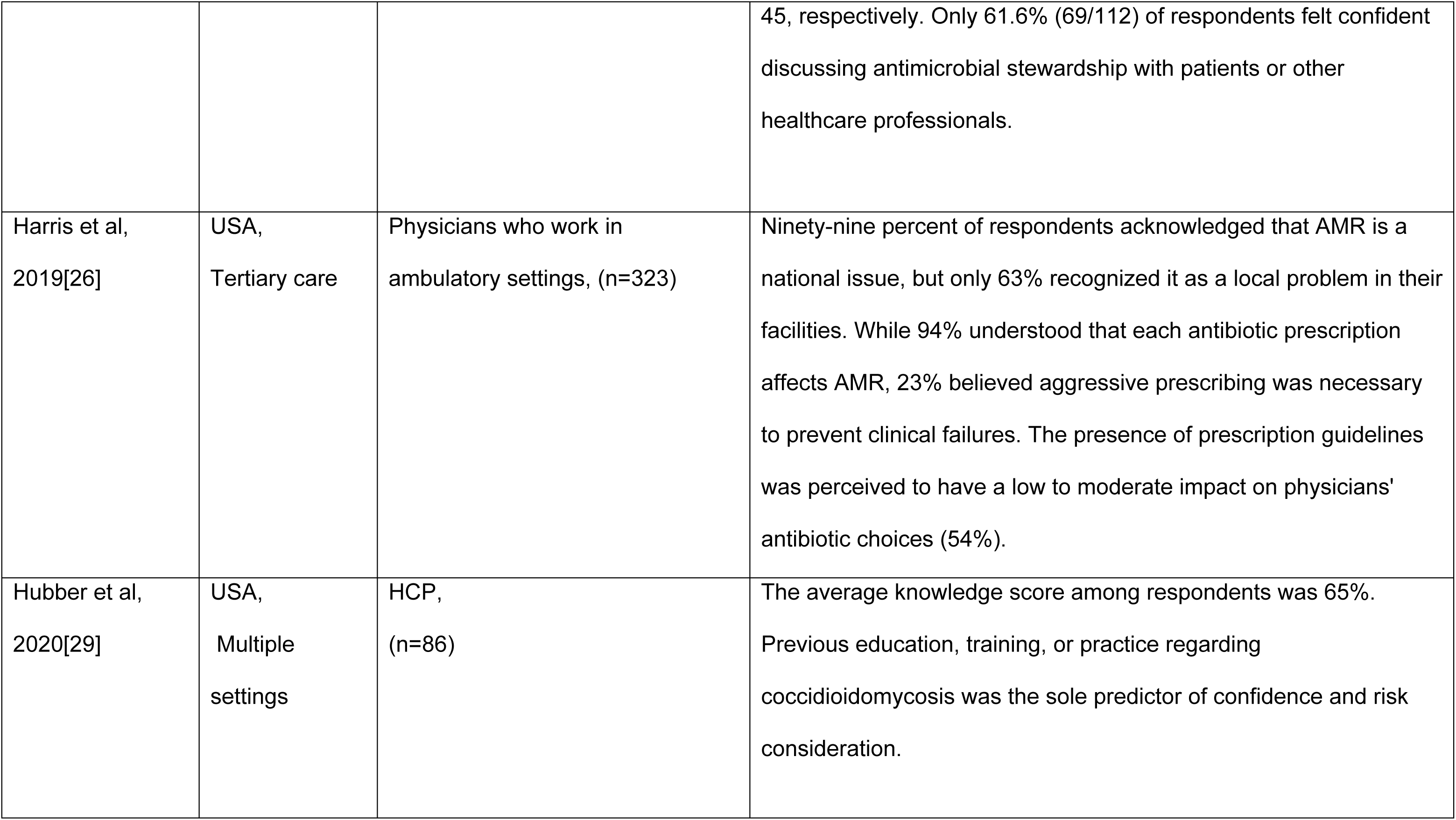
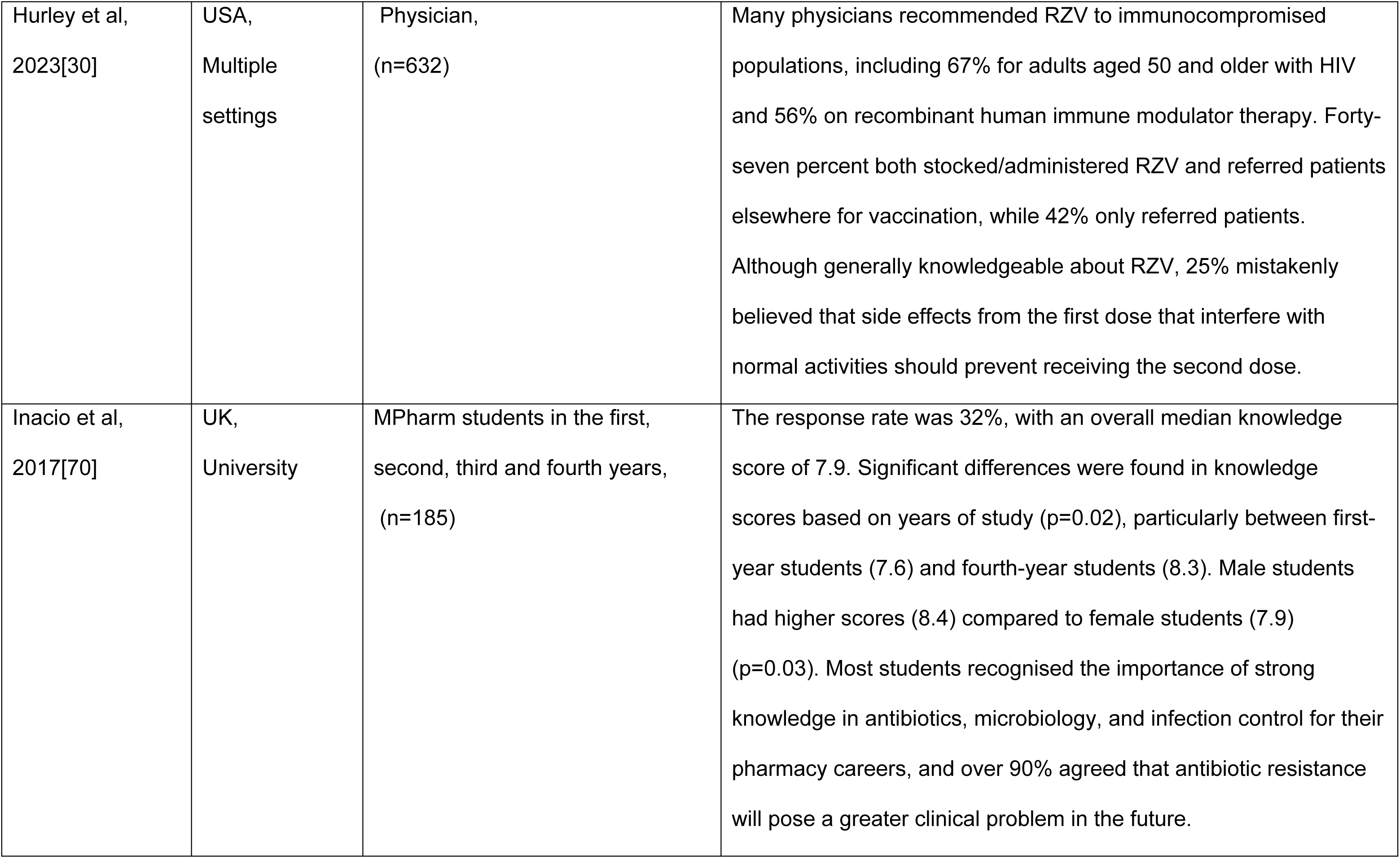
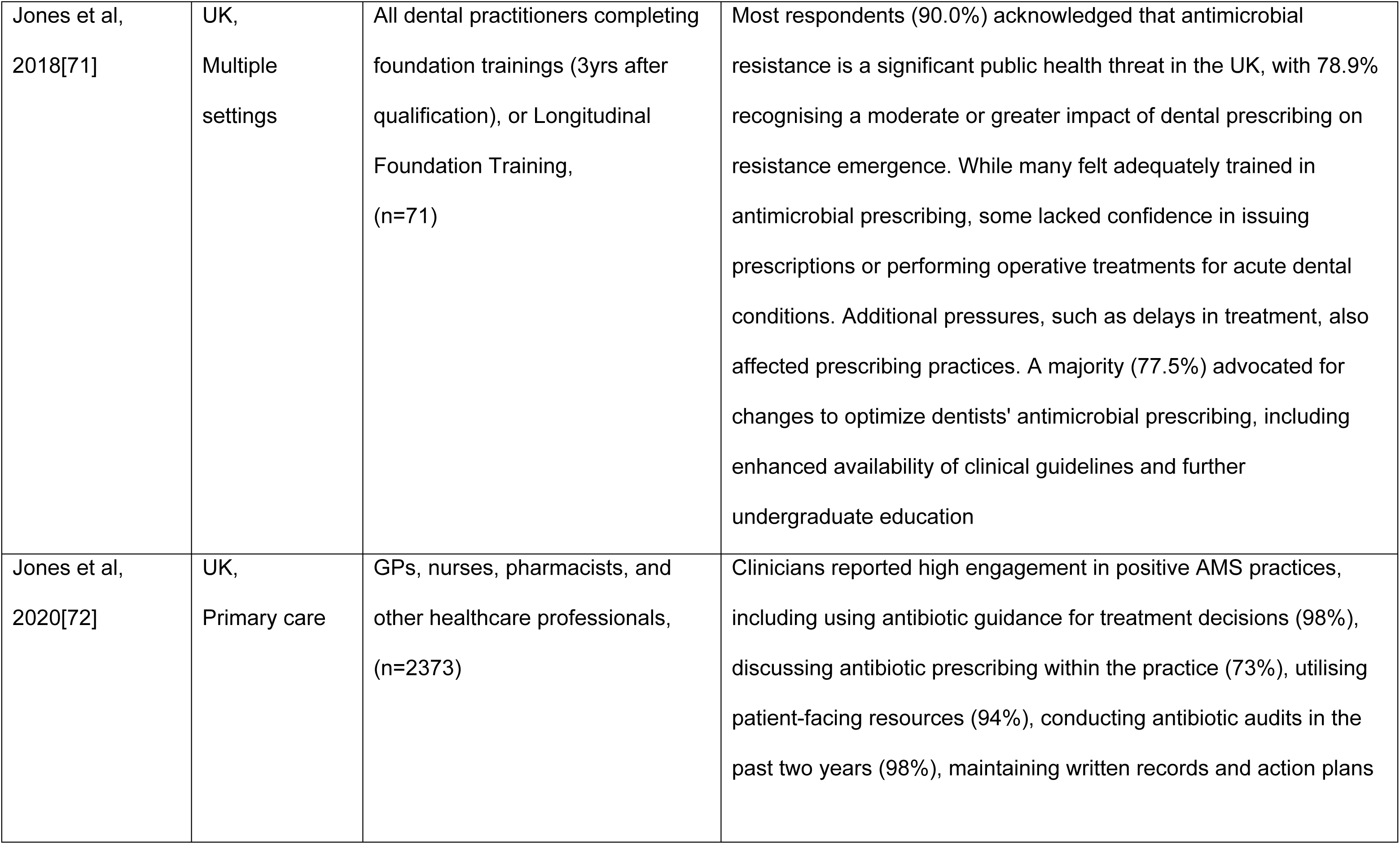
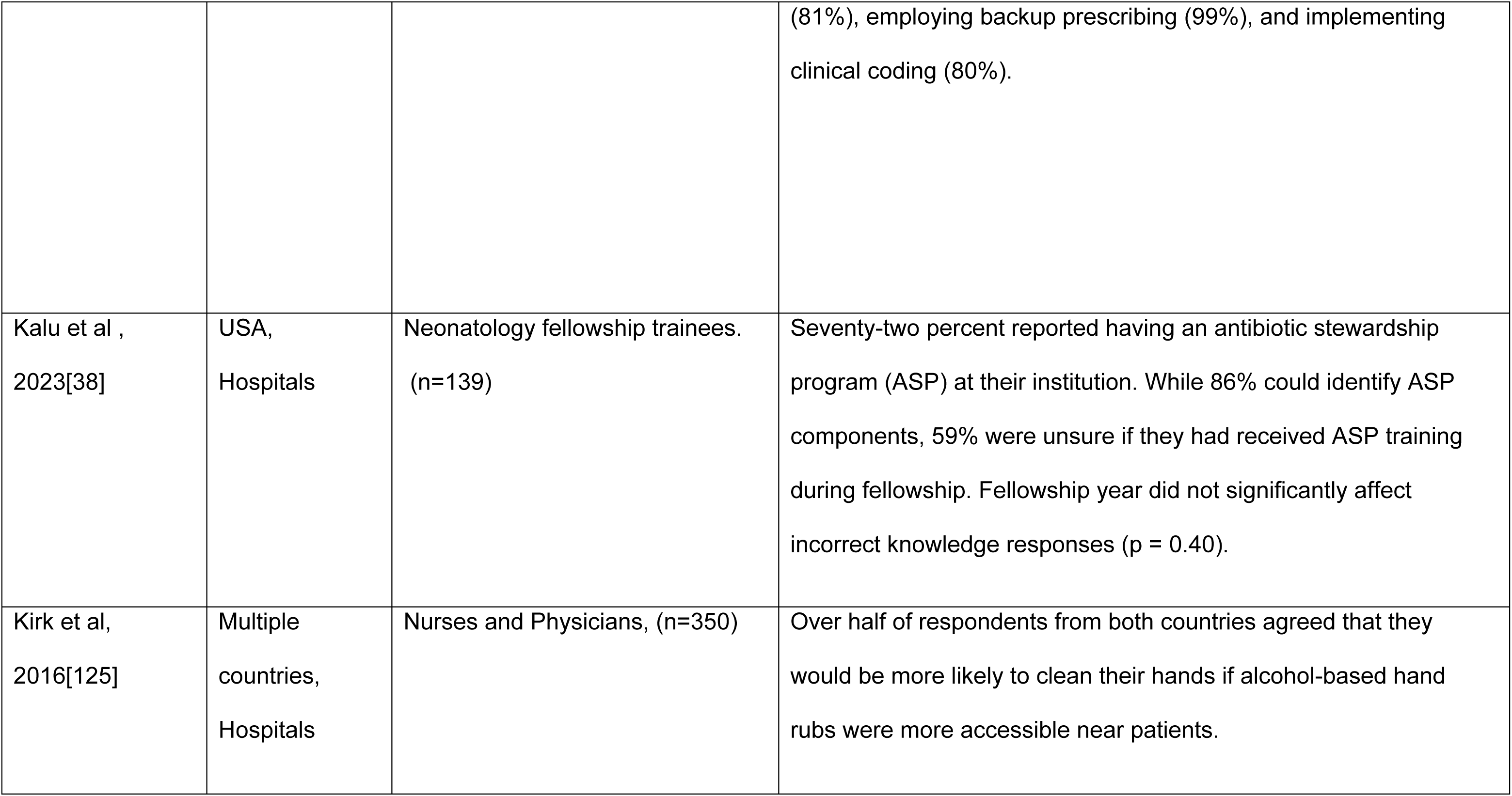
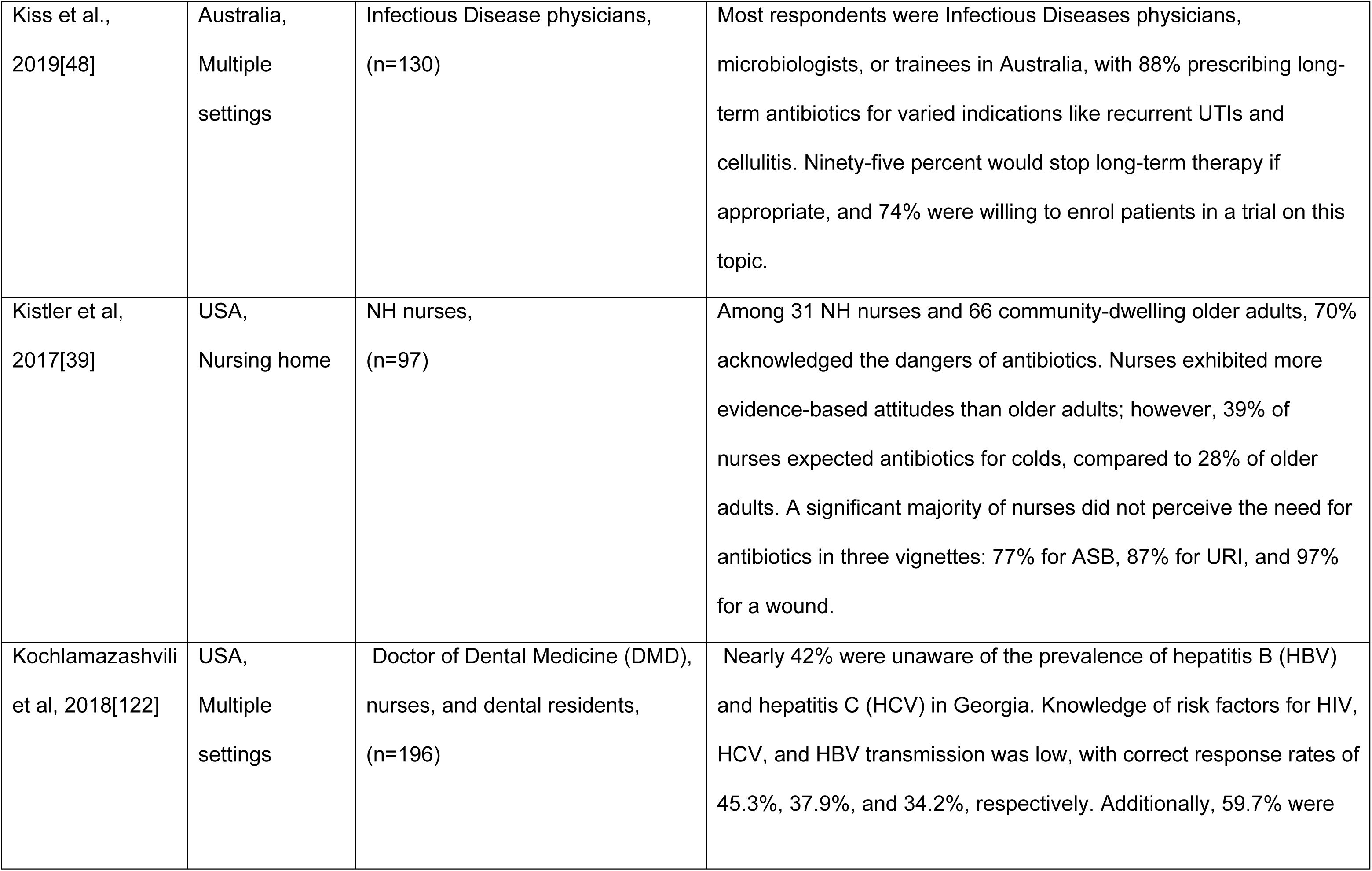
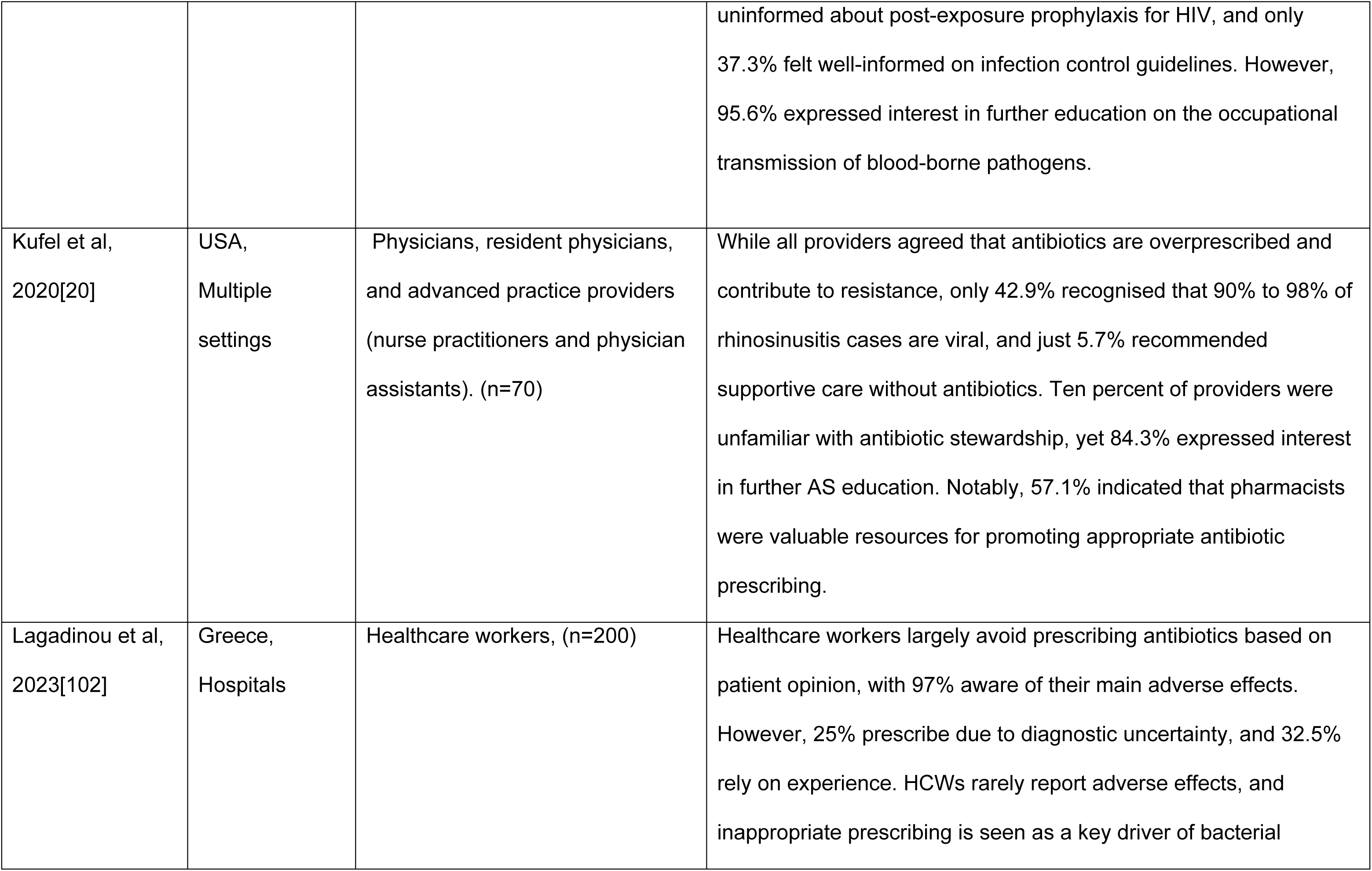
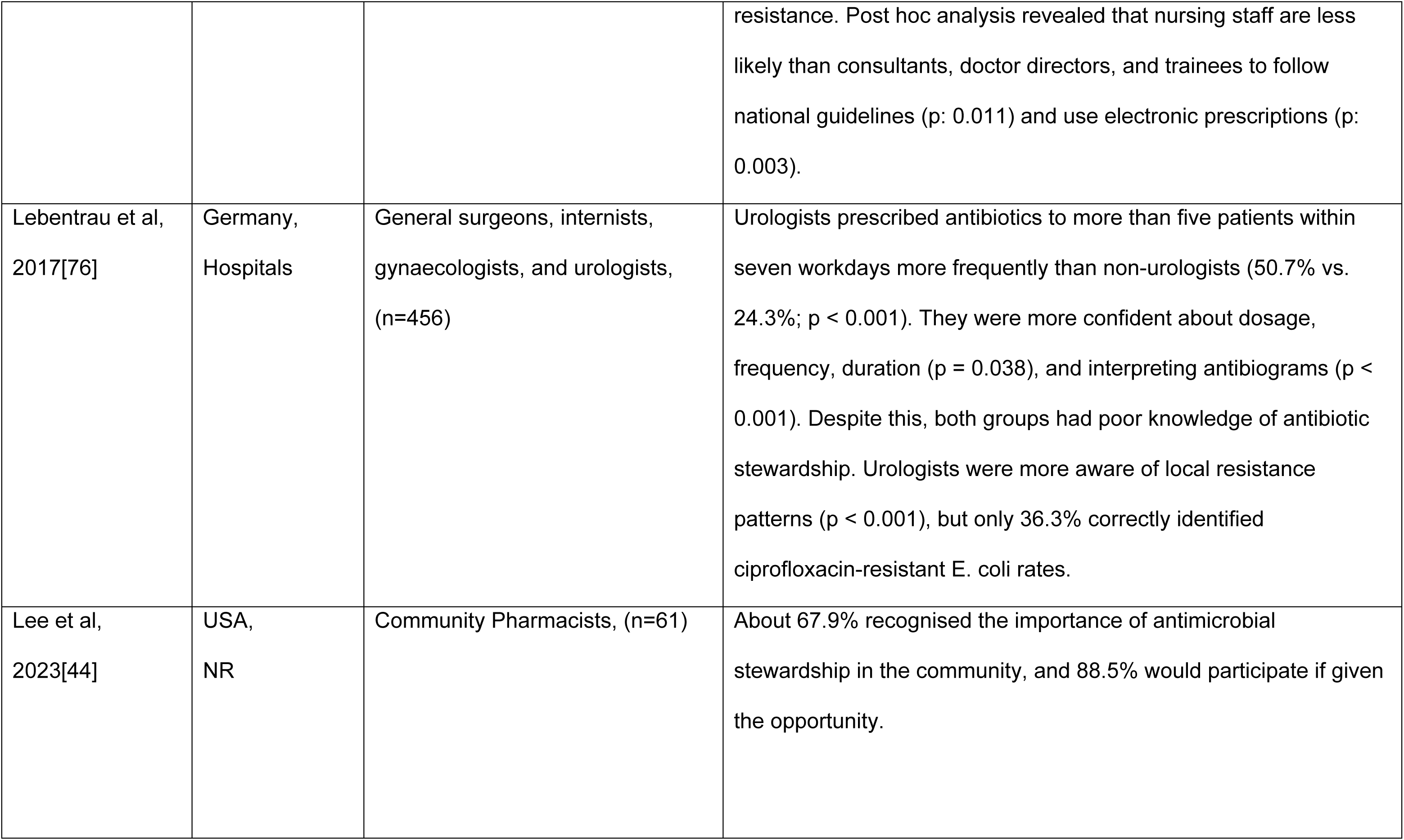
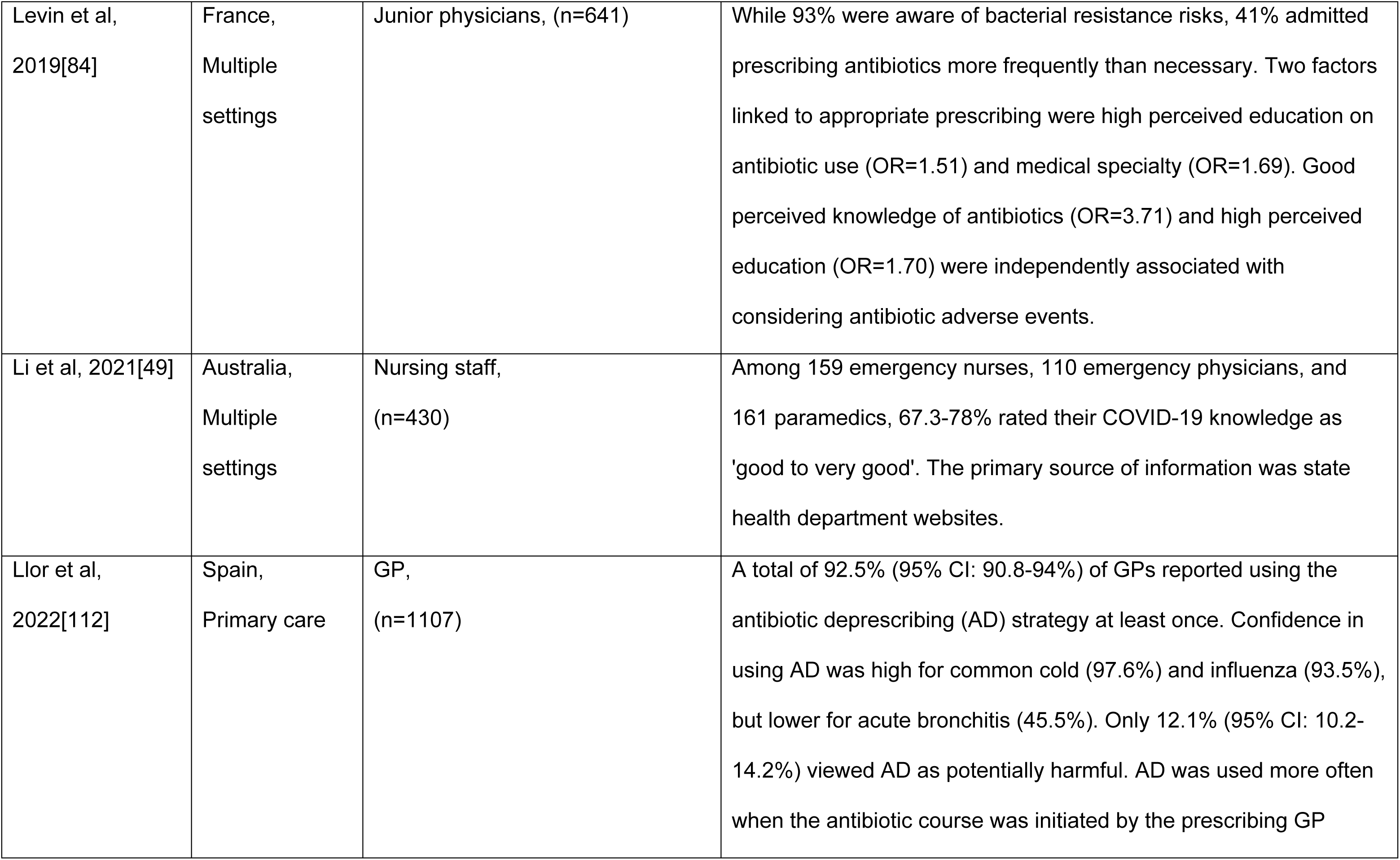
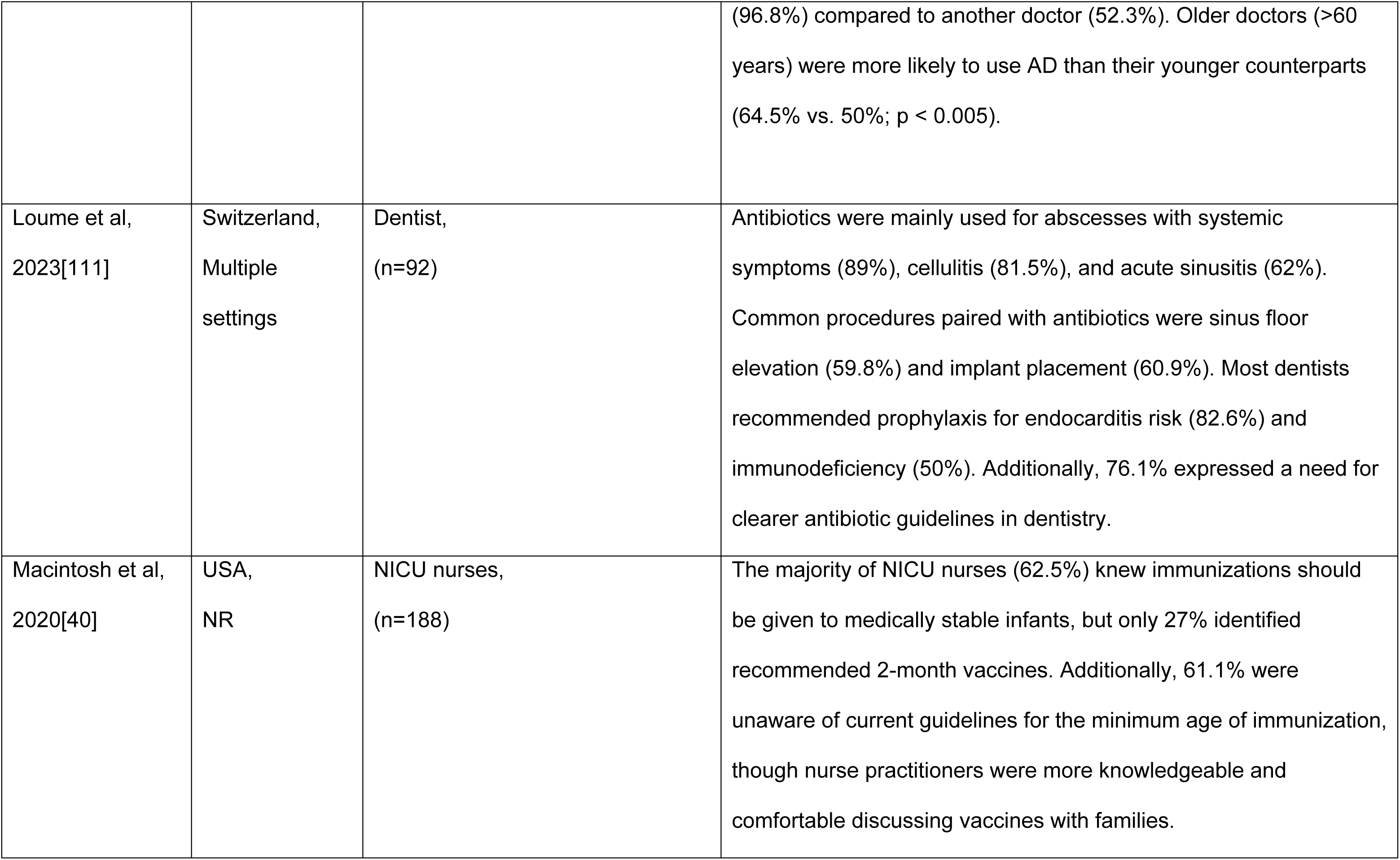
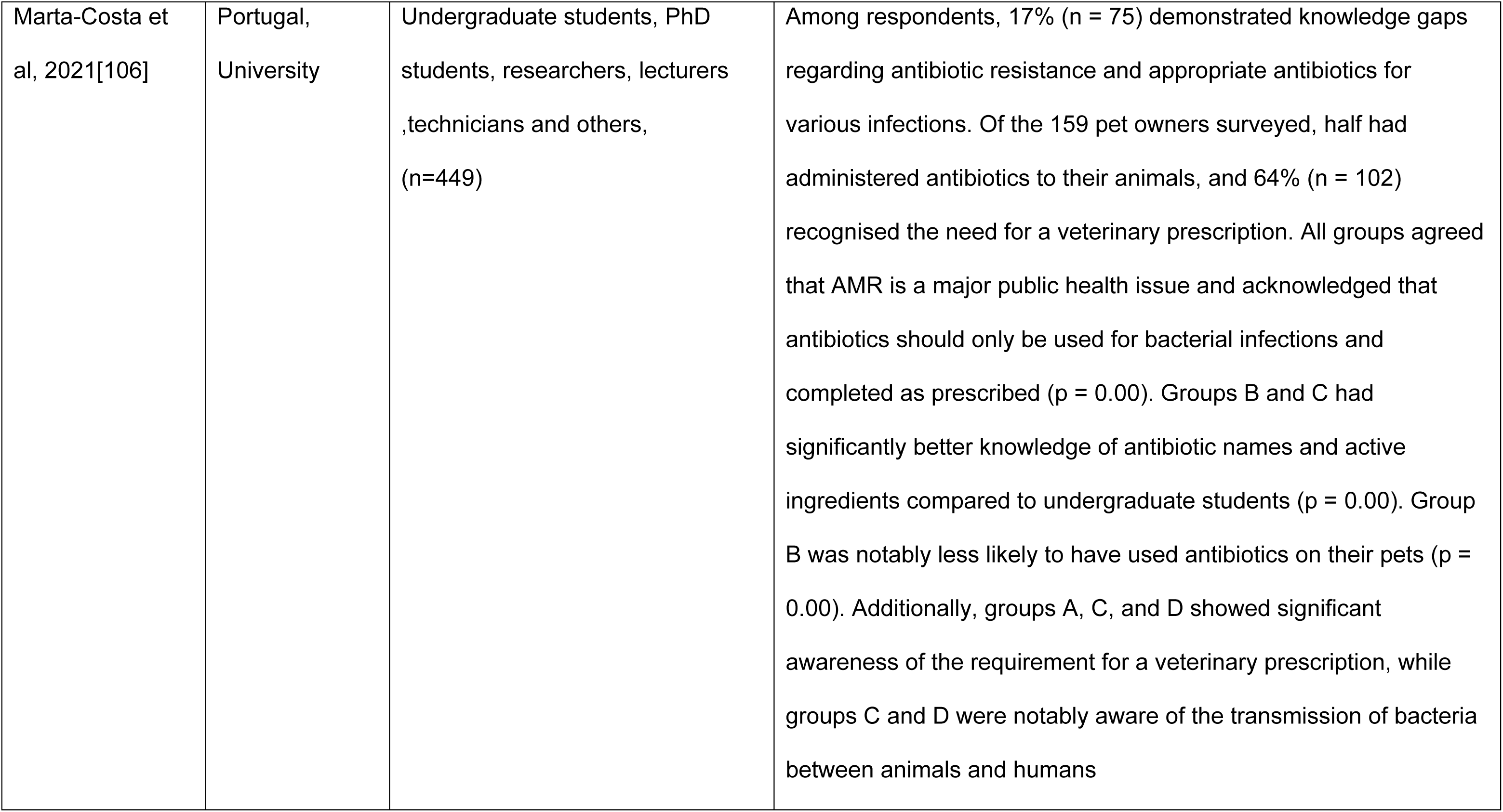
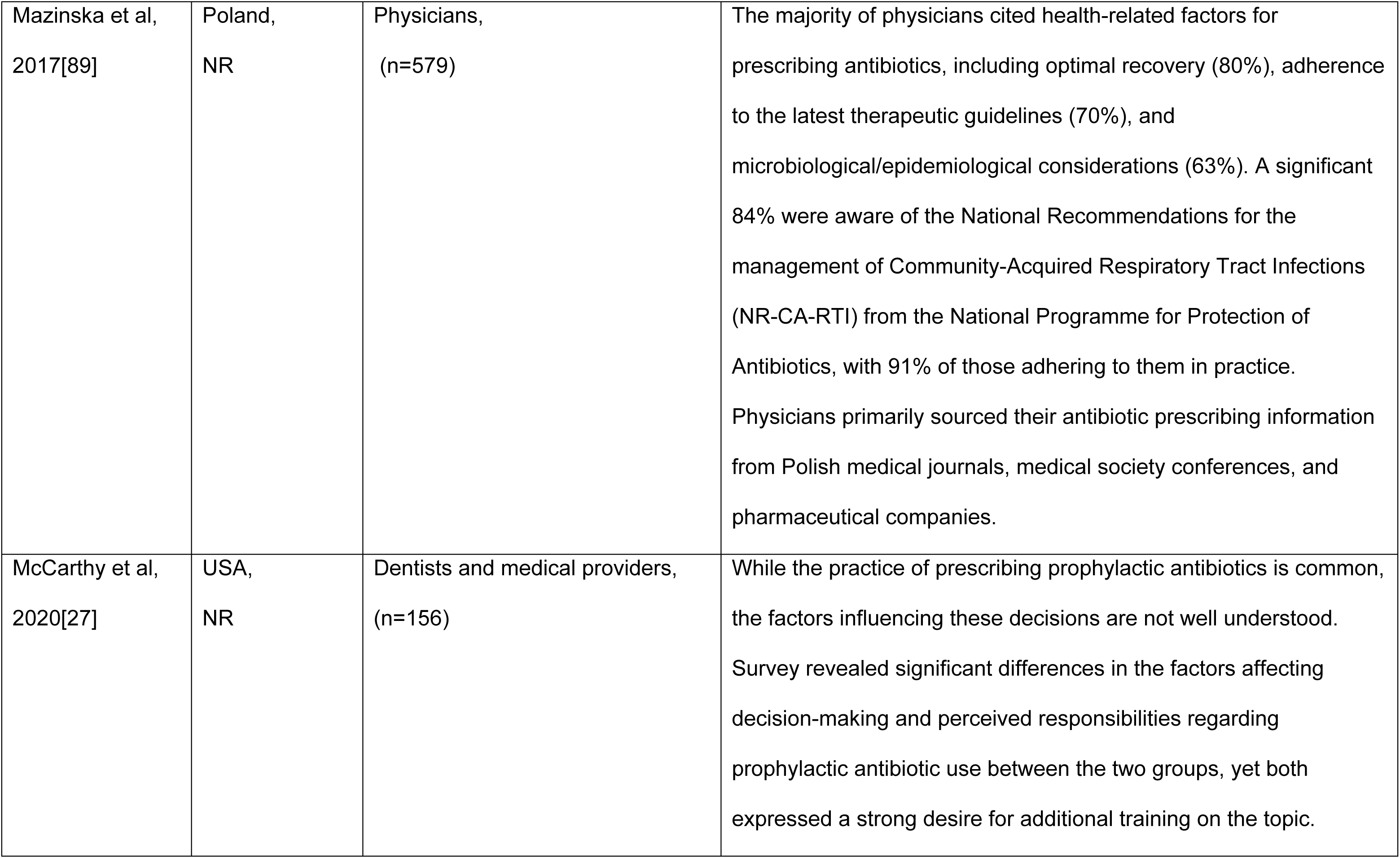
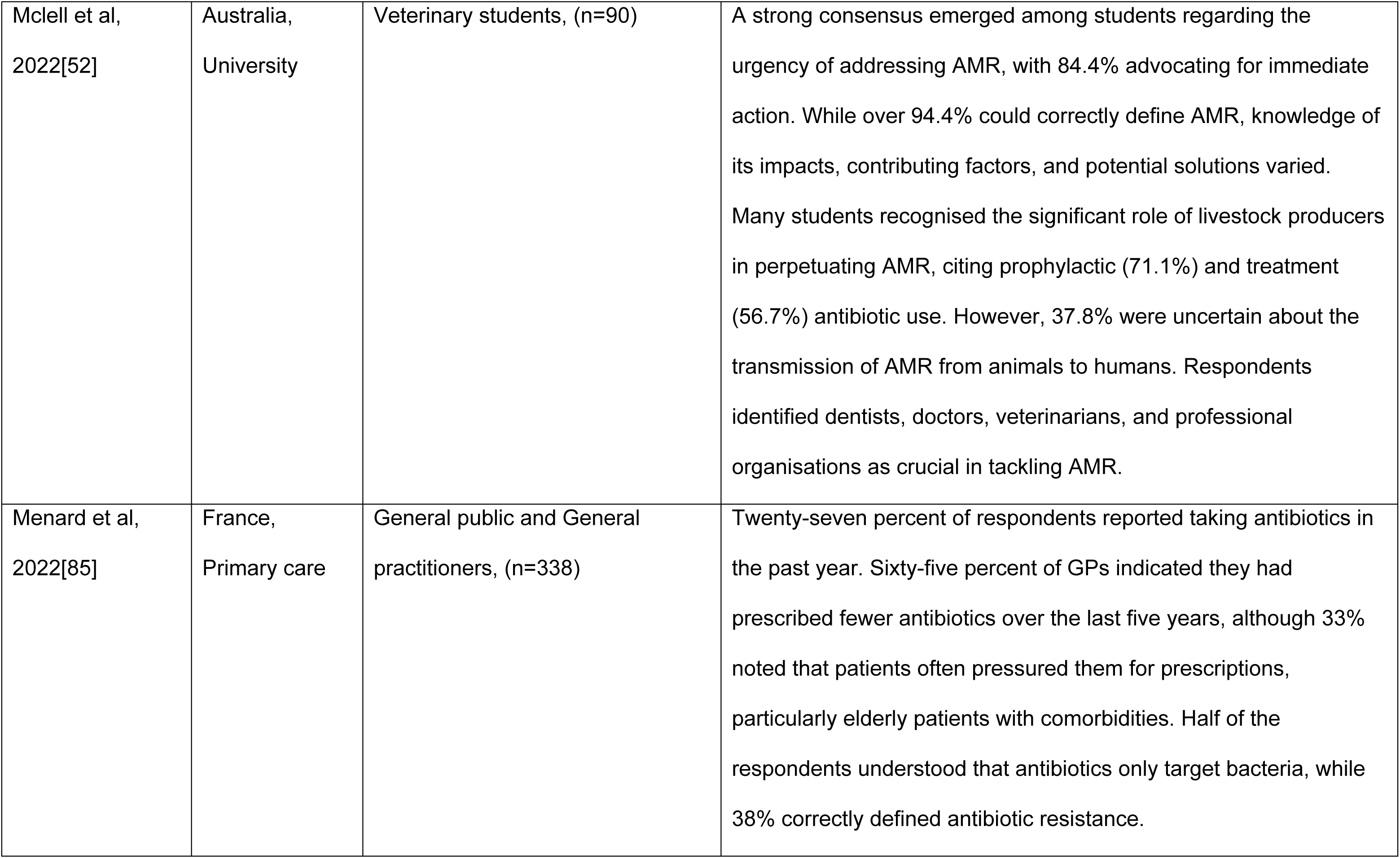
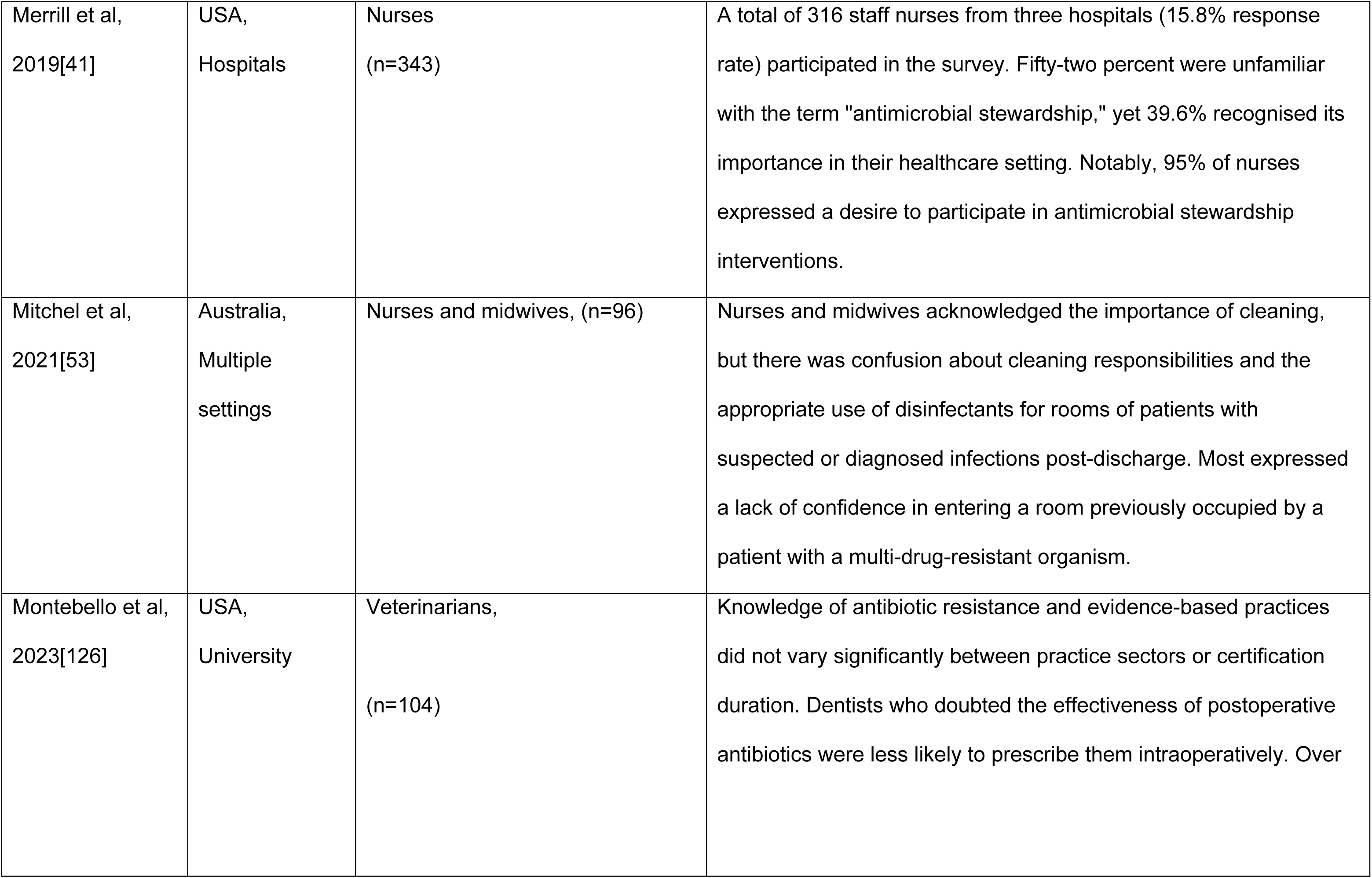
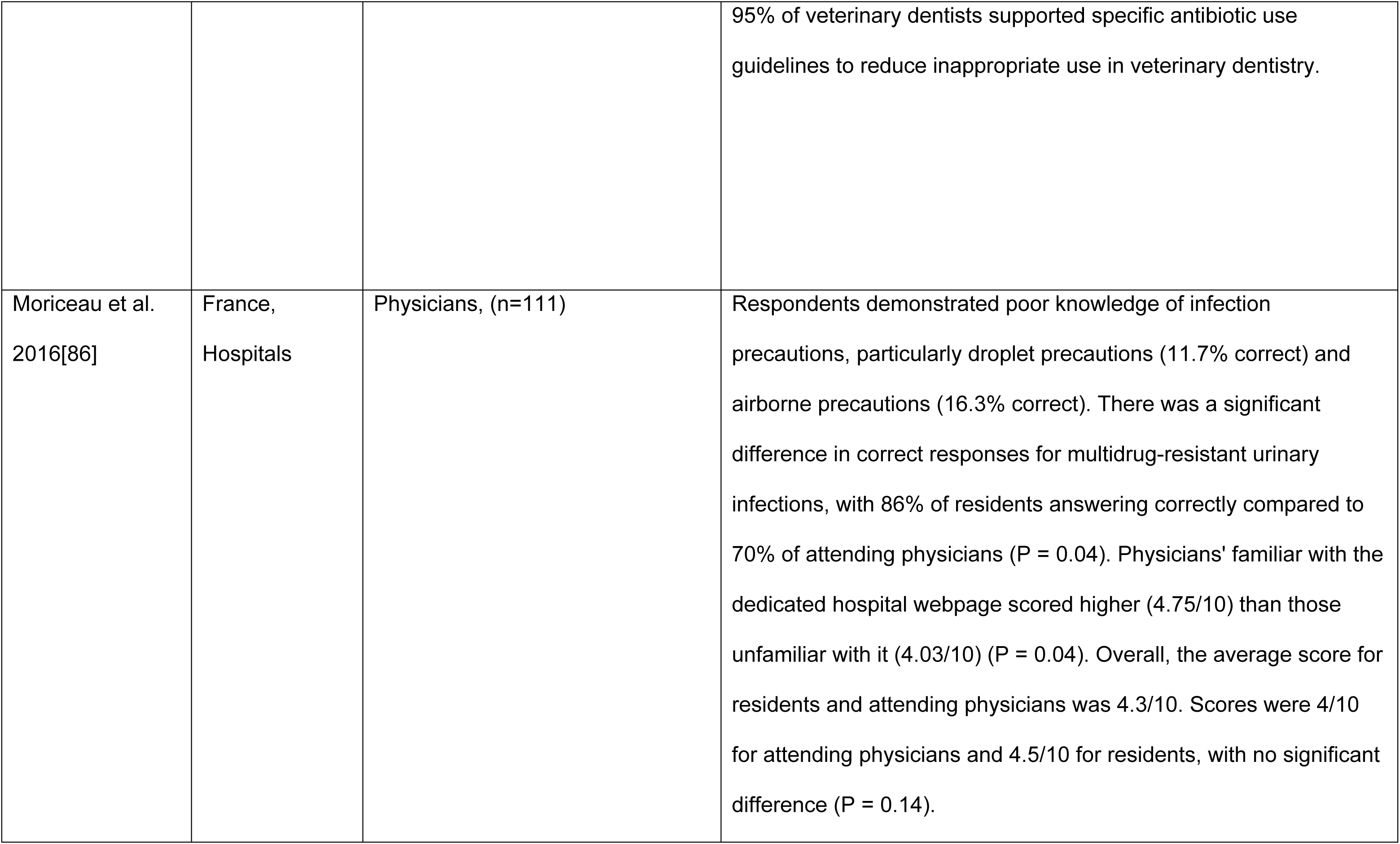
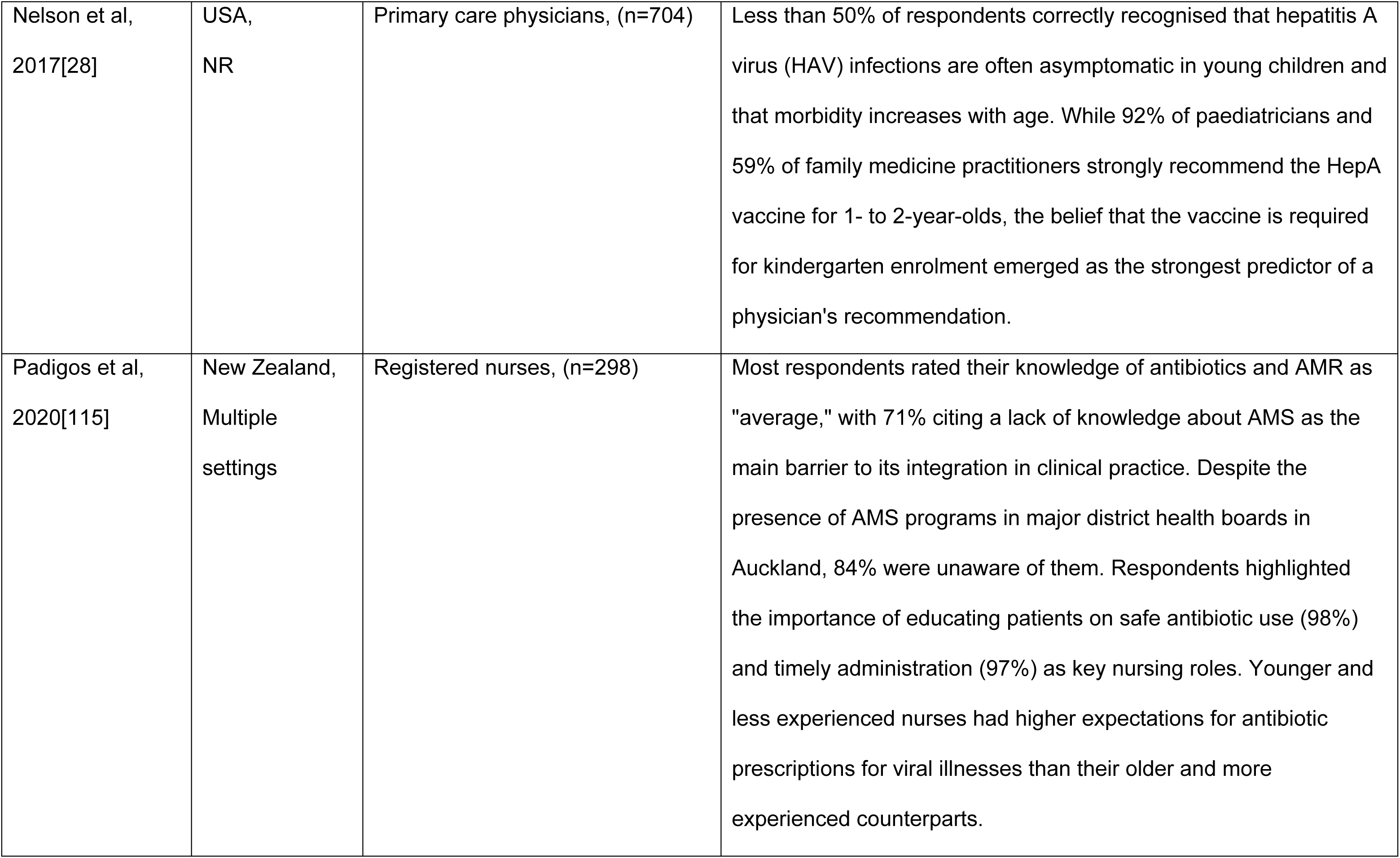
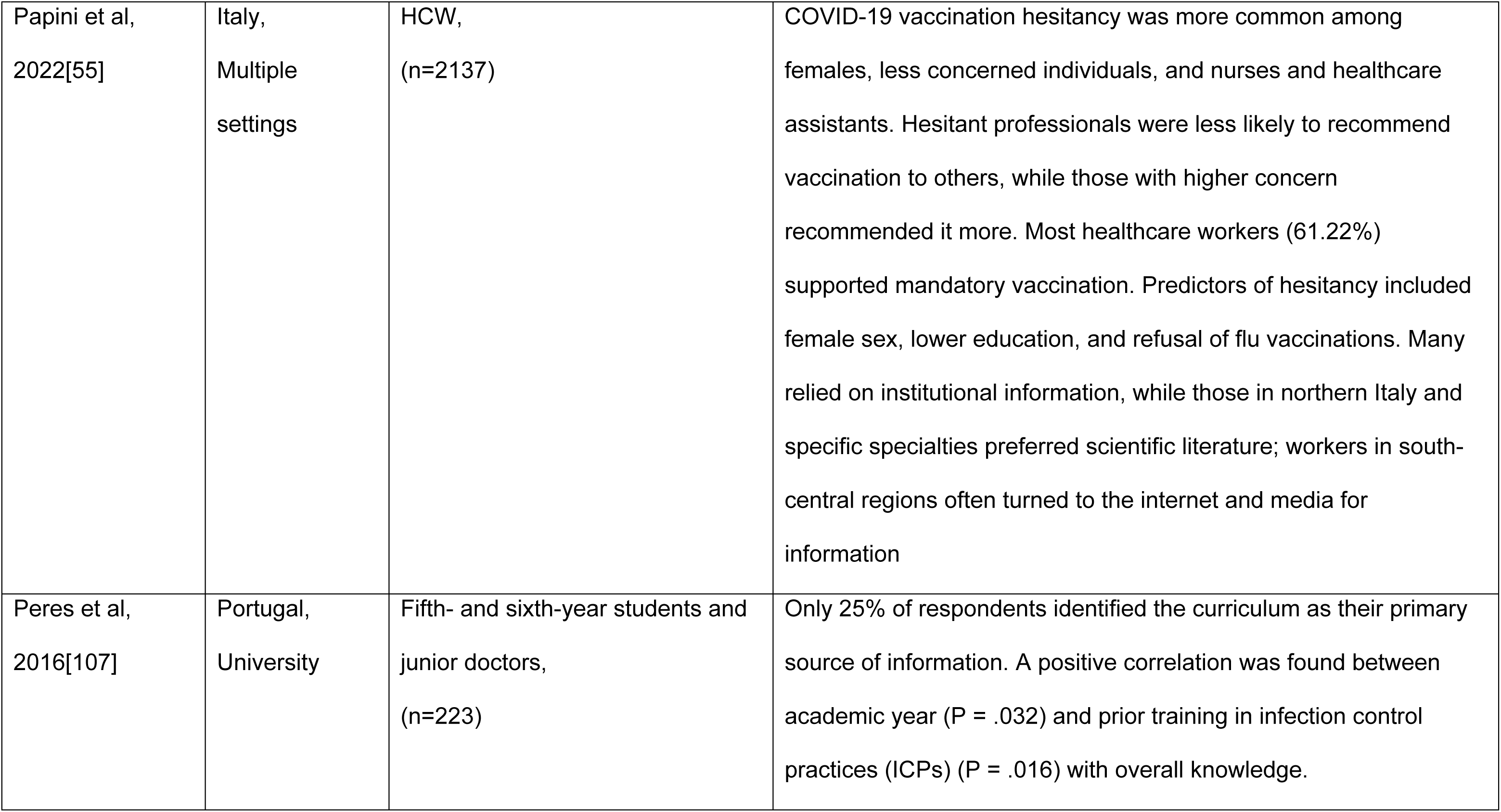
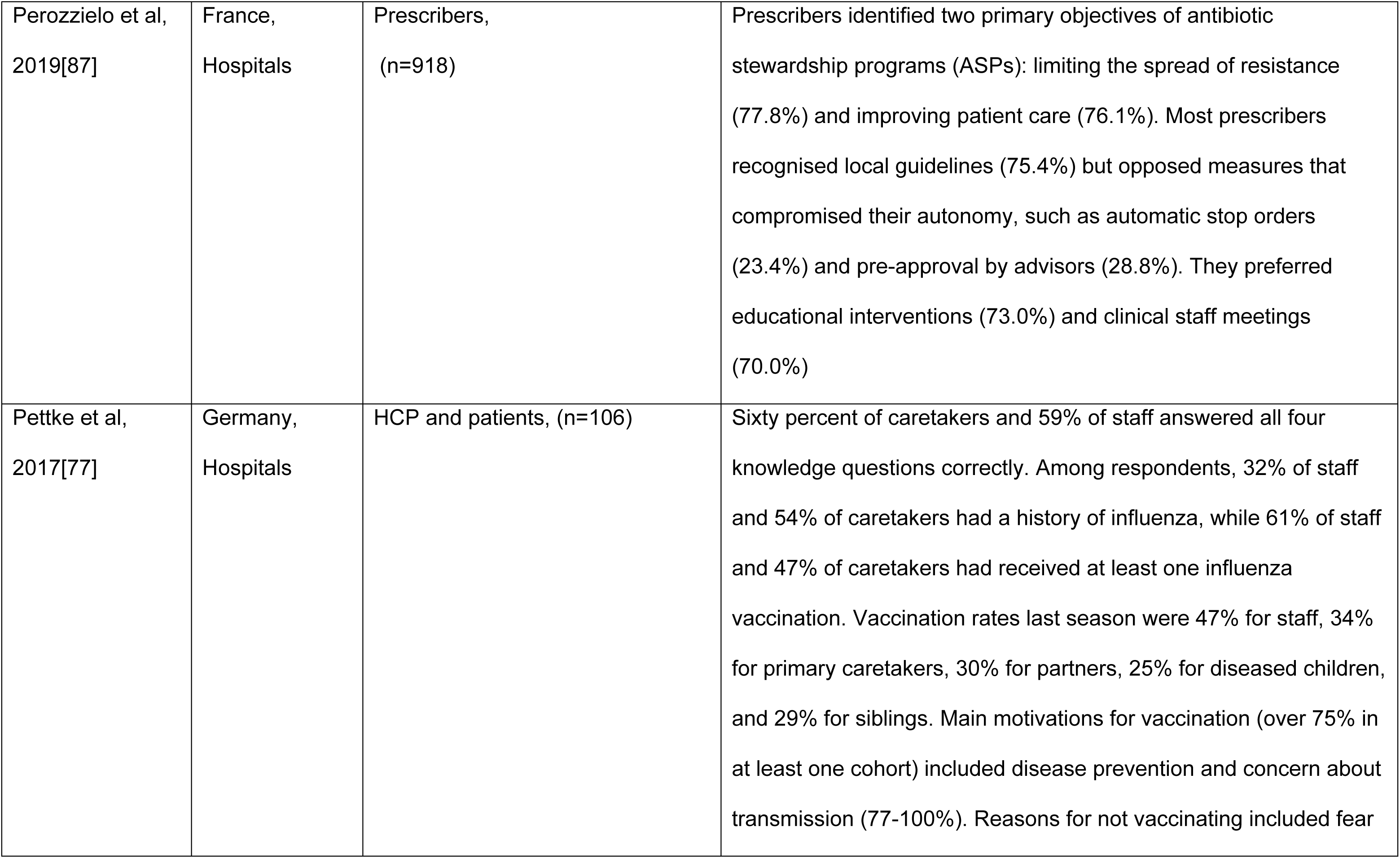
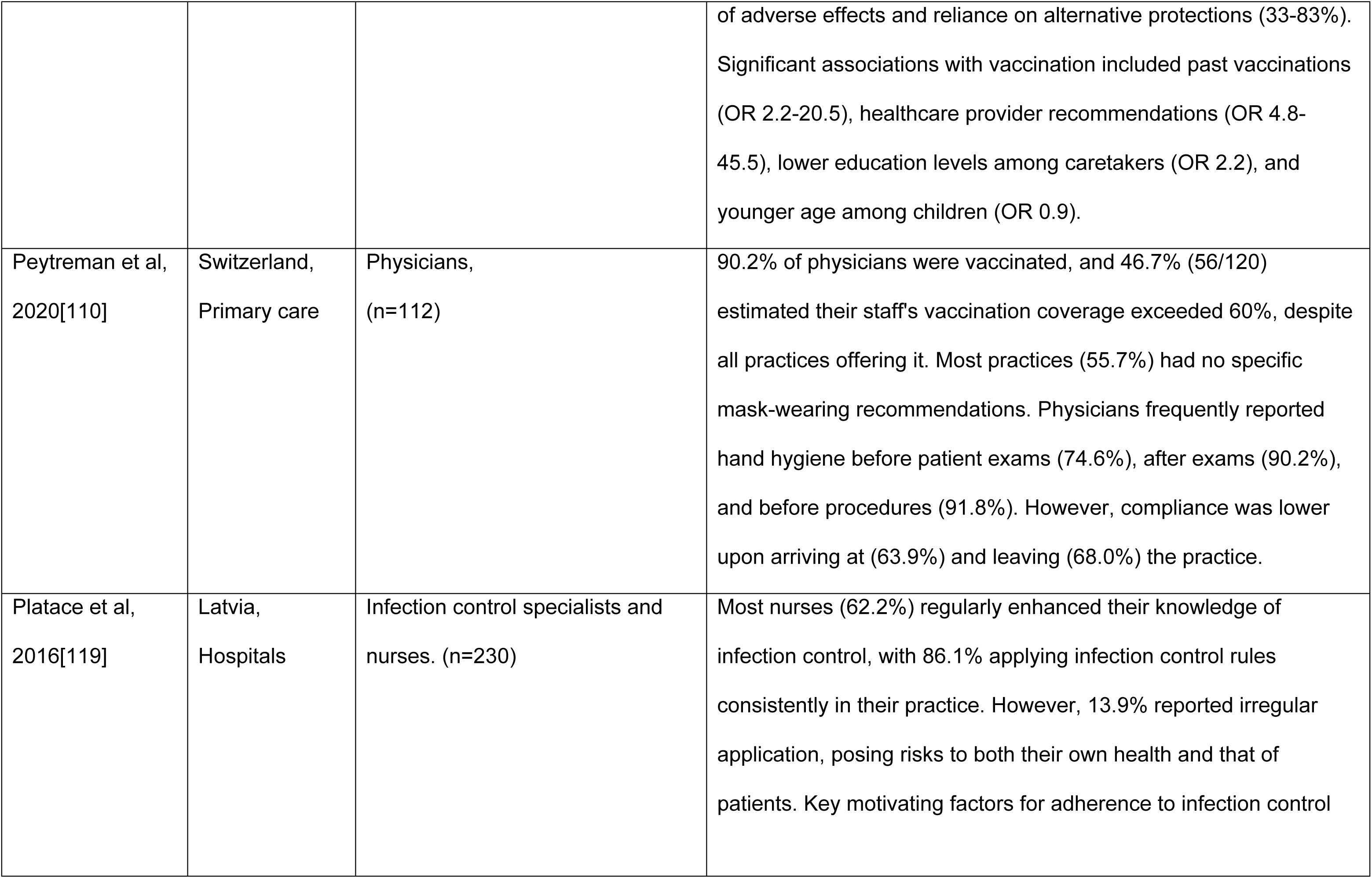
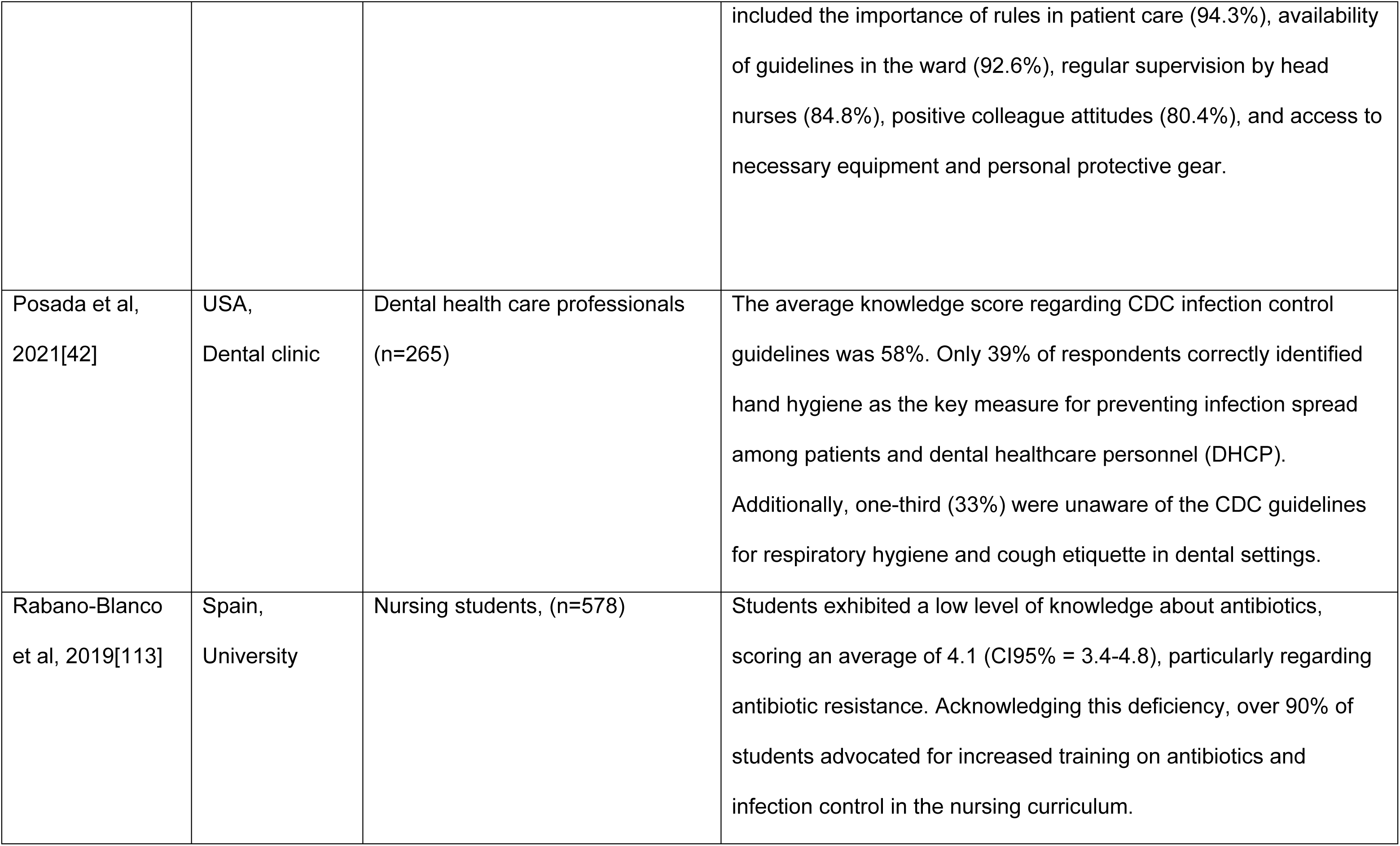
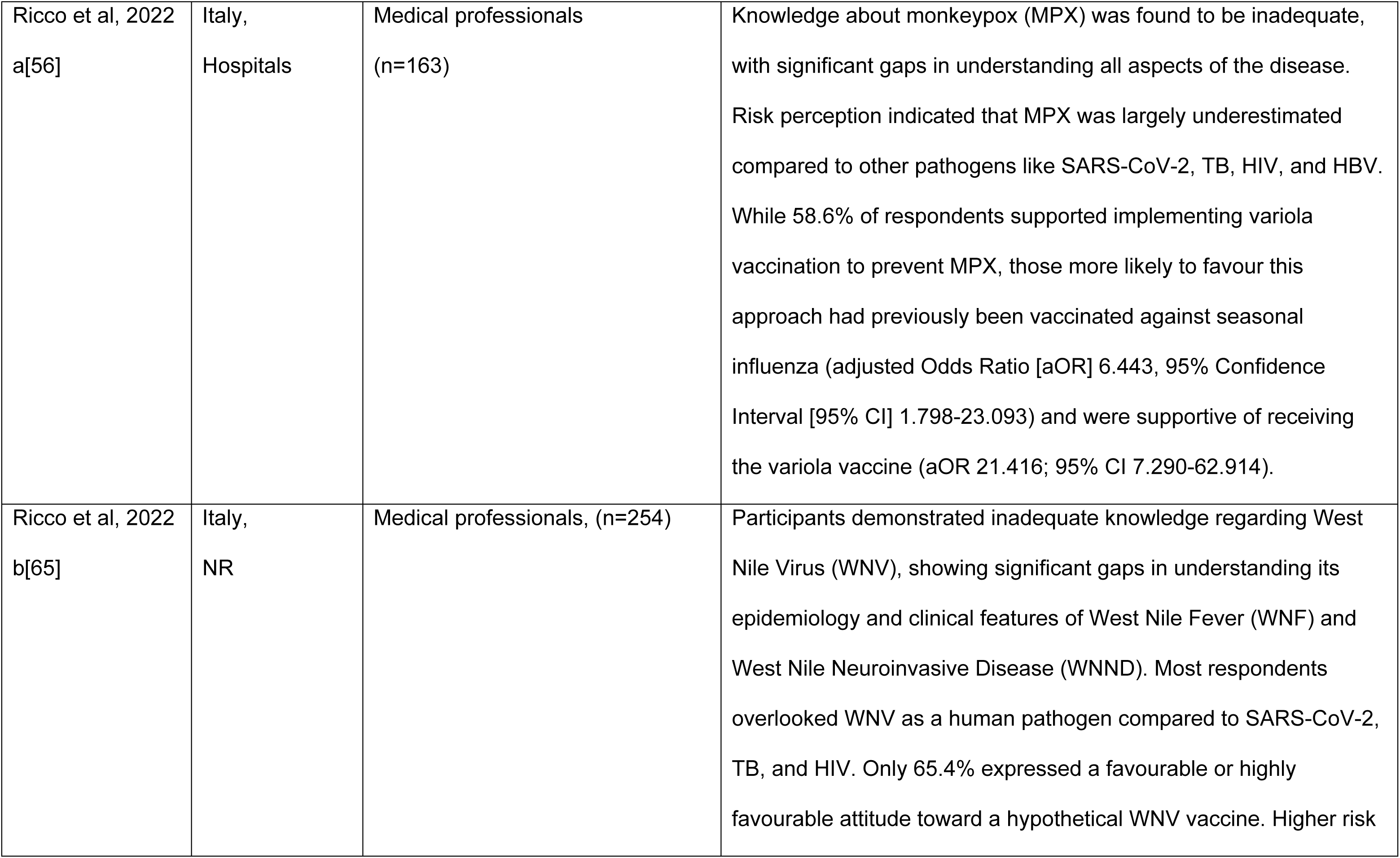
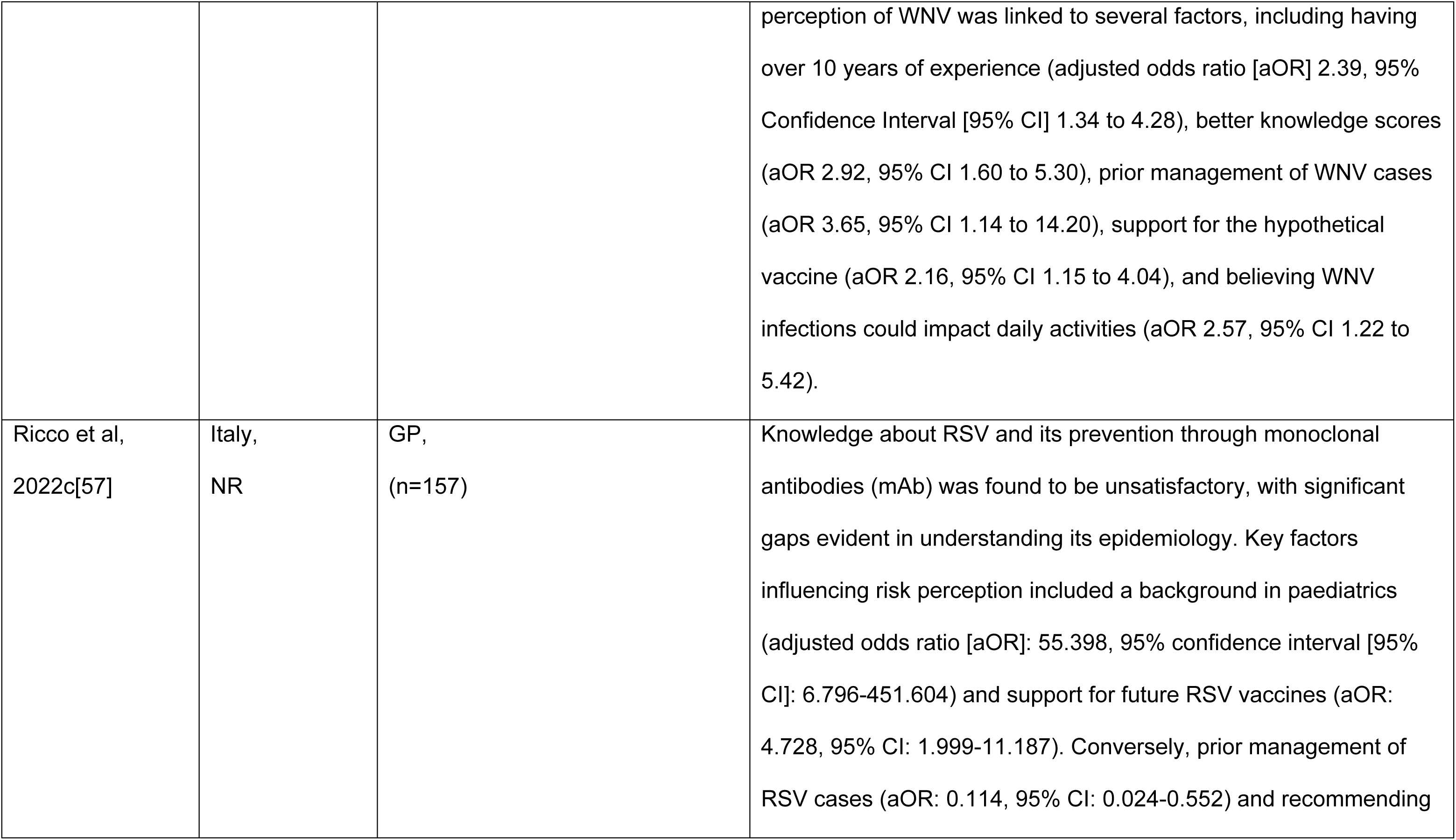
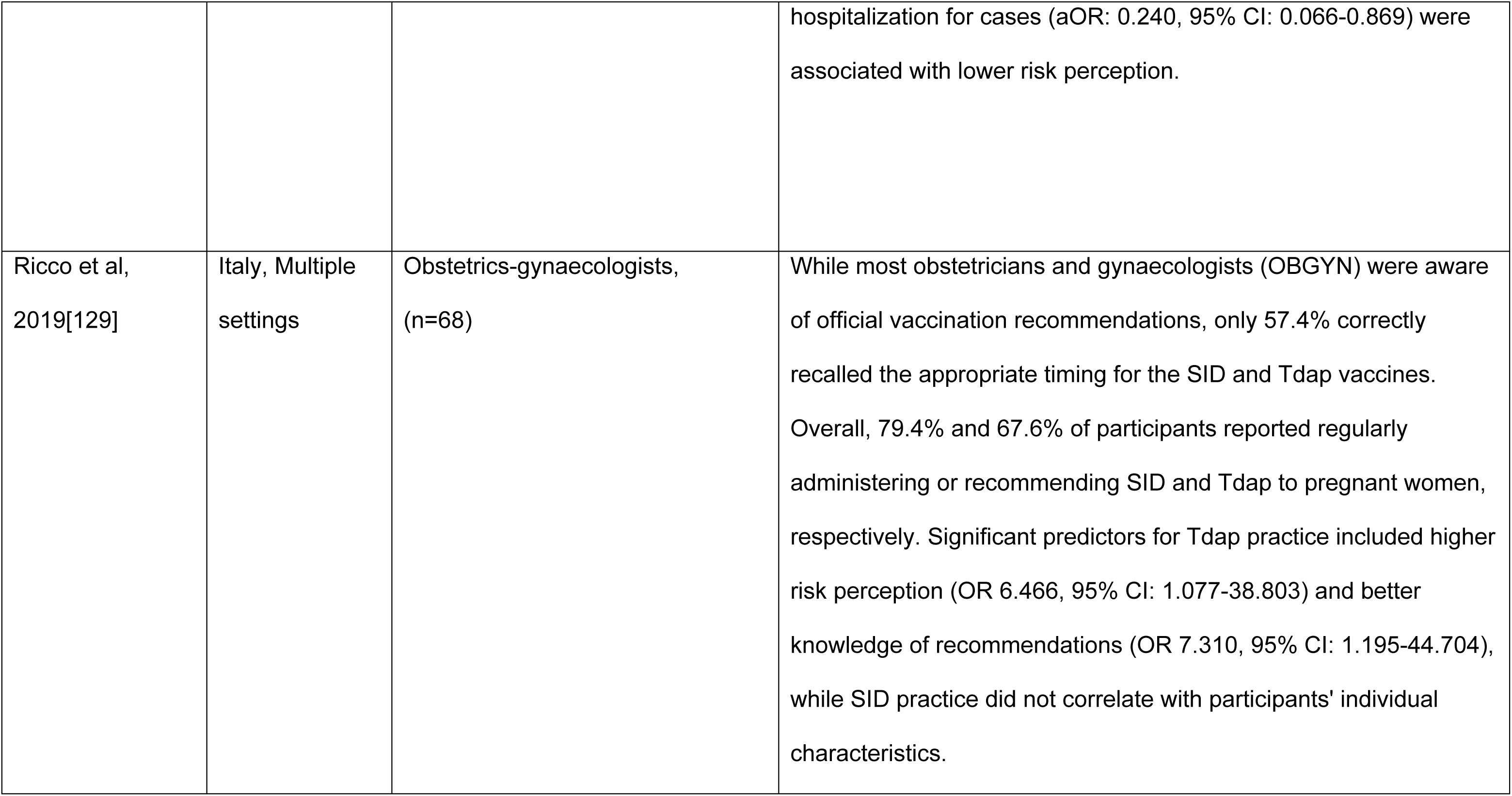
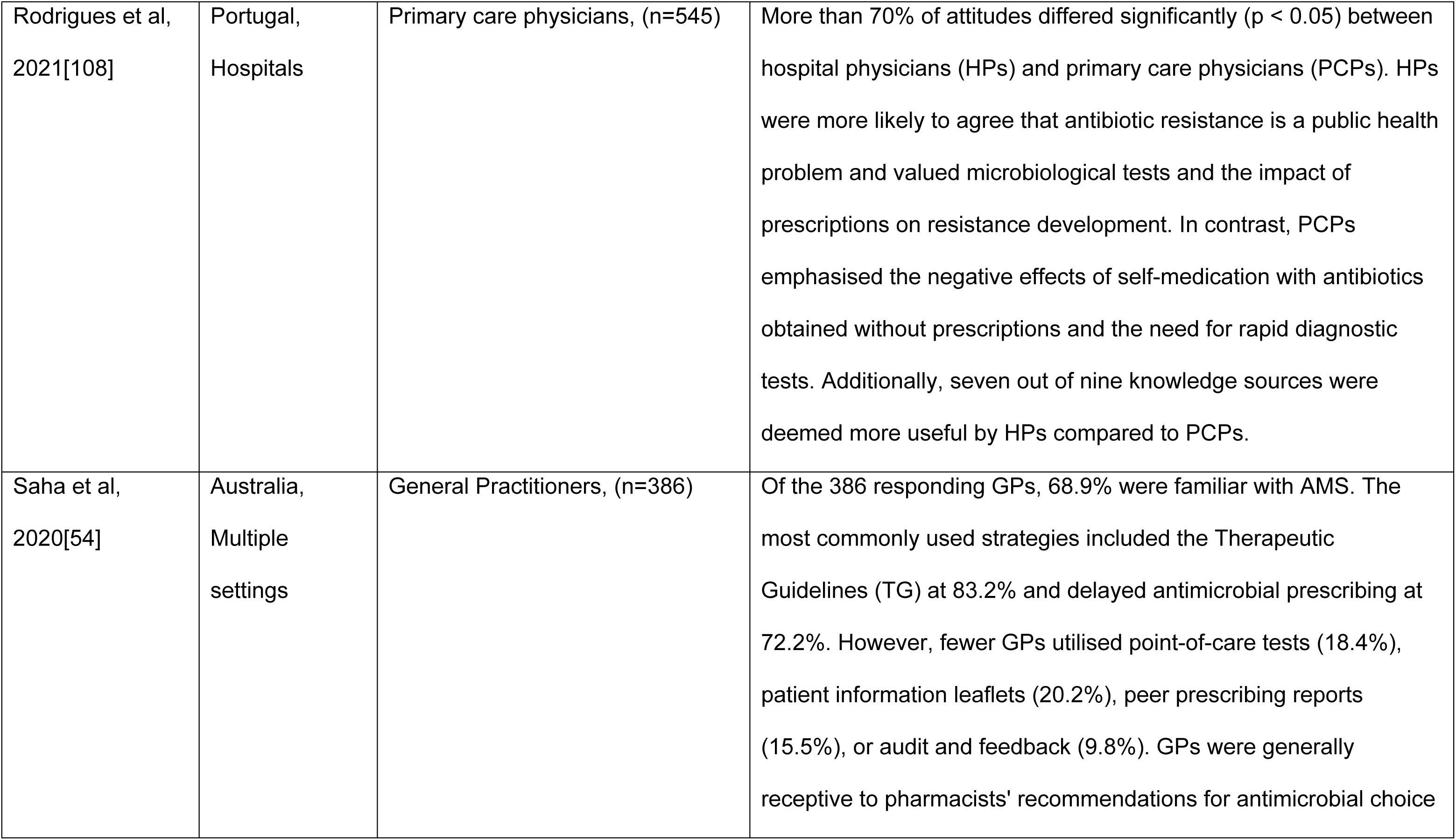
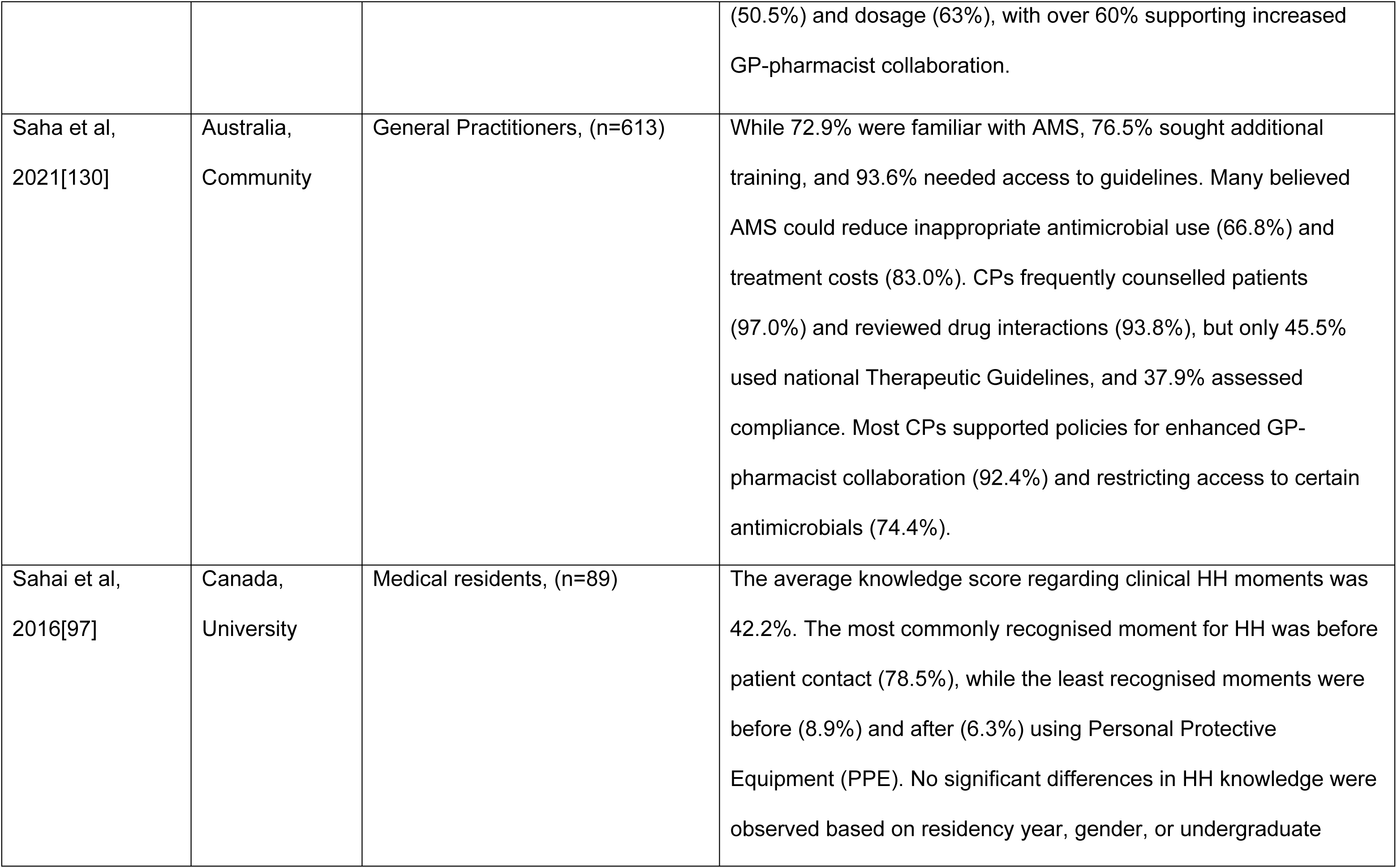
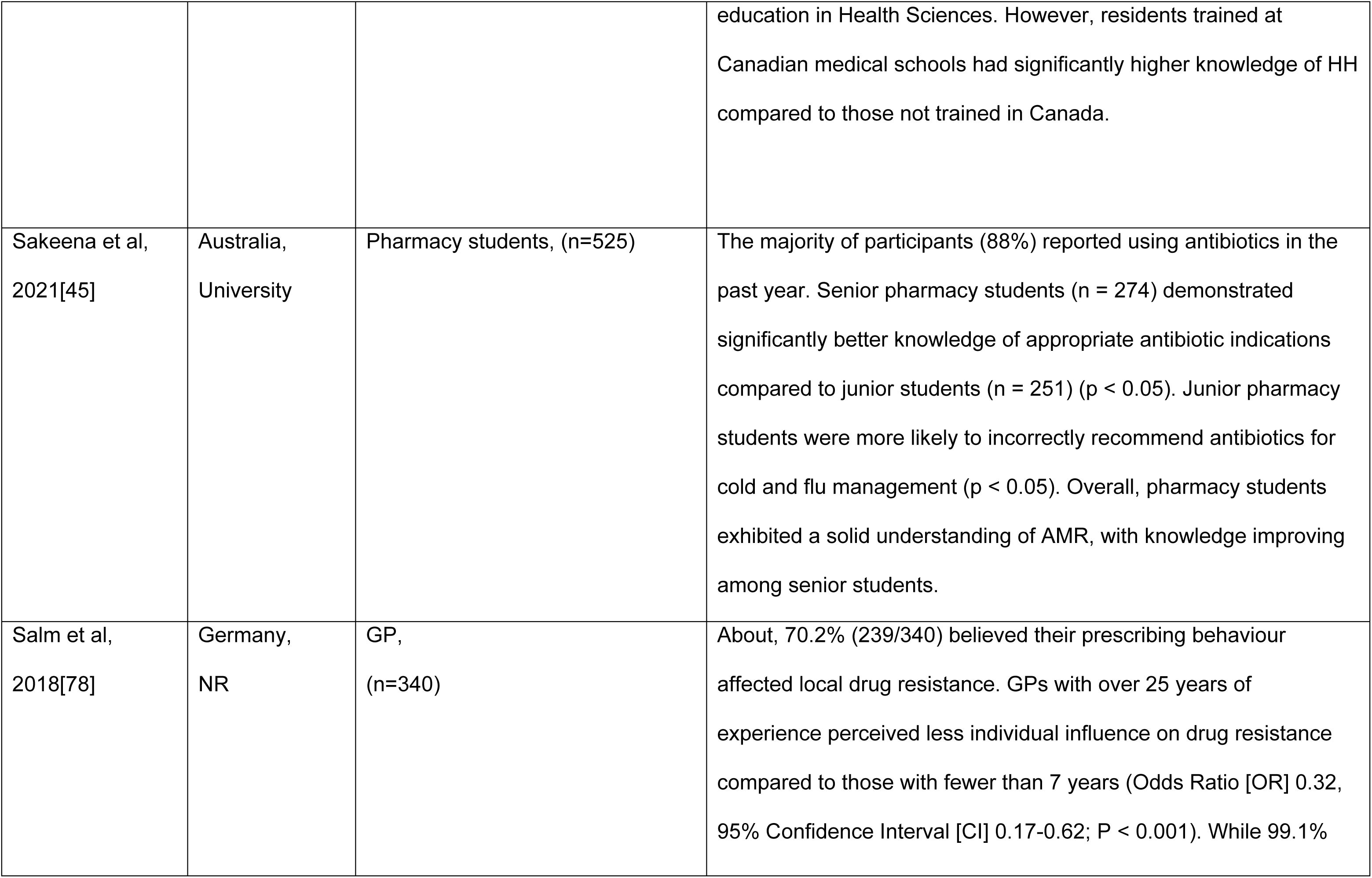
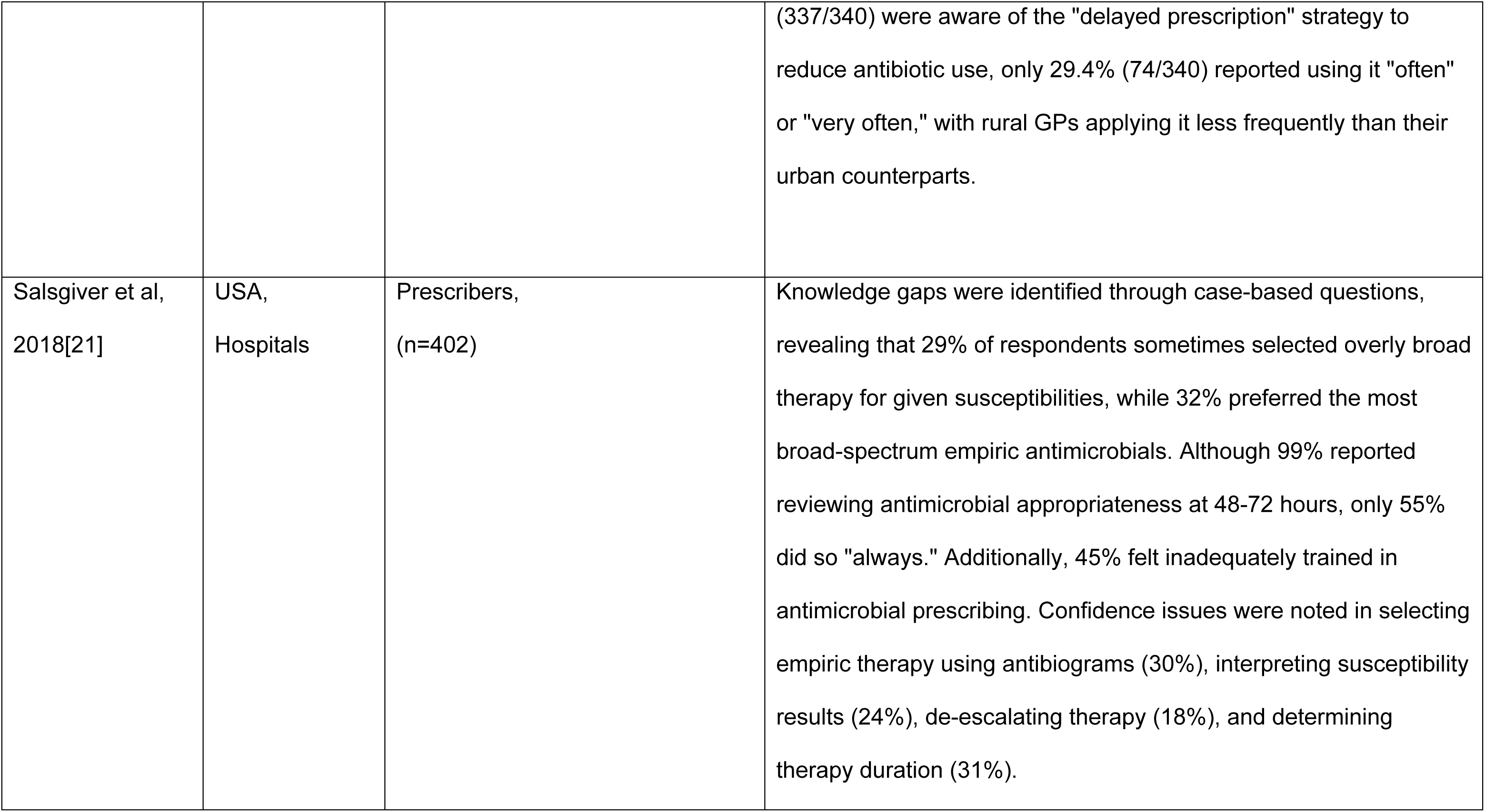
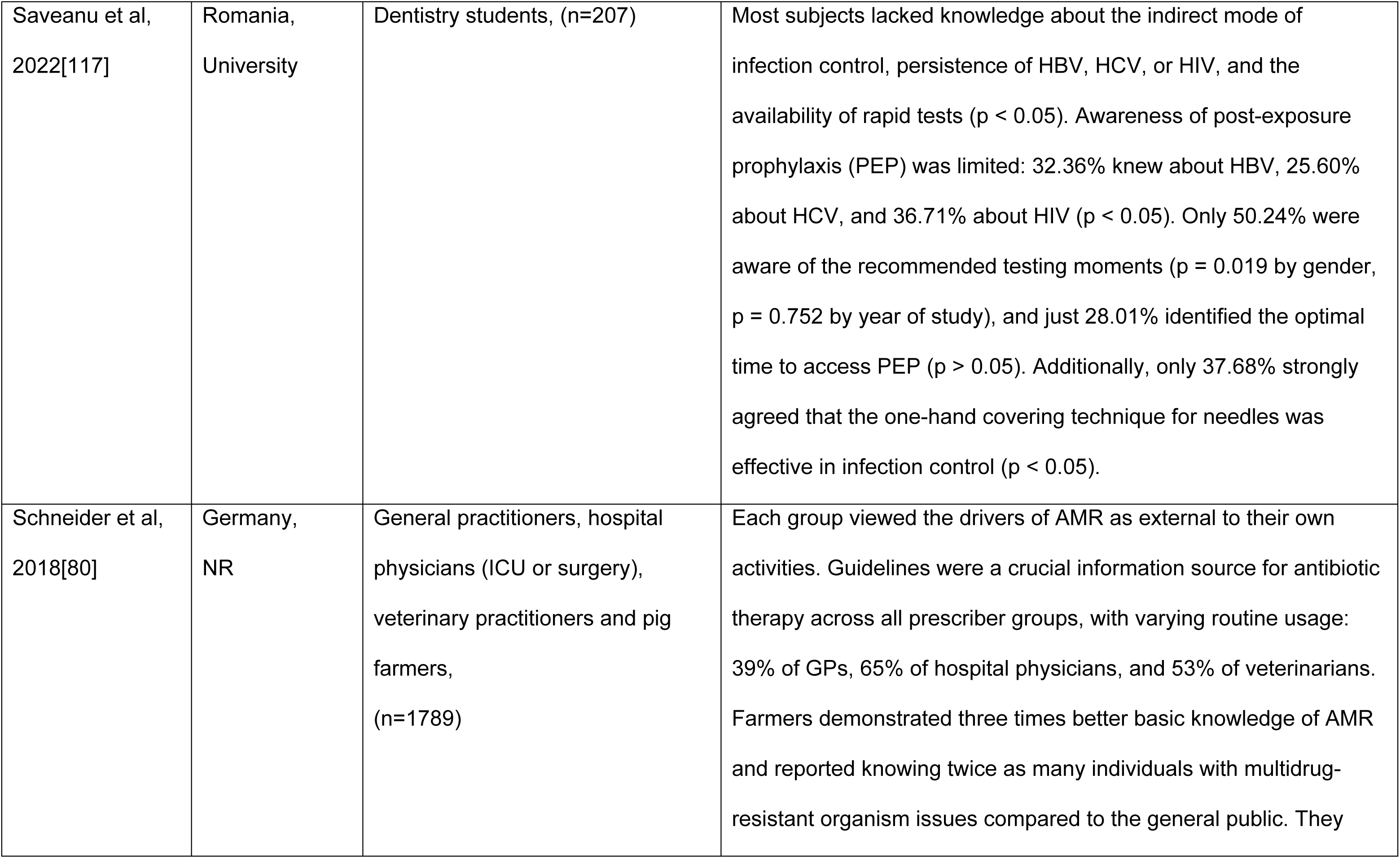
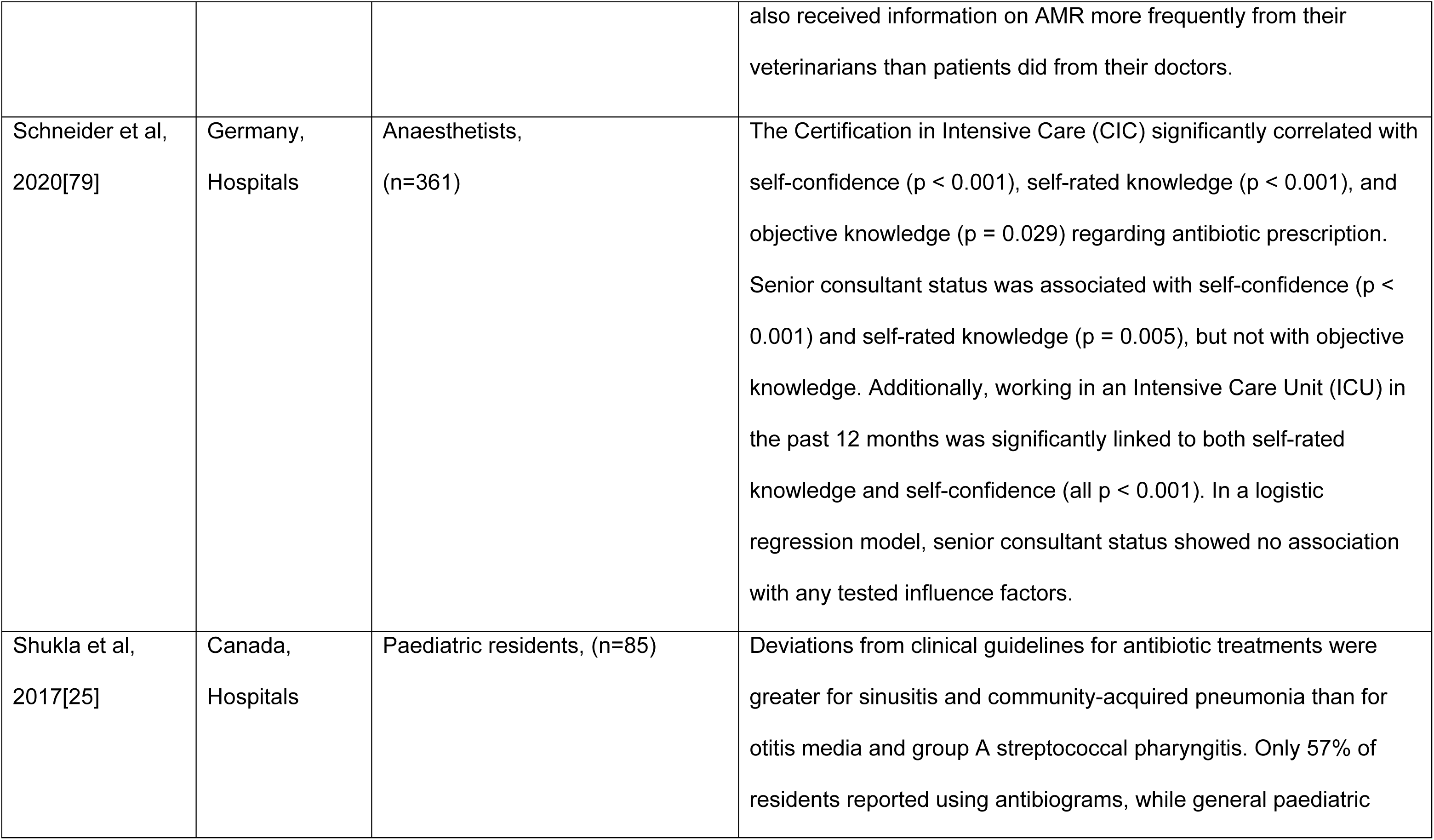
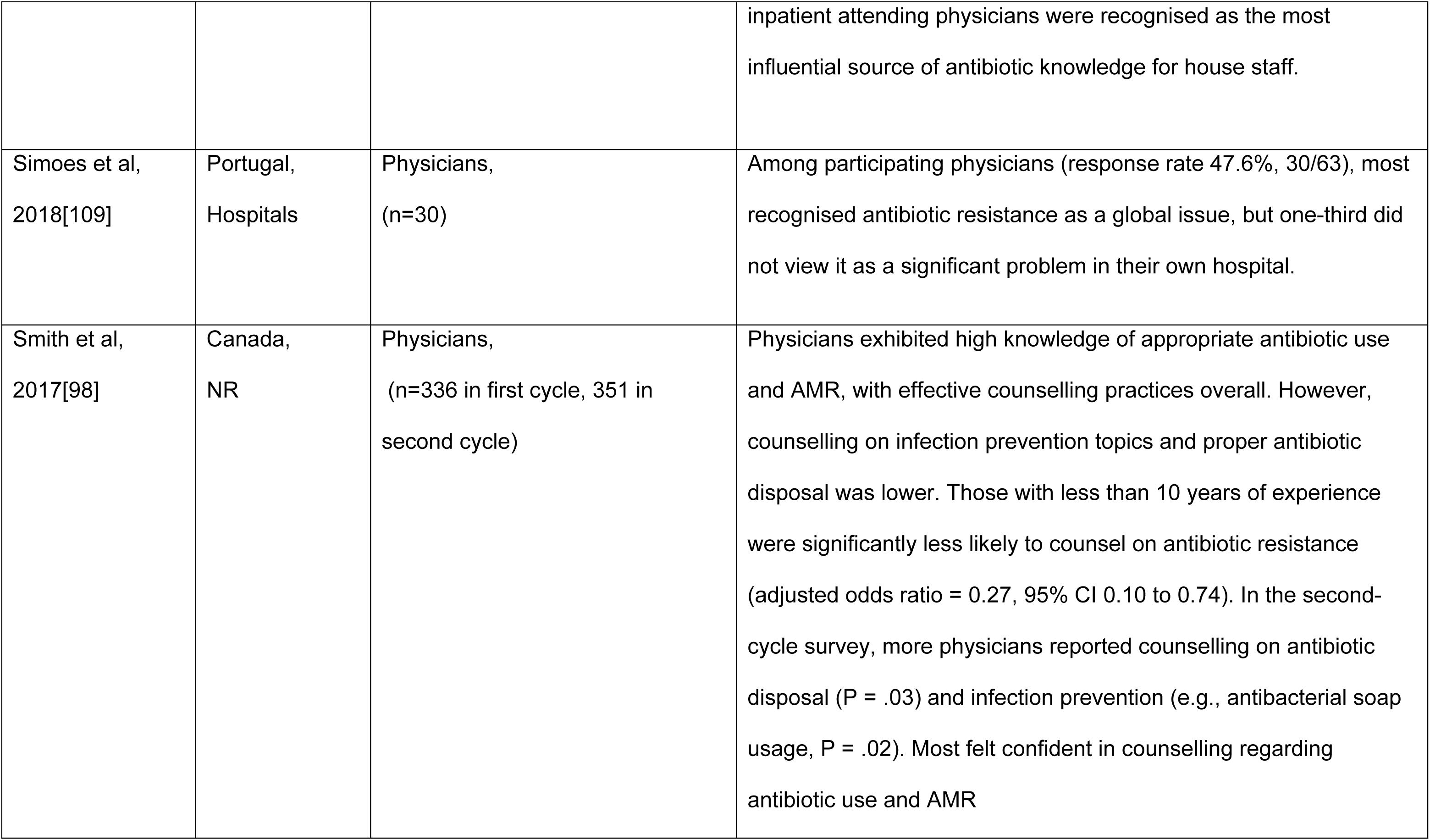
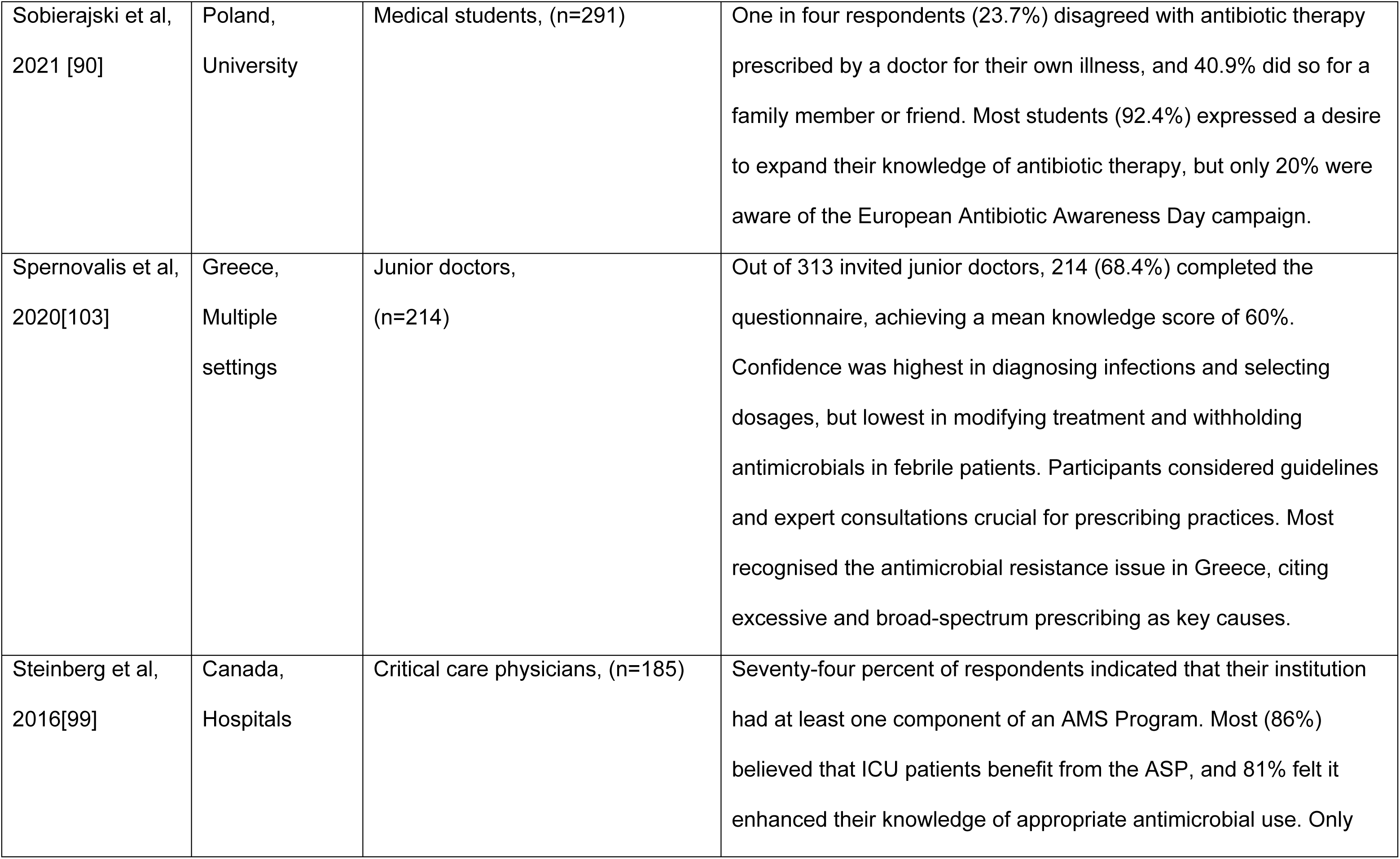
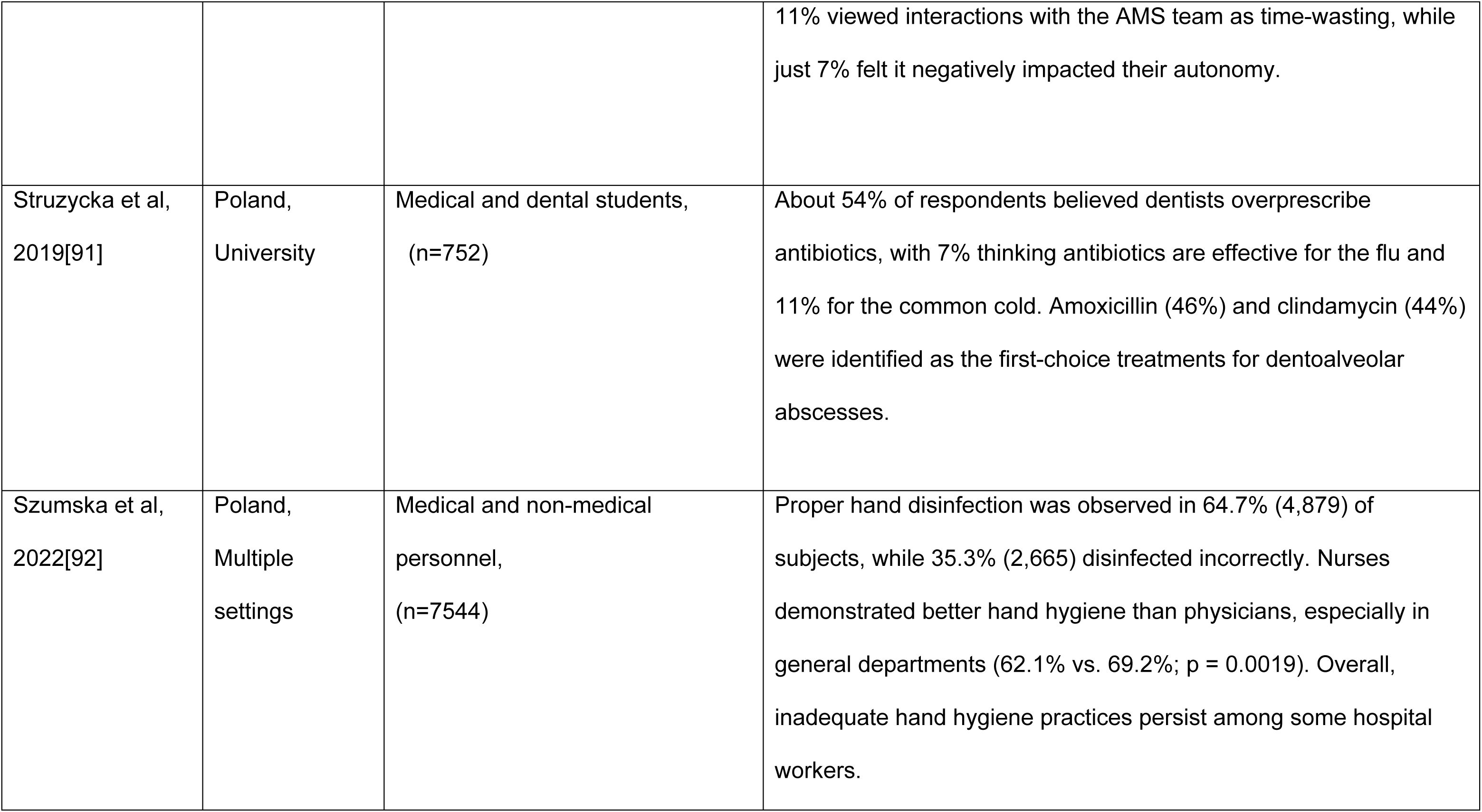
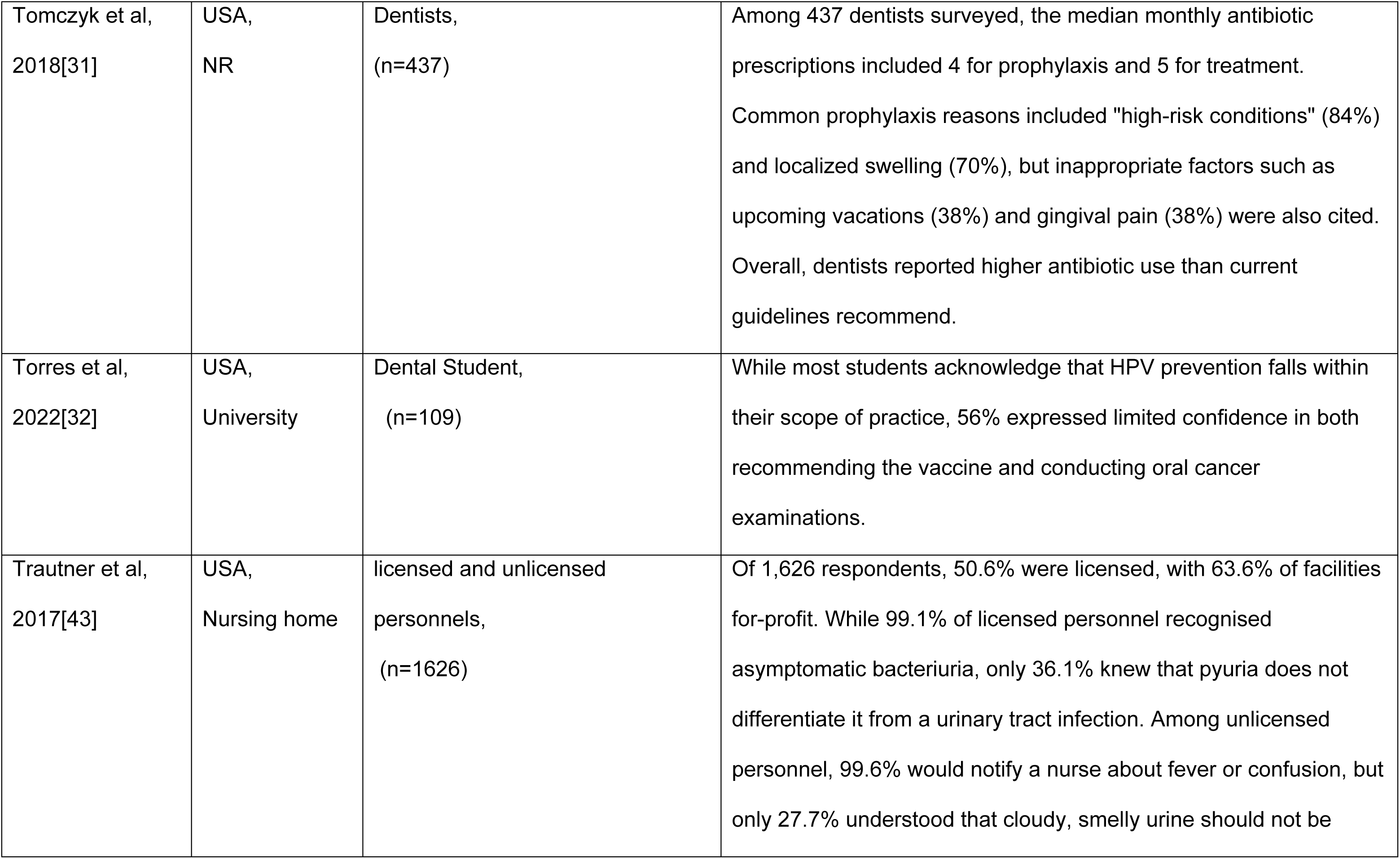
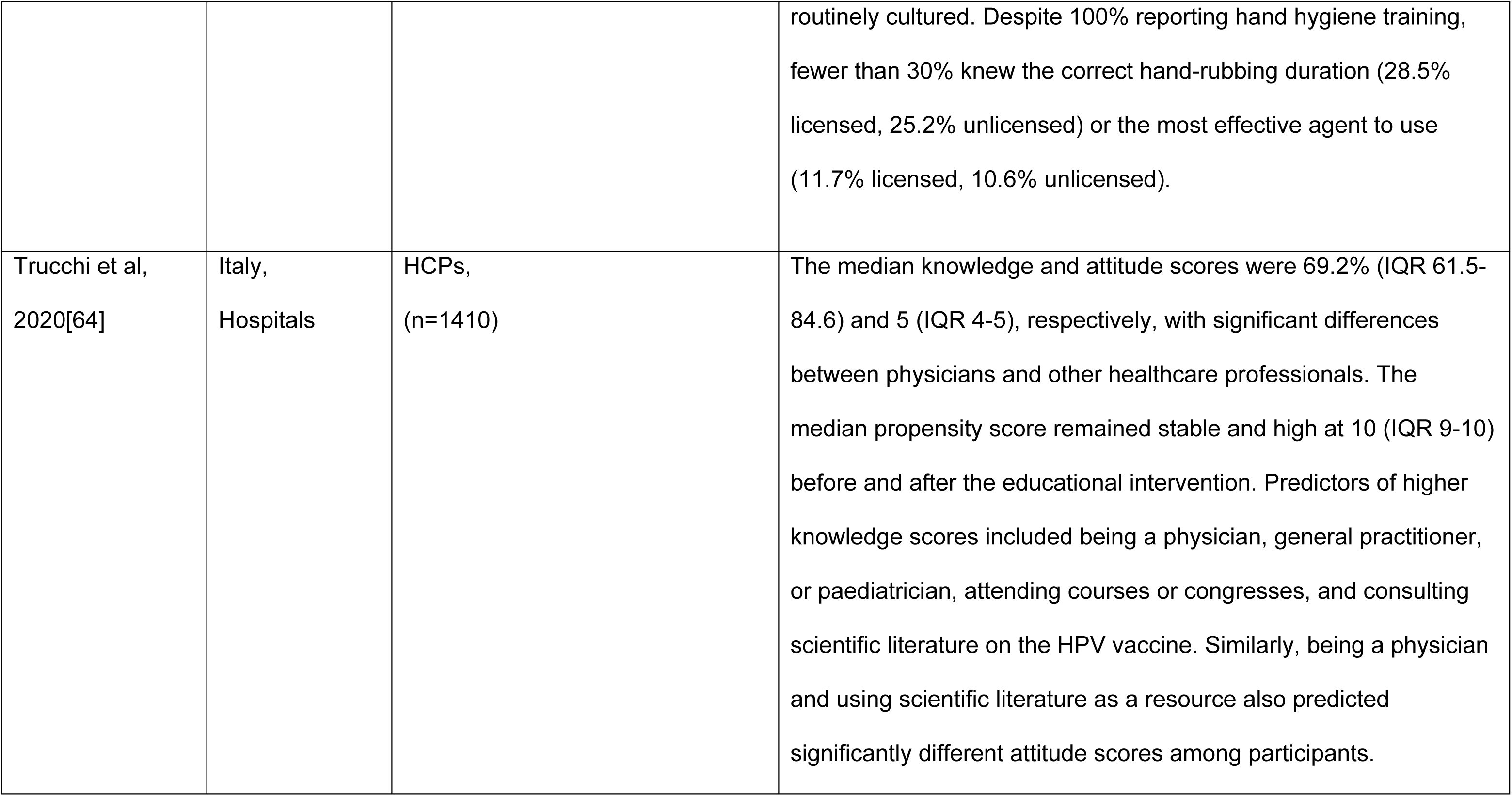
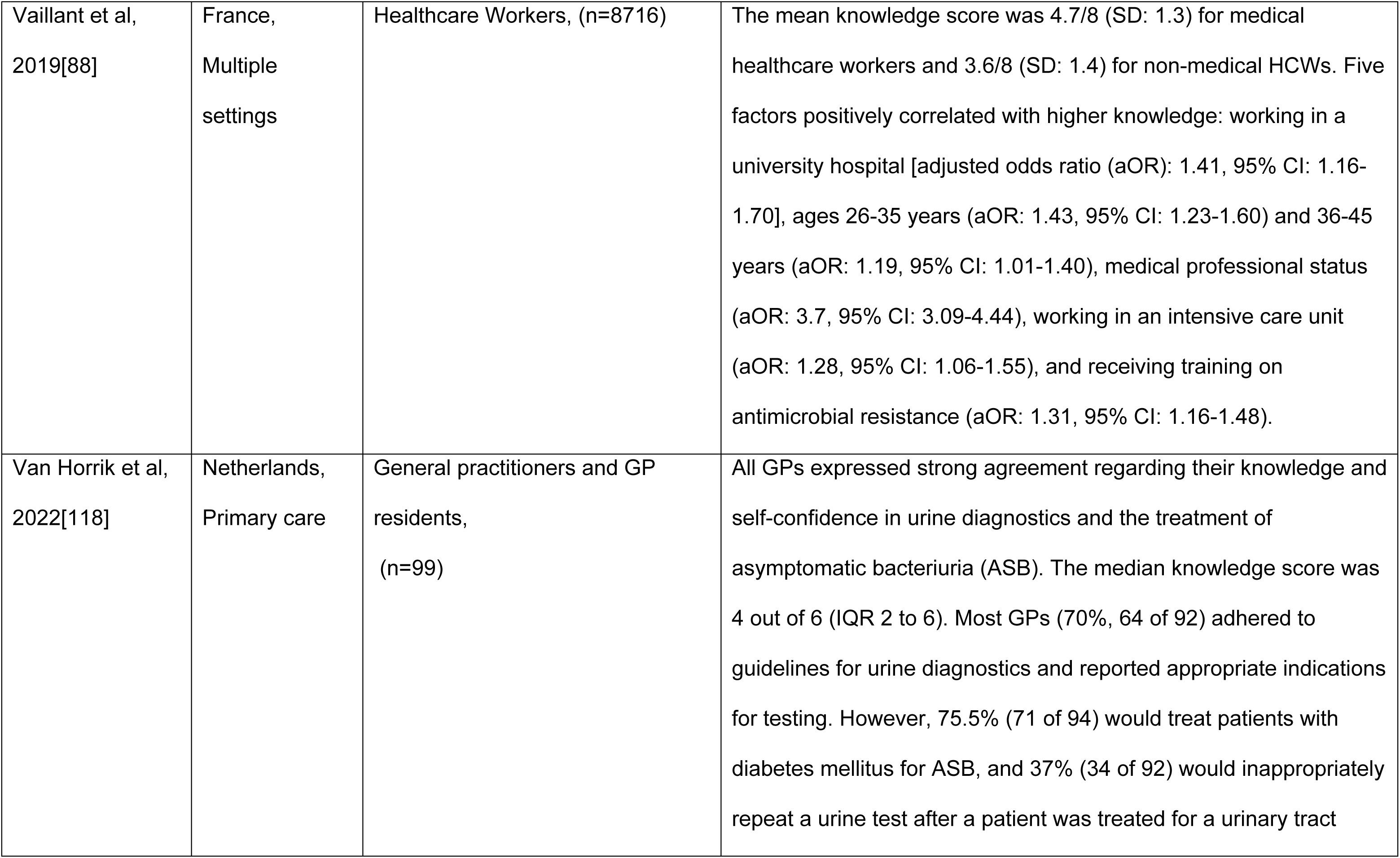
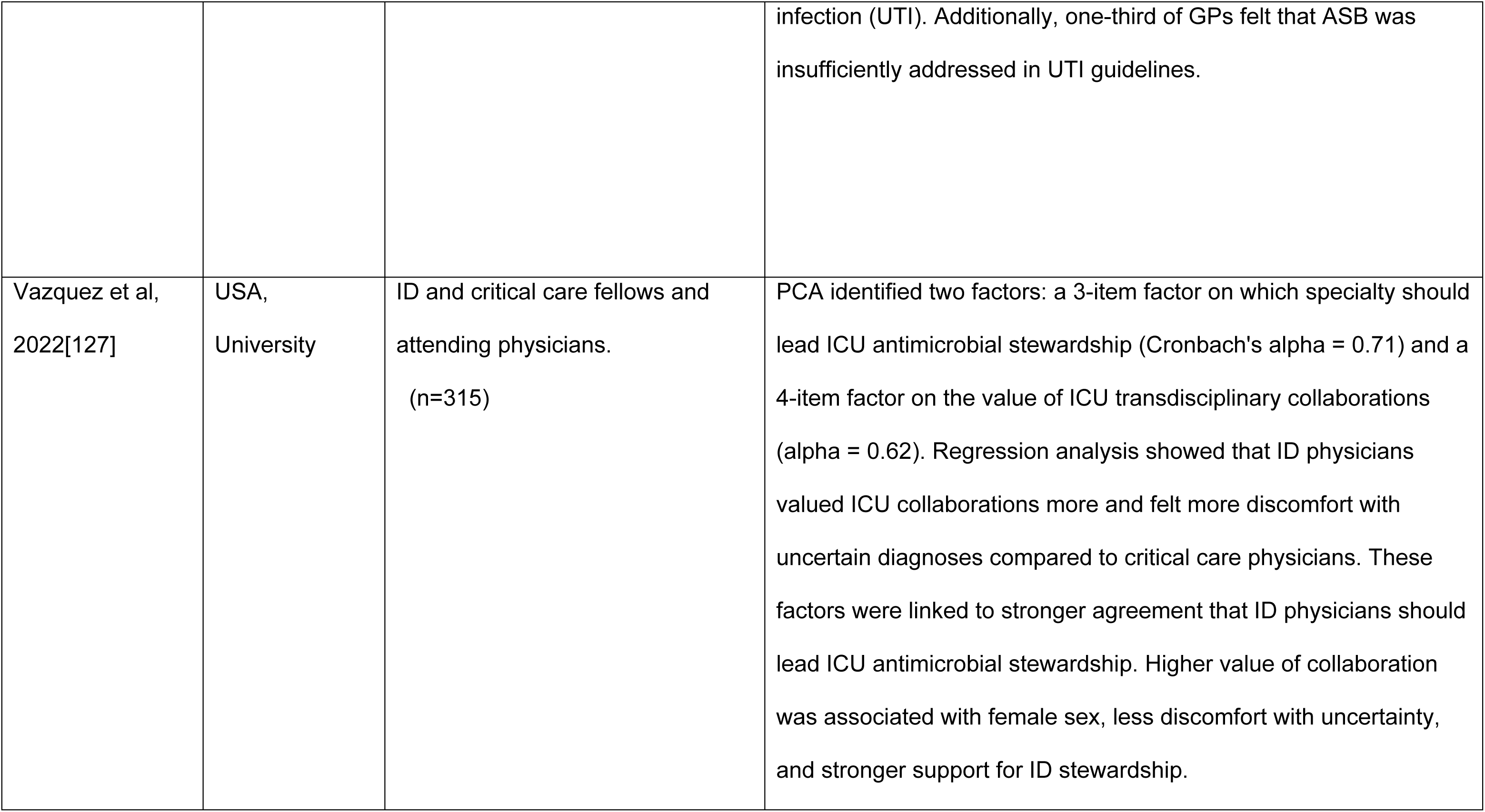
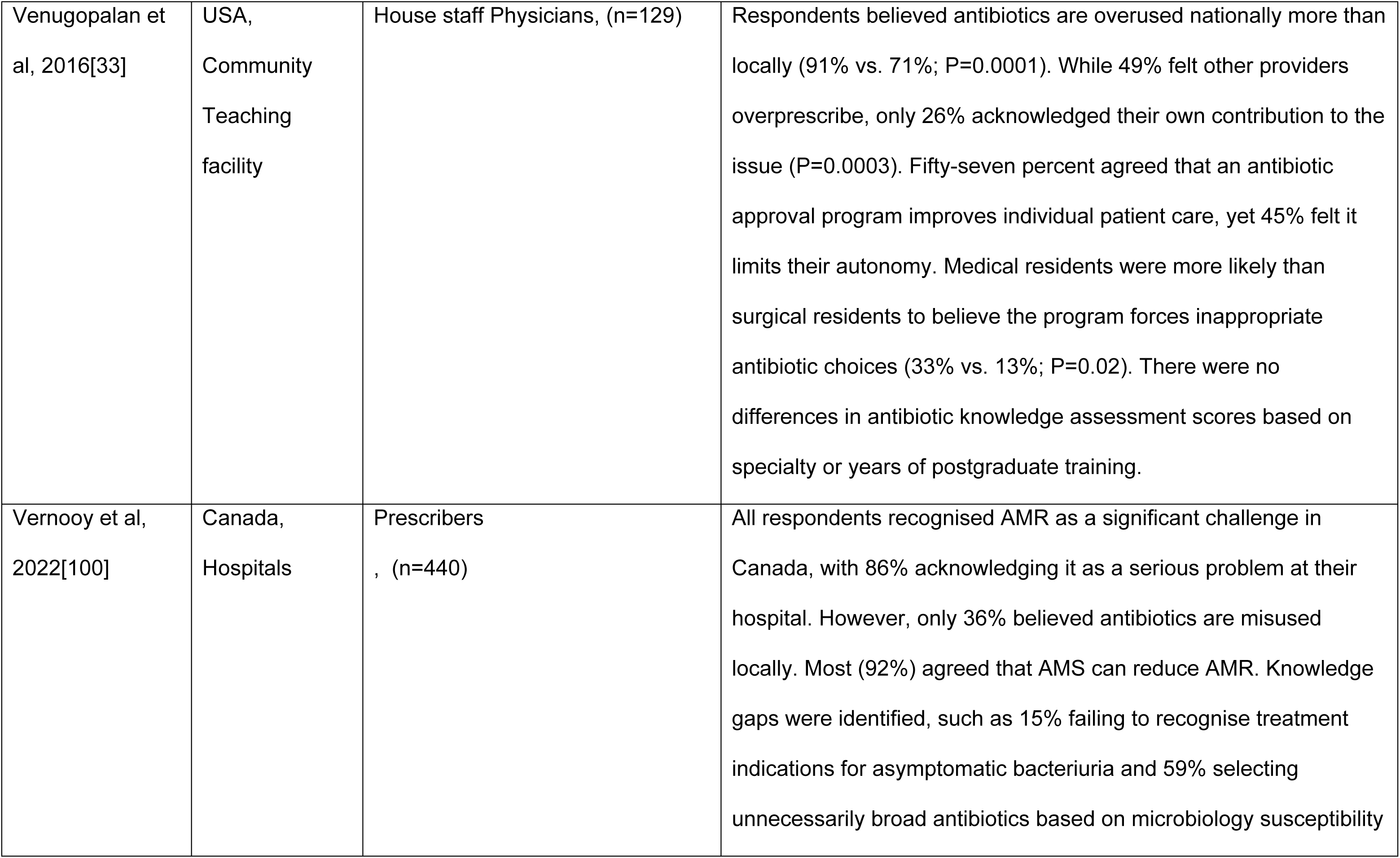
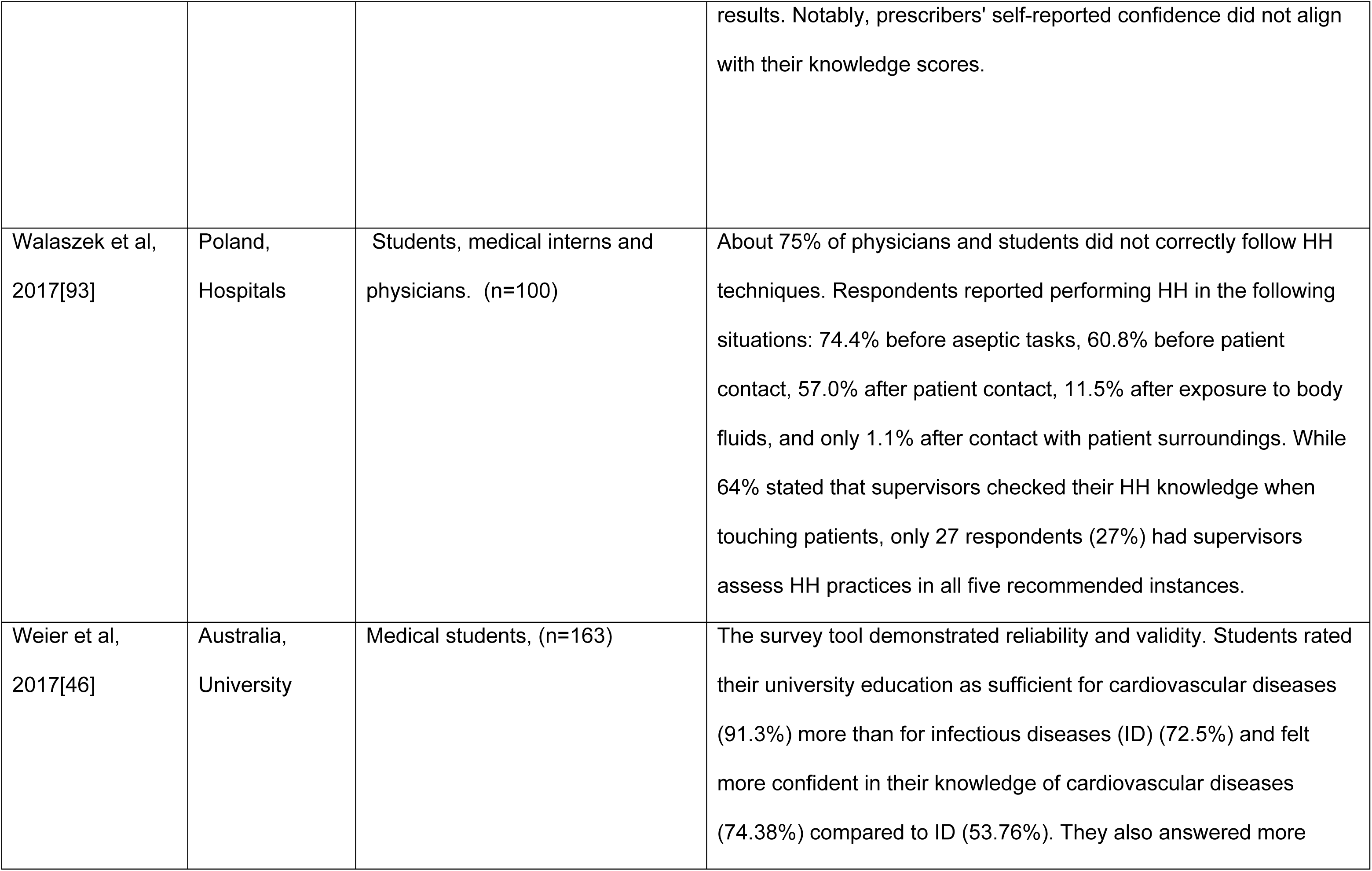
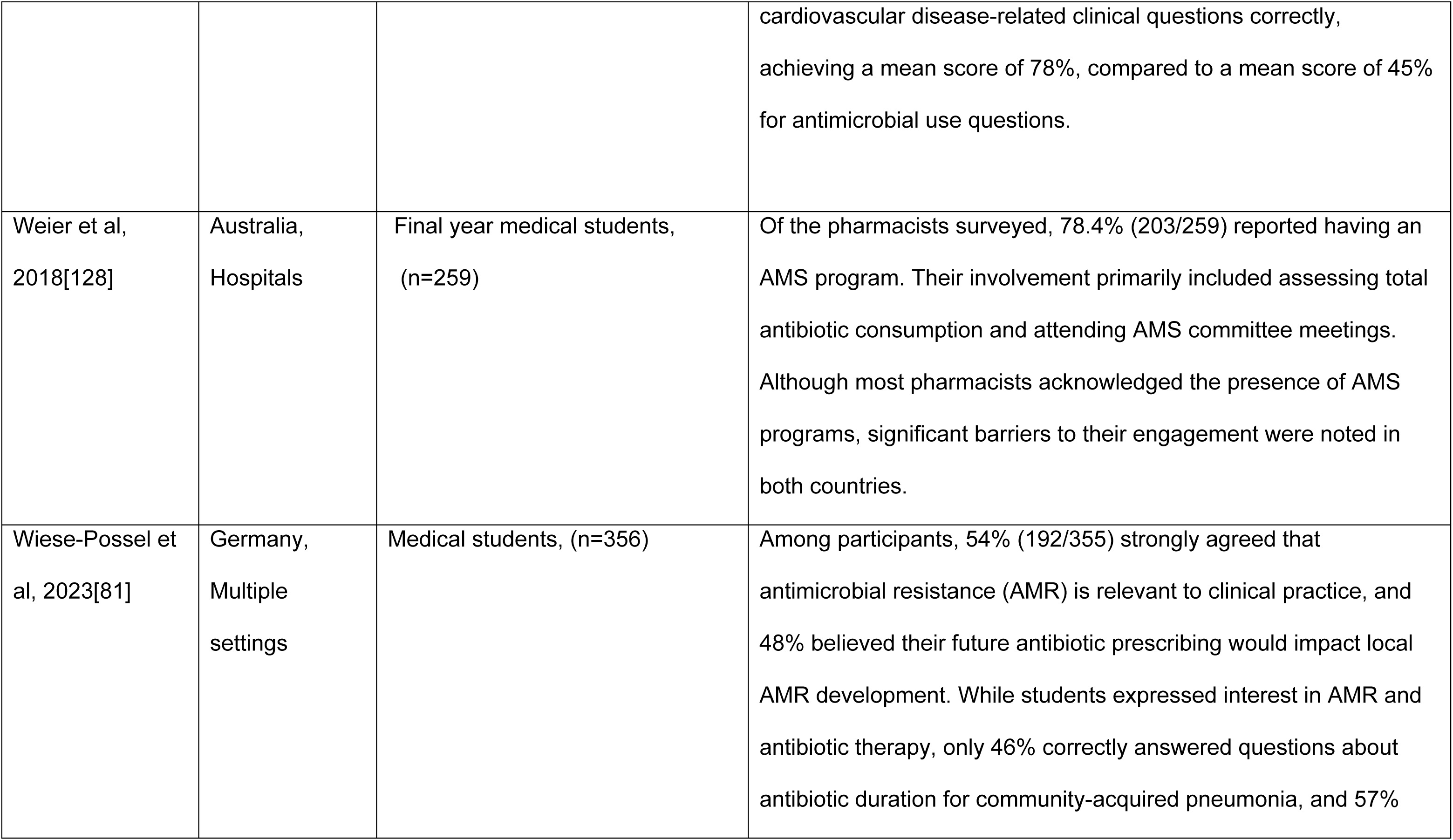
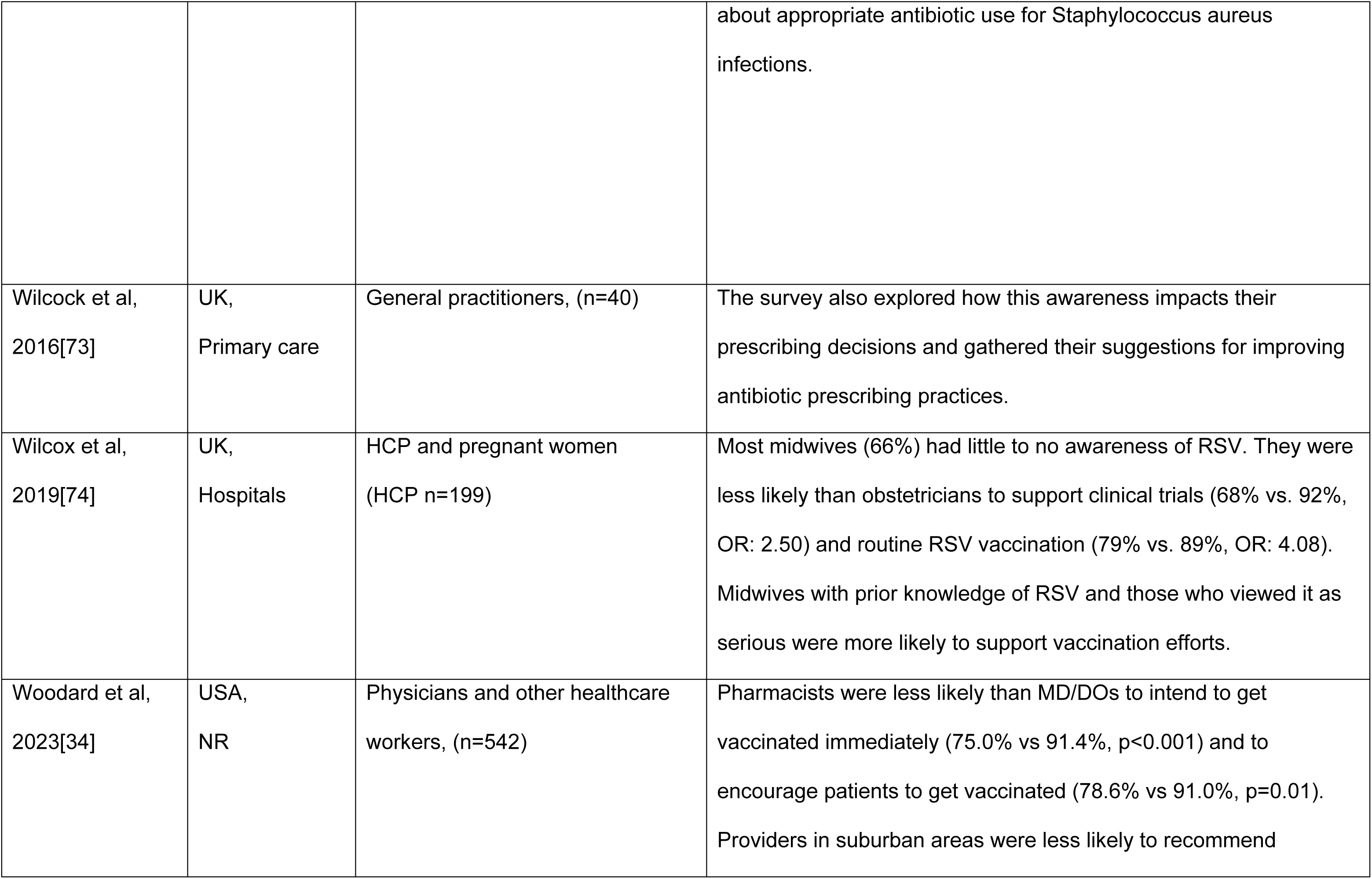
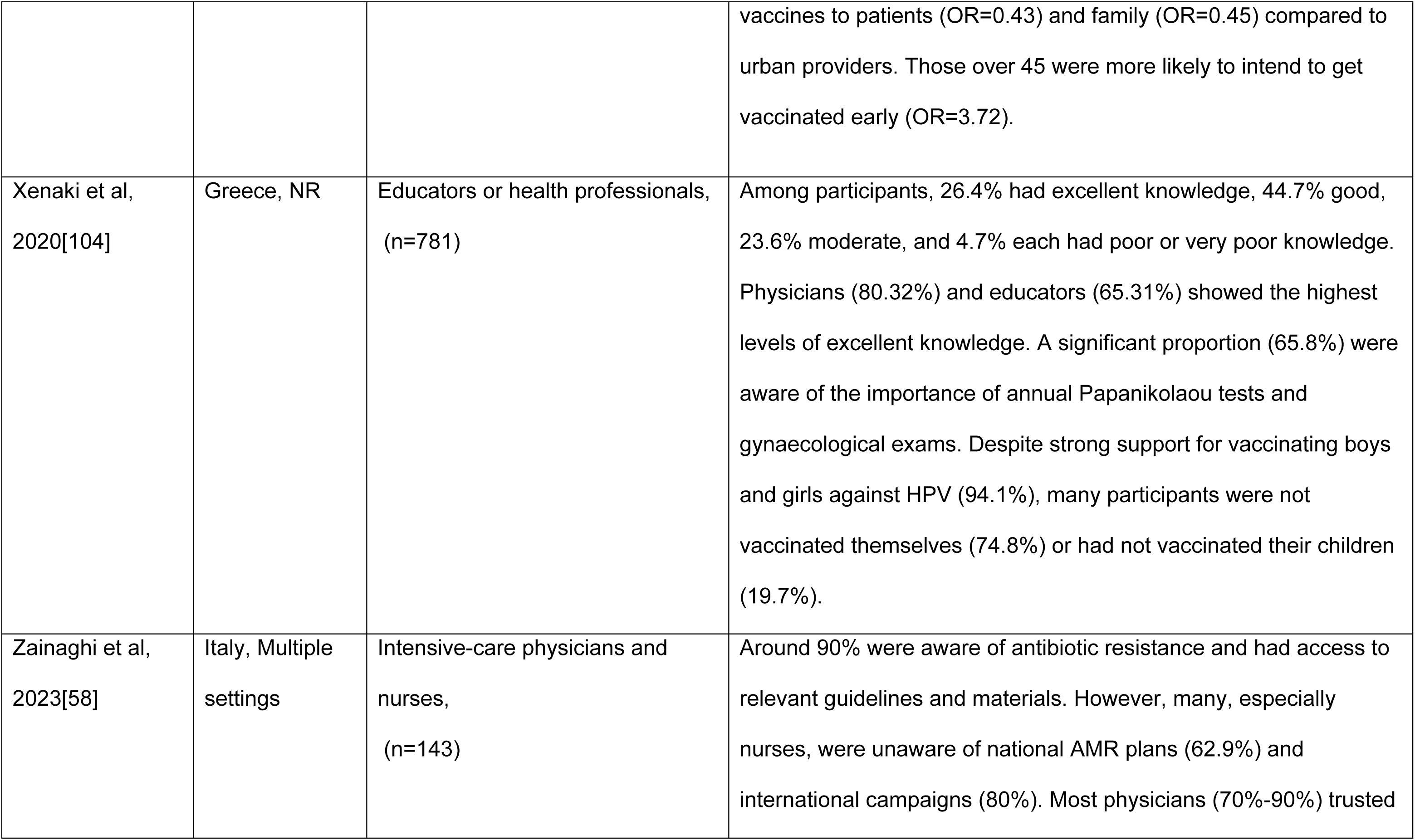
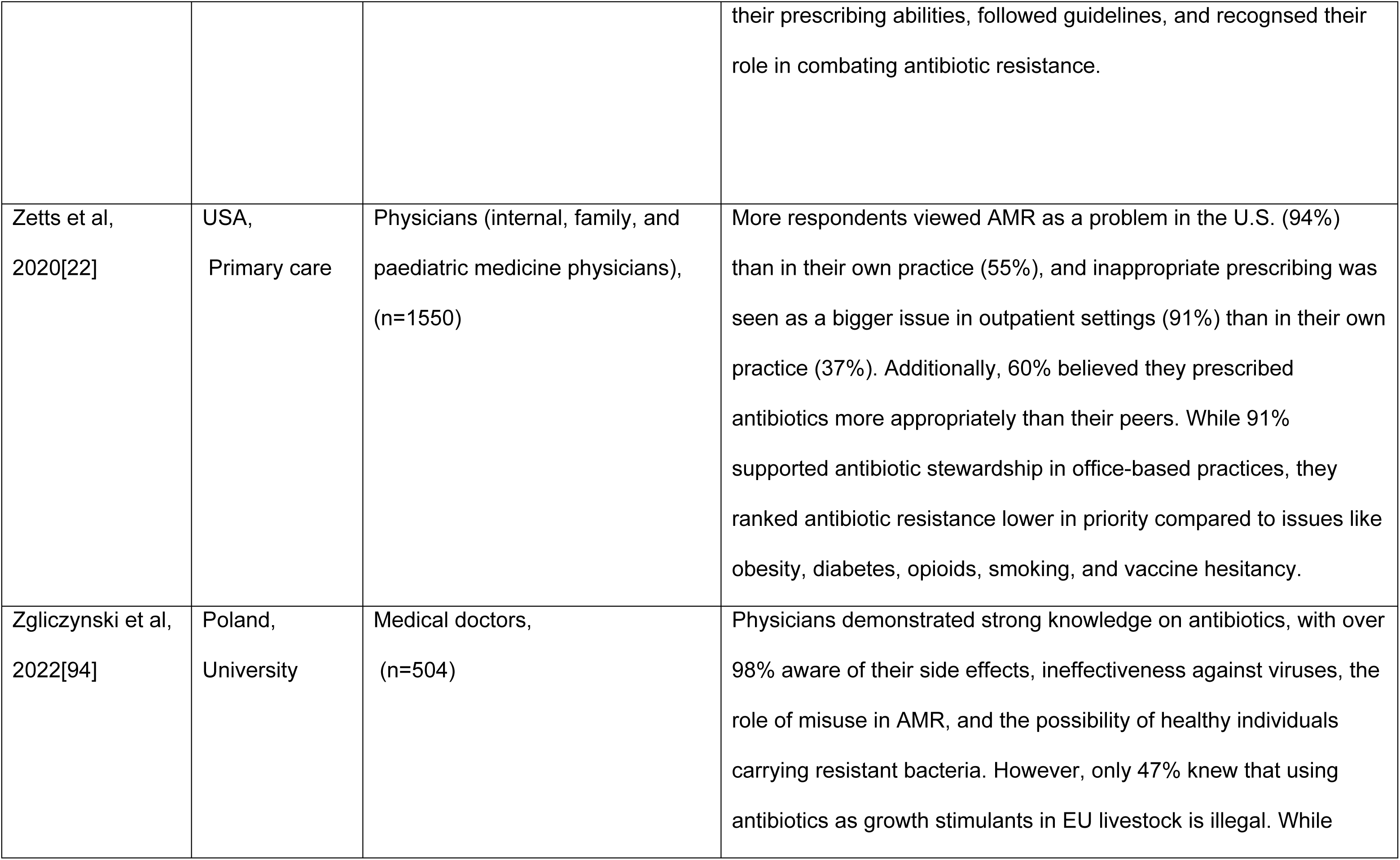
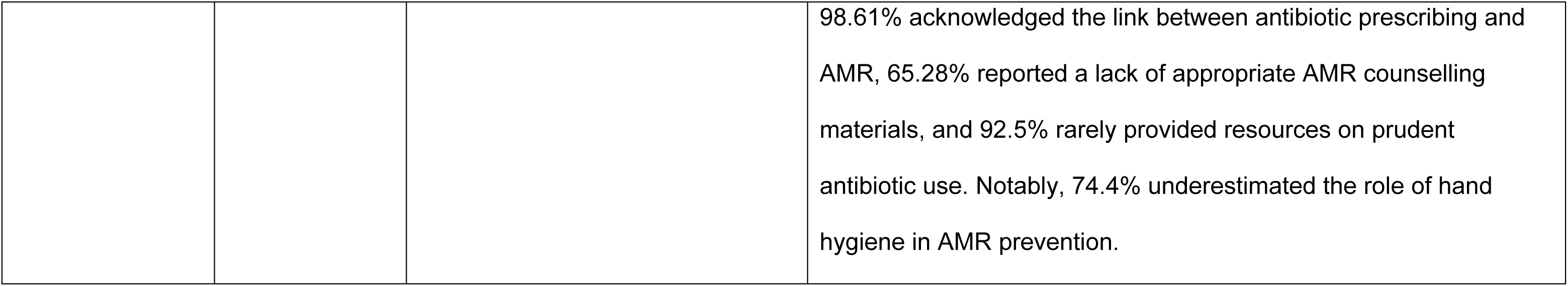
Characteristics of the included studies.

Romania,[117] Netherlands,[118] Latvia,[119] Hungry,[120] Belgium[121] and Georgia.[122] Six[123–128] studies included participants from multiple countries. The sample size of the included study ranged from14 participants [95] to 18,365[123] with a total of 81,710 participants. The included studies covered various categories of healthcare professionals. Some studies focused exclusively on specific groups, such as doctors, including general practitioners, junior doctors, residents, specialists, dentists and veterinarians (n=37);[18, 22, 25, 26, 28, 30, 33, 38, 48, 51, 54, 56, 57, 63, 65, 73, 76, 78, 84–86, 89, 94, 95, 97–99, 103, 108–110, 112, 118, 124, 127, 129, 130] nurses (n=12)[19, 23, 37, 39–41, 49, 75, 115, 119, 120, 131] dentists (n=6);[31, 42, 60, 71, 82] [111] pharmacists (n=2);[44, 61] veterinaries (n=1)[126]. In contrast, some studies combined various categories of healthcare workers such as prescribers, physicians’ assistants, public health professionals, medical researchers, and allied healthcare workers (n=38).[20, 21, 24, 27, 29, 34–36, 50, 53, 55, 58, 59, 64, 66, 67, 72, 74, 77, 79, 80, 87, 88, 92, 93, 100–102, 104, 106, 107, 114, 116, 121–123, 125] 17 studies involved students (medical, dental, nursing, pharmacy and veterinary).[32, 45–47, 52, 62, 68–70, 81, 90, 96, 105, 113, 117, 128]

The majority of participants were recruited from multiple settings, accounting for 20% (n=24)[22, 28–30, 37, 48, 53, 54, 63, 72, 76, 79, 80, 85, 87, 92, 94, 103, 109–111, 115, 122, 125] of the total included studies, with five studies conducted on a national scale.[59, 60, 66, 123, 132] This was followed by a diverse range of hospitals (n=21),[21, 25–27, 41, 51, 55, 58, 64, 74, 77, 86, 93, 95, 99, 101, 102, 108, 119–121] including both speciality and general hospitals. An equal number of studies (n=21) recruited participants from teaching institutions, such as dental schools, nursing schools, and pharmacy universities.[32, 33, 45–47, 52, 62, 68–70, 90, 91, 96, 100, 104–107, 113, 117, 126] Ten studies drew participants from community-based healthcare systems,[24, 35, 39, 44, 61, 88, 114, 116, 130, 131] while two studies each recruited from nursing homes,[43, 75] primary care centres[112, 118], or tertiary health centres.[23, 50] One study specifically mentioned a rural clinic as the source of its sample.[20] Additionally, six studies were conducted among conference or meeting attendees, or recruited participants through a professional network.[19, 36, 40, 67, 73, 89]

The majority of the questionnaires were delivered via a web-based (n=70) platform[18–27, 29, 31, 34–38, 41, 42, 45–49, 51–53, 55–59, 63–68, 70, 72, 81, 84, 86, 95, 98–100, 103, 104, 106, 107, 110–112, 114, 115, 118, 119, 121, 123–129, 131, 132], whereas, 15 were reported as being paper-based,[33, 39, 54, 69, 71, 76, 78, 79, 89, 94, 108, 109, 116, 120] Five studies offered both online- and paper options;[28, 30, 32, 40, 130] one study provided online- and telephone-options [60]and one study was solely telephone-based.[85] Two studies reported using multiple modes of delivery including online-, paper-, and telephone.[61, 80] The remaining studies did not specify the mode of delivery.

### 3.3. Survey findings and instrument items

Figure 2 depicts the number of studies measuring each component of KAP. The included studies reported 106 unique questionnaires to capture KAP; six [58, 59, 66, 94, 121, 123]studies adopted either the original or a modified version of the ECDC 2019 questionnaire. The instrument used in Hamidi et al,[37] was adopted from Merili’ et al’s[41] work on AMS. Similarly, an adapted version of the Boudet et al [82] questionnaire was used in Loume et al.[111] Knowledge and perceived knowledge were measured using a range of questionnaires. Forty [20, 21, 23, 33, 37, 39, 47, 53, 56–59, 63, 64, 66, 69, 70, 75–77, 85, 86, 89, 90, 94, 100, 104–107, 113, 117, 118, 120, 121, 123, 125, 126, 129]studies measured objective knowledge using a choice from yes/no, true/false options, and multiple-choice questions whereas 41 reported use of agree/disagreement statement or Likert scale.[18, 19, 22, 25–28, 32, 35, 36, 40–42, 45, 46, 49, 52, 54, 62, 74, 78–81, 84, 87, 88, 91–93, 97, 99, 101, 103, 108, 109, 115, 116, 118, 128, 130] Almost all studies which have measured attitude either used agree/disagreement statement or Likert scales. The practice questions included Likert scales, multiple-choice options, yes/no responses, and closed questions.

**Figure 2:**
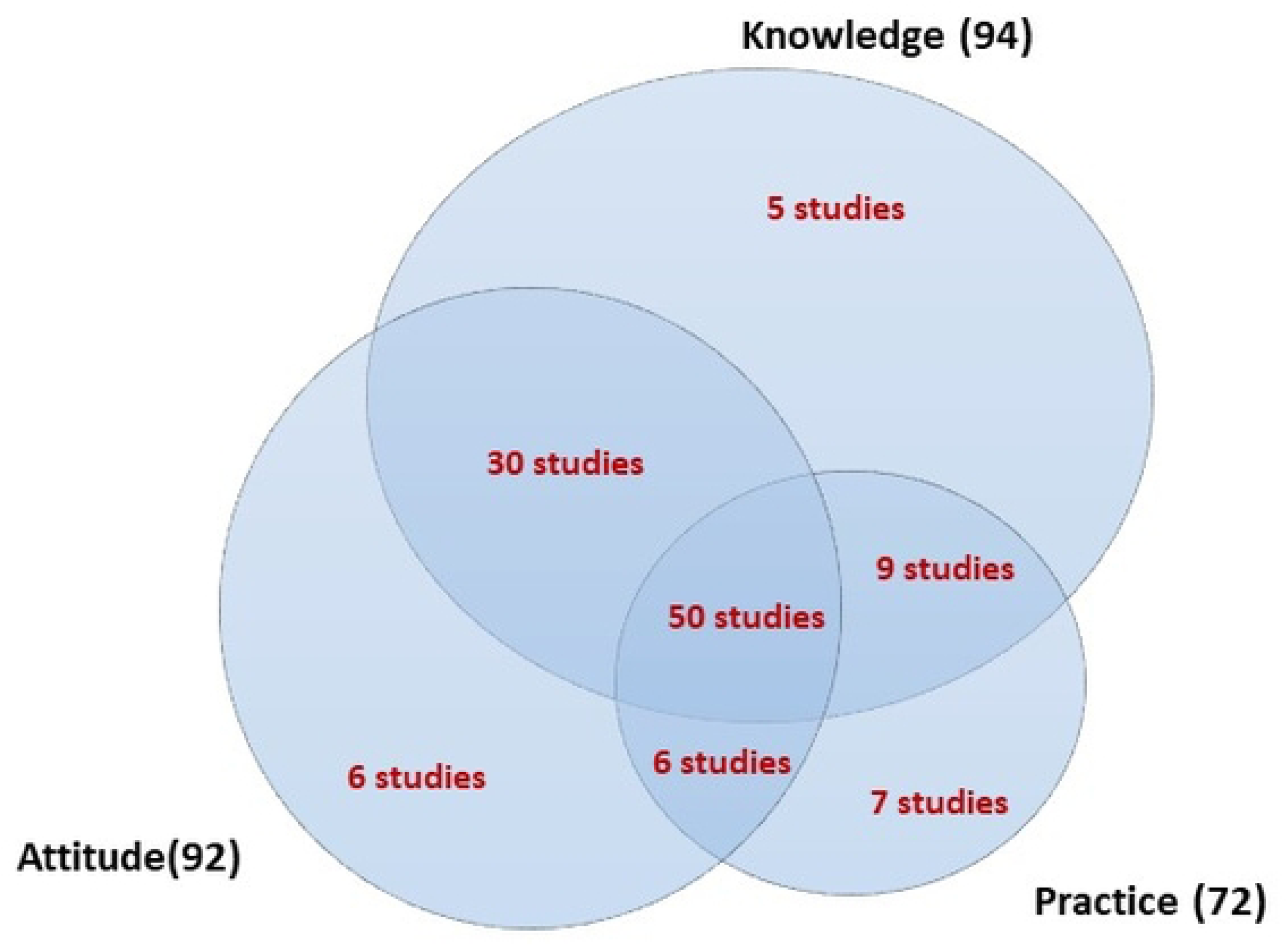
Mapping of the included studies.

The number of items among the questionnaire ranged from 12 questions[23] to 99 questions[34] and the response rate ranged from— about 5%[81] among medical students to 100%[91] among a combined sample of medical and dental students.

### 3.4. Risk of bias

The methodological quality of the included studies is summarised in Figure 3 and Appendix 2. When examining the overall scores, 64 out of 113 studies (57%) were categorised as having a ‘moderate risk of bias,’ with scores ranging from 4 to 6, while the remaining 49 studies (43%) were deemed to have a low risk of bias. Less than half of the studies (29/113) provided a comparison of the target population to the national population on relevant variables (item 1). On the other hand, 67 out of 113 studies justified the sample selection approach (item 3). A large majority (104/113) justified their sample frame (item 2), and 31 out of 113 studies reported a response rate greater than 60% (item 4). These findings suggest that most of the studies have good external validity.

**Figure 3.**
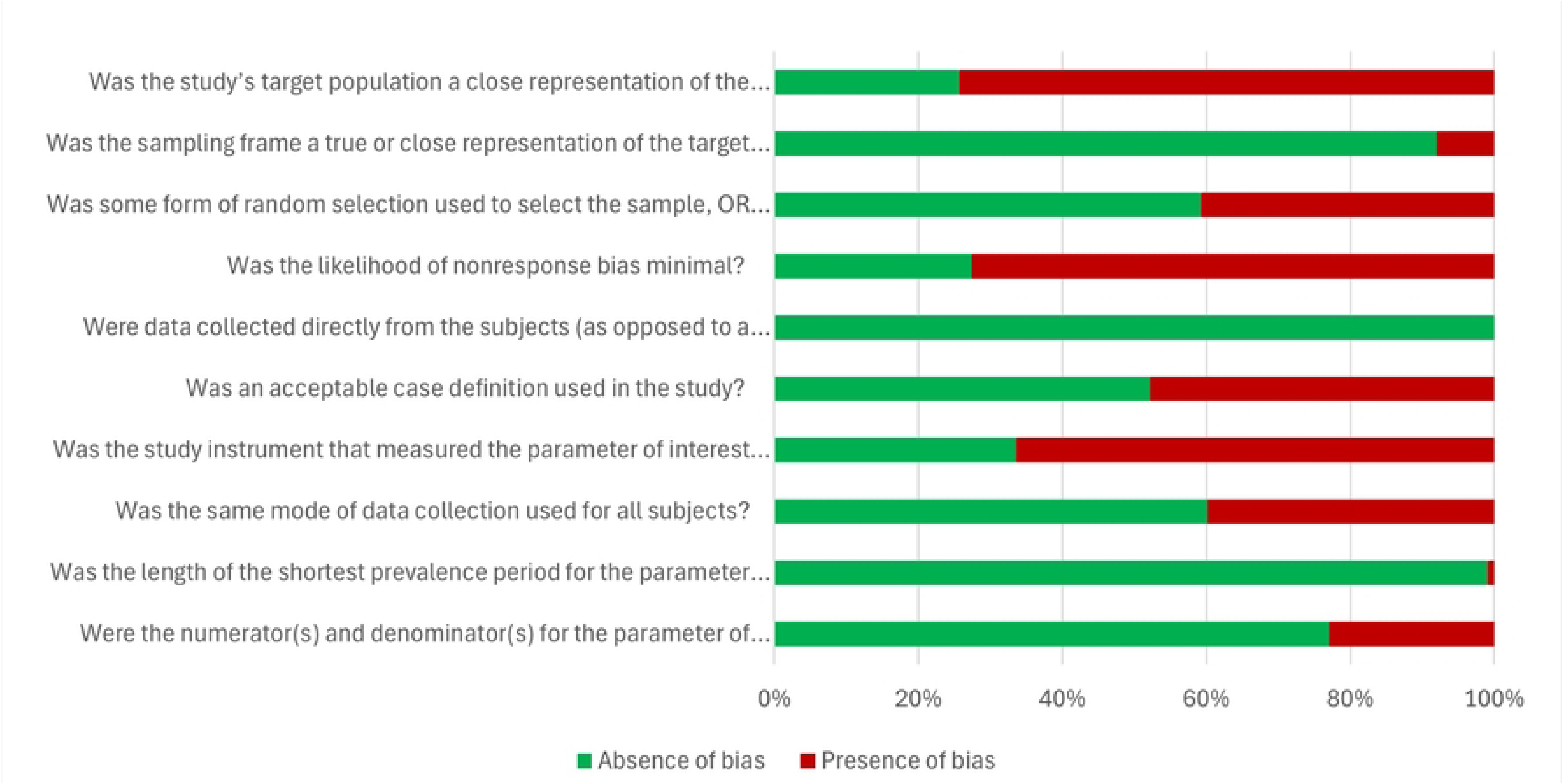
Graphical representation of the quality of the included studies.

Approximately half of the studies (52/113) clearly defined their KAP measurements (item 6), while only 38 studies used validated questionnaires (item 7). The same data collection method was used in 68 out of 113 studies (item 8). Notably, only one study (1/113) employed a recall period of more than four weeks to assess KAP among healthcare professionals (item 9). Furthermore, 26 out of 113 studies did not provide true values for both the numerator and denominator (item 10). All the included studies assessed KAP using self-reported questionnaires (item 5) and were uniformly scored as ‘0’ or at no risk of bias. However, self-reported data are inherently prone to bias in practice. Despite this limitation, no studies were excluded based on their quality.

### 3.5. Synthesis of evidence

The findings from the included studies are presented below, categorised by the outcomes of interest.

#### 3.5.1. Evidence relating to infection control and management (IPCM)

Infection prevention, control and management (IPCM) was summarised as KAP measure for hand-hygiene (HH) practice[58, 75, 88, 92, 93, 96, 97, 105, 125], prevention of infections such as blood-borne infections[117, 122] and hospital-acquired infections[101], attitude towards vaccine implementation[24, 28, 30, 32, 34, 40, 50, 55–57, 64, 74, 77, 104] and prevention of disease including measles[18], hepatitis A[28], coccidioidomycosis[29], and COVID-19.[49]

Two studies provided an overall knowledge score for IPCM, measured among Hungarian nurses (16.69 ± 2.504 out of 23 questions)[120] and among junior doctors from Portugal (9.35±1.65 out of 12 questions)[107]. A study conducted in the USA reported that among 265 participating dental healthcare professionals, two-thirds were able to correctly answer 6 to 9 out of 13 questions, reflecting a perceived knowledge score with a mean of 58% on CDC infection control guidelines.[42] In Australia, 94% of nurses correctly identified the main reason for cleaning as “to reduce the risk for infection”.[53] Two studies measuring knowledge among dental care professionals about blood-borne infections revealed limited perceived knowledge of infection control; only 37.3% reported being well informed on infection control guidelines.[117, 122] Galanis et al[101] measured knowledge and practice among 107 Greek HCPs. The mean overall knowledge score for hospital infections was 59.4, indicating moderate knowledge.[101] While measuring knowledge about vaccination and disease, Greek educators and HCPs showed varied knowledge about Human Papilloma Virus (HPV) vaccination with only 26.4% of 750 respondents having excellent knowledge.[104] In contrast, HCPs from a German paediatric cancer unit poses moderate knowledge as 63 (59.4% of 106 participants) of them were able to answer all knowledge questions correctly.[77]

Attitude towards IPCM, as reported by motivation for HH compliance, found that infection risk motivated HH compliance among the Greek medical students (86.4%) and nursing students (90%).[105] Among Canadian family medicine residents, 50% (of 150 respondents) agreed with the statement “hand hygiene compliance among healthcare providers is a serious problem”, and 45% agreed that better HH prevents Hospital-acquired infections.[97] When measuring attitude towards vaccination and diseases, Trucchi et al[64] also reported a higher attitude score of 5 (25–75, p=4–5, among 1410 subjects) related to disease prevention statements, with Italian medical doctors showing more positive attitude compared to other HCPs.

Practice behaviour among HCPs varies among the studies. Bounou et al[105] found that medical students had better HH compliance (69.9% Vs 59.7%, p <0.001) and proper HH technique (47.1 Vs 32.7%, p <0.001) compared to nursing students. Hammerschmidt et al. [75] reported that 47% of nurses have perceived HH compliance. Two studies highlighted the practice of recommending COVID-19 vaccination as an important measure for IPCM.[34, 55] Papini et al. reported that only 0.88% (18 out of 2052) of all respondents did not recommend the COVID-19 vaccine. Among those, healthcare assistants were more likely to refrain from recommending the vaccine compared to other groups. Conversely, Woodward et al[34] found suburban providers less likely to recommend the vaccine to patients (OR=0.43, 95% CI: 0.22–0.87) and family (OR=0.45, 95% CI: 0.22–0.92) compared to urban providers.

#### 3.5.2. Evidence relating to antimicrobial stewardship (AMS)

Twenty-four [19, 20, 22, 23, 33, 35, 37, 38, 41, 43, 47, 54, 69, 70, 76, 87, 99, 106, 109, 113, 115, 121, 128, 130]studies measured knowledge, awareness or perception about AMS. The familiarity with AMS, reported in the included studies, ranged from 44.5% to 72.9% among HCPs.[47, 130] Another study conducted among Belgian HCPs found that about 40.8% of HCPs knew about AMS, but only 21.5% understood its roles.[121] Familiarity with AMS among HCPs can vary based on setting, profession type and experience. For instance, 23% of prescribers from small community hospitals (SCH) and 28% from large community hospitals (LCH) reported to be aware of AMS.[35] Self-rated AMS awareness, as measured among Australian nurses varied, (33% of participants with no experience Vs 65% of enrolled nurses, p<0.00).[47] Similarly, Inanco et al[70] showed that awareness of AMS improves with education, increasing from 21% in1^st^-year students to 79% in 3^rd^ and 4^th^-year students. Positive AMS awareness was noted among GPs (median 4.0, IQR 1), with 61.7% understanding its role in reducing inappropriate antimicrobial use.[54] Whereas the mean AMS knowledge score among pharmacy students from multiple countries was 71.85±6.06 (out of 91).[69]

None of the studies assessed attitude using a standardised set of questions. The majority evaluated attitude using a Likert scale or agree/disagreement statement. Only one study provided an attitude score among pharmacy students, reported as 36.50 ±5.29 (maximum available score 45).[69] About 67.9% of pharmacists, as reported in the study by Lee et al[44] felt AMS is important in the community and would participate in AMS activities if the opportunity was provided (88.5%). Zetts et al[22] reported that 72% of primary care physicians believe antibiotic stewardship programmes are necessary to combat antibiotic resistance, with the highest agreement among paediatricians (77%) and the lowest among internal medicine physicians (68%) (P =0.024). Almost all prescribers from SCH and LHC agreed that inappropriate use of antimicrobials causes antibiotic resistance (∼98%), antimicrobial use can harm patients (∼96%), antimicrobial resistance is a significant problem nationally (94%) and antimicrobials are overused nationally (>95%).[35]

We summarised AMS practice as participation in AMS programmes. A study among French nursing homes (NH) reported that less than half of the participating NHs have core AMS elements in place and the most frequently practised ASP activities were antibiotic consumption monitoring (65%), antibiotic plan documentation (56%) and antibiotic prescription guide distribution (54%).[131] When compared among pharmacists from SCH and LCH, 76% from SCH and 65% from LCH were engaged in AMS practice.[35] Comparing self-Assessment (SAT)data from 2014-2016 and 2019, it appeared that the clinicians showed strong adherence to positive AMS practices, including using antibiotic guidelines for treatment decisions (98%), discussing antibiotic prescribing within their practices (5% increased), utilising patient-facing educational resources (23% increases), and conducting antibiotic audits in the past two years (53% increased).[72]

#### 3.5.3. Evidence relating to antimicrobial resistance (AMR) and antibiotic use

Knowledge about AMR was measured in a varied way across the studies. According to the ECDC 2019[66, 123] survey 96% of 18,365 HCWs across 30 EU countries believed they had sufficient knowledge and 80% felt knowledgeable about antibiotic use in their practice, yet only 58% of them answered all knowledge questions correctly. When measured among 143 Italian HCPs, it was found that 68.5% perceived having sufficient knowledge about the use of antimicrobials but 90% to 95% of respondents correctly answering 5/7 key knowledge questions.[58] Padigos et al[115] found that most registered nurses in New Zealand perceived their AMR knowledge as average. Another study, conducted in a US nursing home, reported that 70% of nurses were aware of the dangers of taking antibiotics.[39] Studies focusing on students found that, senior UK veterinary students had good knowledge about AMR when compared to early year (Mean, SD= 6.67,1.05) and later year students (7.25 0.84), [68] 98% (180) of UK pharmacy students had heard of AMR[70] and 93% (254/274) of senior Australian pharmacy students were familiar with AMR, compared to 80% (200/251) of junior students (p<0.0001).[45] Among prescribers, participant from three Canadian teaching hospitals showed limited knowledge of antibiotic therapy for treatment of infectious disease with only 14 (3%) of them correctly answered all seven knowledge questions.[100] Similarly, a knowledge gap was also observed among US antimicrobial prescribers (n=402), where just 5 of 303 respondents (2%) answered all knowledge questions correctly.[21]

Attitude for AMR and antibiotic use was measured as motivation, belief and opportunity. Among HCPs, 51.6%[23] of nurses and about 90%[71, 82] of dentists believed AMR is a concern. 97.1%(of 415) UK pharmacists believed that misuse of antibiotics can lead to ineffective treatment.[60] Australian veterinary students showed strong awareness of AMR, with 84.4% agreeing on the need for immediate action.[52] In contrast, UK veterinary students perceived their responsibility in addressing and preventing AMR as lower.[68] Regarding the attitude towards antibiotic use, US nurses often reported evidence-based attitudes towards antibiotics.[39] However, incomplete knowledge regarding the ‘effectiveness of antibiotic prescribing’ was reported in one study.[116] Corrente et al[62] reported data on attitudes towards antibiotic consumption among 119 veterinary students from Italy. Seventy-four respondents (70%) believed it is necessary to not miss doses, 87 (82%) comply with doses when feeling better and 43 (40.6%) preferred to choose older generation antibiotics over a newer generation.

Practice related to AMR was measured among 26 [15, 22, 26, 33, 37, 40, 41, 45, 58, 59, 61, 63, 66, 68, 73, 80, 81, 84, 89–91, 94, 98, 103, 121, 132]studies and 30 [20, 21, 25, 31, 33, 37, 45, 46, 51, 60–62, 66–68, 78, 82, 84, 87, 91, 94, 102, 103, 106, 109, 111, 113, 121, 123, 124, 126, 133]reported practices related to antimicrobial use. A large study among European HCPs showed that only 17% (2,430/14,294) provided resources like leaflets, and 55% (7,861/14,294) offered advice on prudent antibiotic use or infection management.[123] At the individual profession level, 89% of hospital physicians(n=170) and 87% of veterinarians(n=60) identified ambulatory antibiotic prescriptions as the primary starting point for reducing AMR.[80] When compared to medical and dental providers, dentists were more often influenced by concerns about patient compliance when prescribing prophylactic antibiotics (13% versus 1%, P = .02).[27] In a study by Schneider et al[79] approximately 80% of all prescriber groups stated that antibiotic guidelines were an important information source. However, application varied across HCPs: 65% of hospital physicians, 53% of veterinarians, and 39% of GPs reported frequent routine use of antibiotic guidelines.[80] For antibiotic deprescribing, only 2.5% (95% CI, 90.8–94%) of Italian GPs reported using the antibiotic deprescribing strategy at least once.[112] While GPs are involved in deprescribing, a study among Italian dentists found they reported over-prescribing antibiotics with 70.9% (of 563) respondents reporting having inappropriately prescribed antibiotic prophylaxis. [60] Reasons for antibiotic prescription varied among dentists depending on their location. A study among 92 French dentists (n=92) reported that abscesses with systemic symptoms (89%), cellulitis (81.5%), acute sinusitis (62%) were the top three reasons for prescribing, with amoxicillin being the preferred antibiotic.[111] In contrast, from a study among 437 US dentists prophylaxis before invasive procedures for patients with “high-risk conditions” (84%) emerged as the main reason for antibiotic prescription.[31] A graphical presentation of evidence is provided in figure 4.

**Figure 4:**
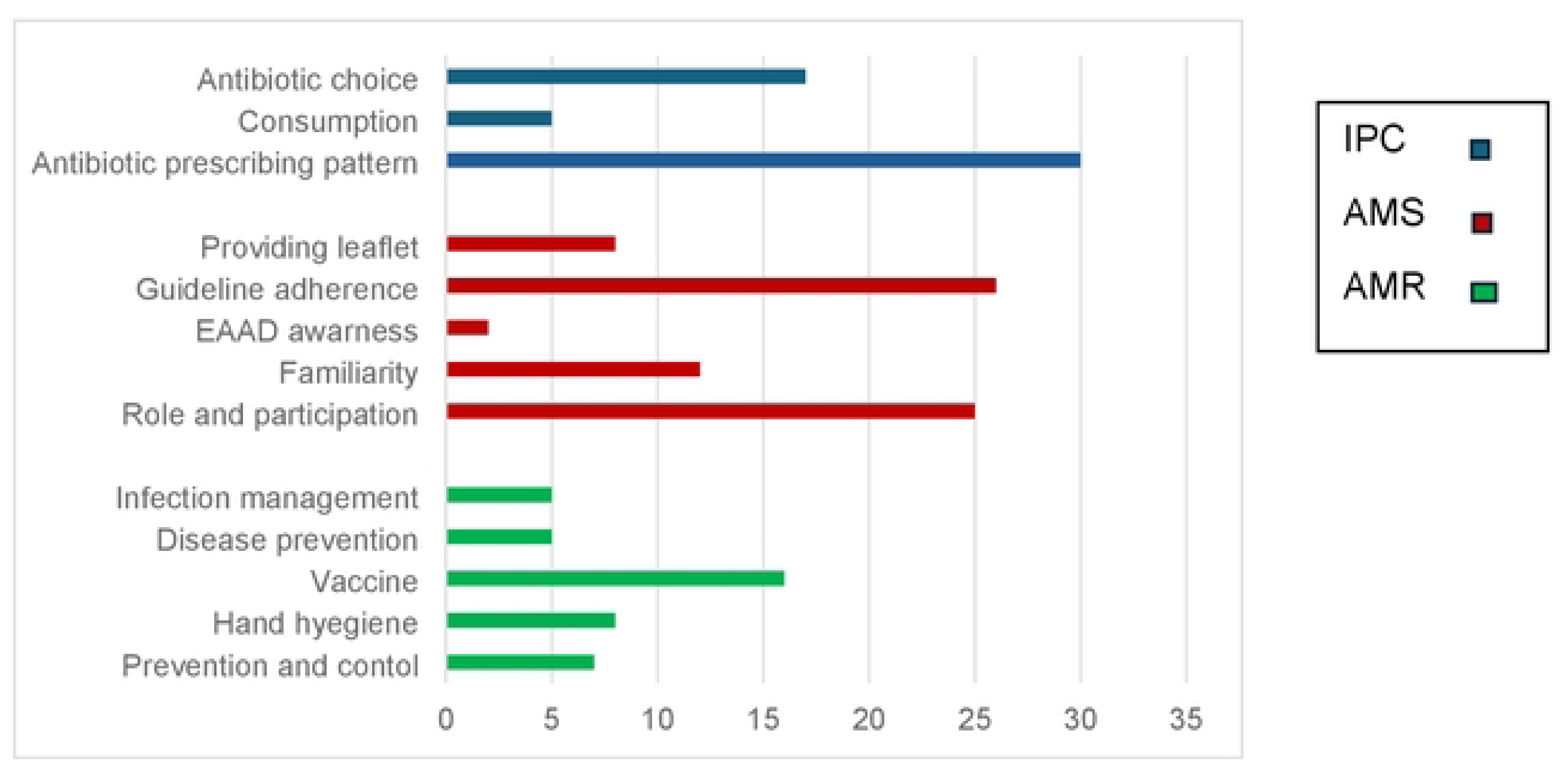
Graphical representation of evidence.

## 4. Discussion

This systematic review presents an in-depth synthesis of self-reported data concerning knowledge, attitudes, and practices of health and social care workers and students engaged in infection prevention and control, antimicrobial use, stewardship, and resistance. We have explored these three domains—knowledge, attitudes, and practices—to present a comprehensive overview. Additionally, the findings reveal significant variability across different professions, settings, and levels of experience.

The knowledge analysis indicates that while many HCPs possess robust knowledge of IPCM guidelines and hand hygiene practices, gaps persist, particularly among dental professionals and those less exposed to vaccination protocols. AMS knowledge varies widely across professions, settings, and experience levels. Pharmacists generally have a stronger grasp of AMS principles and could play a key role in promoting antimicrobial stewardship, but significant gaps remain among doctors, nurses, and pharmacy students. Despite recognising AMR as a major global health issue, HCPs often overestimate their knowledge, especially regarding antibiotic use and stewardship. Awareness of initiatives like the European Antibiotic Awareness Day (EAAD)[134] and World Antimicrobial Awareness Week (WAAW)[135] remains low, underscoring the need for better outreach. Variability in understanding AMR’s contributing factors and preventive strategies highlights the necessity for targeted, comprehensive training. Significant gaps also exist in adherence to antibiotic use guidelines, especially among nursing home nurses, physicians, and prescribers.[136] Personal factors, such as familial ties to healthcare, may further influence knowledge and attitudes toward antibiotic use.

A limited number of studies were focused on attitude. HCPs, especially dental practitioners, demonstrate a proactive commitment to infection control, adhering to CDC guidelines.

However, evidence indicates low compliance with some infection control measures. Medical and nursing students exhibit strong motivation to follow hand hygiene protocols, recognising the risks of healthcare-associated infections. Regarding AMS, most HCPs, including prescribers, acknowledge its importance in combating AMR. However, despite confidence in their education, there is a high demand for further training, particularly in infectious disease management. Many physicians, however, focus on other public health concerns, and nurses and young physicians often express uncertainty about their roles, indicating the need for clearer guidance and education.[137]

In addition to the reported knowledge and attitude, this systematic review reveals significant variability in healthcare professionals’ adherence to infection control, AMS, and antibiotic prescribing practices. In the realm of HH and vaccination, compliance rates varied considerably across professions and regions. Medical students and other healthcare providers, particularly in Italy, showed higher adherence to HH practices, another component of IPCM. This suggests that professional training and exposure to infection control programmes may influence adherence to IPCM measures.[138] AMS practices among HCPs also revealed variability. While pharmacists in smaller community hospitals and some French nursing homes demonstrated relatively high engagement with AMS protocols, gaps remain in the broader implementation of stewardship programmes, particularly in delayed antibiotic prescribing and consulting with infectious disease specialists. Antibiotic prescribing practices, as assessed in the studies, present clear challenges related to AMR. While many HCPs recognise overprescription issues, adherence to guidelines is inconsistent, especially among less experienced physicians and in dental care. The dispensing of antibiotics without prescriptions, as observed among Italian pharmacists, underscores regulatory shortcomings. Misuse of prophylactic antibiotics by dental professionals and frequent prescribing for non-recommended reasons in the U.S. and Europe point to persistent knowledge gaps and patient compliance challenges. Patient expectations also influence prescribing behaviours, with Swedish professionals noting significant pressure from patients and some German general practitioners prescribing antibiotics without strong indications, especially before weekends. Additionally, veterinary students often favour older-generation antibiotics, which may not align with best practices, underscoring the need for enhanced education and training. Despite AMS progress, substantial opportunities remain for improvement through targeted education, stricter guidelines, and regulatory enforcement. Early stewardship training for young doctors could help mitigate antibiotic overuse.[139]

Despite our rigorous approach to summarise KAP among the wider health and social care workforce, we acknowledge the possibility that some relevant studies may have been missed. The lack of standardised and validated survey[140] instruments presented challenges in achieving consistent data analysis. Additionally, since the review included self-reported data, there is potential for bias, which we attempted to address through critical appraisal, although no study was excluded based on quality. Evidence was limited for certain healthcare professional subgroups, such as allied health professionals, complicating comparisons. Furthermore, by concentrating on settings similar to the UK, we may have overlooked contexts in lower- and middle-income countries (LMICs), policies on antimicrobial use are less strictly enforced and less widely followed. Nonetheless, our findings align with existing literature emphasising the need to improve HCWs’ compliance with IPCM measures.[141] Additionally, strengthening medical students’ understanding of antimicrobial attitudes and appropriate use is crucial for combating AMR, reinforcing the reliability and relevance of our review.[142]

In conclusion, this systematic review underscores the significant variability in HCPs’ knowledge, attitudes, and practices related to infection prevention, AMS, and AMR. While many HCPs demonstrate a proactive approach to infection control and recognize the importance of AMS, persistent knowledge gaps and barriers hinder optimal practices. To effectively combat AMR and improve infection management, targeted educational initiatives, enhanced guideline enforcement, and stronger stewardship programs are crucial. By fostering collaboration and continuous professional development across all healthcare sectors, we can drive progress in responsible antimicrobial use and patient safety.

## Supporting information

The search strategy and terms used are presented in the Appendix 1

The methodological quality of the included studies is summarised in Figure 3 and Appendix 2.

## Data availability

The datasets used and/or analysed during the current study are available from the corresponding author on reasonable request.

## Author contribution

The study was commissioned by DAO. AAy, DAO, EG, IG and PA conceived and designed the study. SJ developed and ran the searches. AA, EA, VA and IG screened the title and abstracts. EA, VA and IG screened the full texts. AA, EA, VA, IG and CW extracted data and critically appraised the included studies. AAy, IG and PA developed the data synthesis plan. IG drafted the manuscript. All authors contributed to the final manuscript.

