## Supplementary material for "Self-reported knowledge attitude and practice of healthcare professionals in the management of infection and antimicrobial stewardship: a systematic review": The search strategy and terms used are presented in the Appendix 1

Abi AMR V6 Stakeholder edits

Searches run: 24 Jan 2024

|  | Nos. References retrieved | Deduplicated rreferences |
| --- | --- | --- |
| Medline | 1466 | 1466 |
| Embase | 6329 | 5595 |
| CINAHL | 3643 | 2925 |
| Web of Science | 1972 | 1004 |
| TOTAL | 13,410 | 10,990 |

Ovid MEDLINE(R) ALL <1946 to January 23, 2024>

1	Drug Resistance, Microbial/	61621

2	exp Drug Resistance, Bacterial/	103770

3	Antimicrobial Stewardship/	3504

4	antimicrobial resistan*.mp.	42262

5	(antimicrobial adj3 ("use" or stewardship or resistan*)).mp. [mp=title, book title, abstract, original title, name of substance word, subject heading word, floating sub-heading word, keyword heading word, organism supplementary concept word, protocol supplementary concept word, rare disease supplementary concept word, unique identifier, synonyms, population supplementary concept word, anatomy supplementary concept word]	57771

6	Infection Control/ or infection prevention control.mp.	29067

7	exp Anti-Bacterial Agents/tu [Therapeutic Use]	285870

8	exp Antifungal Agents/tu [Therapeutic Use]	55873

9	exp Antiviral Agents/tu [Therapeutic Use]	138482

10	exp Antiparasitic Agents/tu [Therapeutic Use]	89421

11	1 or 2 or 3 or 4 or 5 or 6 or 7 or 8 or 9 or 10	686376

12	exp "Attitude of Health Personnel"/	170731

13	Health Knowledge, Attitudes, Practice/	127734

14	Behavior/ or Habits/	36346

15	(attitude* or knowledge or practice* or opinion* or belief* or perspective* or behaviour* or behavior* or "com-b" or values or values or understanding or awareness or culture).mp. [mp=title, book title, abstract, original title, name of substance word, subject heading word, floating sub-heading word, keyword heading word, organism supplementary concept word, protocol supplementary concept word, rare disease supplementary concept word, unique identifier, synonyms, population supplementary concept word, anatomy supplementary concept word]	7645072

16	12 or 13 or 14 or 15	7667761

17	11 and 16	136460

18	exp Health Personnel/	625213

19	exp Students/	172645

20	(careworker* or care worker* or care-worker* or healthcare worker* or health care worker* or health-care worker* or social adult care or heathcare support worker* or health care support worker* or health-care support worker*).mp. [mp=title, book title, abstract, original title, name of substance word, subject heading word, floating sub-heading word, keyword heading word, organism supplementary concept word, protocol supplementary concept word, rare disease supplementary concept word, unique identifier, synonyms, population supplementary concept word, anatomy supplementary concept word]	44203

21	18 or 19 or 20	802141

22	11 and 16 and 21	7083

23	exp Africa, Central/ or exp Africa, Southern/ or exp Africa, Northern/ or exp "Africa South of the Sahara"/ or exp Africa, Western/ or exp Africa, Eastern/ or exp Africa/ or exp South Africa/	332080

24	exp Latin America/ or exp South America/	211662

25	exp Asia, Northern/ or exp Asia, Eastern/ or exp Asia, Southern/ or exp Asia/ or exp Asia, Southeastern/ or exp Asia, Central/ or exp Asia, Western/	1055090

26	23 or 24 or 25	1570628

27	22 not 26	5153

28	limit 27 to yr="2016 - 2024"	2310

29	exp animals/ not humans.sh.	5189902

30	28 not 29	2274

31	limit 30 to english language	2207

32	exp "Surveys and Questionnaires"/	1228636

33	(questionnaire* or survey*).mp. [mp=title, book title, abstract, original title, name of substance word, subject heading word, floating sub-heading word, keyword heading word, organism supplementary concept word, protocol supplementary concept word, rare disease supplementary concept word, unique identifier, synonyms, population supplementary concept word, anatomy supplementary concept word]	1619033

34	(assess* or tool or tools or instrument* or measur* or scale or index).mp. [mp=title, book title, abstract, original title, name of substance word, subject heading word, floating sub-heading word, keyword heading word, organism supplementary concept word, protocol supplementary concept word, rare disease supplementary concept word, unique identifier, synonyms, population supplementary concept word, anatomy supplementary concept word]	9683886

35	32 or 33 or 34	10452063

36	31 and 35	1466

Embase <1980 to 2024 Week 03>

1	antimicrobial resistance.mp. or exp antibiotic resistance/	222042

2	antimicrobial stewardship.mp. or exp antimicrobial stewardship/ or exp antibiotic resistance/	221072

3	microbial drug resistance.mp.	349

4	(antimicrobial adj3 ("use" or stewardship or resistan*)).mp. [mp=title, abstract, heading word, drug trade name, original title, device manufacturer, drug manufacturer, device trade name, keyword heading word, floating subheading word, candidate term word]	72493

5	exp infection control/	123529

6	exp infection prevention/ or infection prevention control.mp.	80115

7	anti-bacterial agent*.mp. or antiinfective agent/dt, th [Drug Therapy, Therapy]	59181

8	exp antifungal agent/dt, th [Drug Therapy, Therapy]	137794

9	exp antivirus agent/dt [Drug Therapy]	432935

10	exp antiparasitic agent/dt [Drug Therapy]	169632

11	1 or 2 or 3 or 4 or 5 or 6 or 7 or 8 or 9 or 10	1060918

12	health personnel.mp. or exp health care personnel/	2027302

13	physicians.mp. or exp physician/	1227845

14	nurses.mp. or exp nurse/	376652

15	exp graduate nursing student/ or student/ or exp occupational therapy student/ or exp physician assistant student/ or exp baccalaureate nursing student/ or exp paramedical student/ or exp audiology student/ or exp allied health student/ or exp health student/ or exp medical student/ or exp pharmacy student/ or exp physical therapy student/ or exp public health student/ or exp graduate student/ or exp nursing student/ or exp dental student/	263658

16	exp administrative personnel/	33216

17	exp hospital administrator/	9642

18	exp hospital personnel/	148679

19	exp pharmacist/	99607

20	(careworker* or care worker* or care-worker* or healthcare worker* or health care worker* or health-care worker* or social adult care or healthcare support worker* or health care support worker* or health-care support worker*).mp.	53497

21	exp caregiver/	118024

22	12 or 13 or 14 or 15 or 16 or 17 or 18 or 19 or 20 or 21	2536318

23	exp health personnel attitude/ or exp physician attitude/ or attitude/ or exp attitude to health/	383618

24	health knowledge.mp.	6164

25	exp behavior/ or exp habit/	4942578

26	(attitude* or knowledge or practice* or opinion* or belief* or perspective* or behaviour* or behavior* or "com-b" or values or value or understanding or awareness or culture or "human factor*").mp. [mp=title, abstract, heading word, drug trade name, original title, device manufacturer, drug manufacturer, device trade name, keyword heading word, floating subheading word, candidate term word]	11234251

27	23 or 24 or 25 or 26	13386440

28	11 and 22 and 27	48760

29	limit 28 to yr="2016 -Current"	25384

30	(exp animal/ or exp invertebrate/ or nonhuman/ or animal experiment/ or animal tissue/ or animal model/ or exp plant/ or exp fungus/) not (exp human/ or human tissue/)	7338457

31	29 not 30	25019

32	exp "Africa south of the Sahara"/ or exp South Africa/ or exp Africa/ or exp North Africa/ or exp Central Africa/	409974

33	exp central Asia/ or exp western Asia/ or exp Asia/ or exp Southeast Asia/ or exp South Asia/ or exp northern Asia/	1382497

34	exp Central America/ or exp "South and Central America"/ or exp South America/	293809

35	32 or 33 or 34	2003173

36	31 not 35	18512

37	limit 36 to (article or article in press)	10779

38	limit 37 to english language	10391

39	exp health survey/ or exp health care survey/	300289

40	exp questionnaire/	949016

41	(Survey* or questionnaire*).mp. [mp=title, abstract, heading word, drug trade name, original title, device manufacturer, drug manufacturer, device trade name, keyword heading word, floating subheading word, candidate term word]	2718791

42	assess*.mp.	6938601

43	instrument*.mp.	680389

44	(tool or tools or scale or index).mp. [mp=title, abstract, heading word, drug trade name, original title, device manufacturer, drug manufacturer, device trade name, keyword heading word, floating subheading word, candidate term word]	4243413

45	39 or 40 or 41 or 42 or 43 or 44	11299804

46	38 and 45	6329

CINAHL

| **#** | **Query** | **Results** |
| --- | --- | --- |
| S1 | (MH "Drug Resistance, Microbial+") OR "antimicrobial resistance" OR (MH "Antiviral Agents+") OR (MH "Antifungal Agents") OR (MH "Antiparasitic Agents+") OR (MH "Antibiotics, Antifungal+") | Display |
| S2 | (MH "Antimicrobial Stewardship") OR "antimicrobial stewardship" | Display |
| S3 | (MH "Antiinfective Agents+") OR "antiinfective agents" | Display |
| S4 | (antimicrobial N3 (use or stewardship or resistan*)). | Display |
| S5 | (MH "Infection Control+") OR "infection control" | Display |
| S6 | "infection prevention control" | Display |
| S7 | S1 OR S2 OR S3 OR S4 OR S5 OR S6 | Display |
| S8 | (MH "Attitude+") OR (MH "Attitude of Health Personnel+") OR (MH "Employee Attitudes") or (MH "Habits+") | 634,261 |
| S9 | (MH "Health Knowledge") OR (MH "Professional Knowledge+") OR (MH "Nursing Knowledge") OR (MH "Student Knowledge") | Display |
| S10 | attitude* or knowledge or practice* or opinion* or belief* or perspective* or behaviour* or behavior* or com-b | Display |
| S11 | S8 OR S9 OR S10 | 1,962,204 |
| S12 | (MH "Health Personnel+") OR "health personnel" OR (MH "Administrative Personnel+") | Display |
| S13 | (MH "Students+") OR "students" | Display |
| S14 | (MH "Nurses+") OR "nurses" | Display |
| S15 | S12 OR S13 OR S14 | Display |
| S16 | S7 AND S11 AND S15 | 13,298 |
| S17 | s16 NOT (((MH "Animals+") OR (MH "Animal Studies") OR (TI "animal model*")) NOT (MH "human")) | 13,272 |
| S18 | (MH "Africa+") OR (MH "Africa South of the Sahara+") OR (MH "Africa, Western+") OR (MH "Africa, Eastern+") OR (MH "Africa, Southern+") OR (MH "Africa, Northern+") OR (MH "Africa, Central+") OR (MH "South Africa") | Display |
| S19 | (MH "Asia") OR (MH "Asia, Southern") OR (MH "Asia, Western") OR (MH "Asia, Southeastern") OR (MH "Asia, Central") | Display |
| S20 | (MH "South America+") OR (MH "Latin America") OR (MH "Central America+") | Display |
| S21 | S18 OR S19 OR S20 | Display |
| S22 | s17 not s21 | 12,017 |
| S23 | s17 not s21 | 6,109 |
| S24 | (MH "Questionnaires+") | Display |
| S25 | (MH "Surveys+") | Display |
| S26 | questionnaire* or survey* or scale or instrument* or index | Display |
| S27 | tool or tools or measur* or assess* | Display |
| S28 | S24 OR S25 OR S26 OR S27 | Display |
| S29 | S23 AND S28 | 3,643 |

Web of Science Search Strategy

1. "antimicrobial resistan*" or "anti-microbial resistan*" or "antimicrobial stewardship" or "microbial drug resistan*" or "bacterial drug resistan*" or "infection control" or "infection prevention control" or "anti-bacterial agent*" or "antibacterial agent*" or "anti-viral agent*" or "antiviral agent*" or "anti-fungal agent*" or "antifungal agent*" or "anti-parasitic agent*" or "antiparasitic agent*" or ((antimicrobial or anti-microbial) NEAR/2 ("use" or stewardship or resistan*)) (Topic)
2. attitude* or knowledge or practice* or opinion* or belief* or perspective* or value or values or understanding or awareness or culture or behaviour* or behavior* or com-b (Topic)
3. "health personnel" or "healthcare worker*" or "health care worker*" or doctor* or physician* or nurs* or therapist*or student* or pharmacist or "community health worker" or careworker* or "care worker*" or "care-worker*" or "healthcare worker*" or "health care worker*" or "health-care worker*" or "social adult care" or "healthcare support worker*" or "health care support worker*" or "health-care support worker*" or caregiver* or "care-giver*" (Topic)
4. questionnaire* or survey* or tool or tools or instrument* or assess* or measur* or scale or index (Topic)
5. 1 AND 2 AND 3 AND 4
6. “Low and middle income countries" OR "developing countries" OR BRICS OR BRIC OR Africa OR "north africa" OR "northern Africa" OR "Africa South of the Sahara" OR "Central Africa" OR "east Africa" OR "Eastern Africa" OR "Southern Africa" OR "Western Asia" OR "central Asia" OR "southeastern Asia" OR "Caribbean" OR "West Indies" OR "South America" OR "Latin America" OR "Central America" OR Afghanistan OR Albania OR Algeria OR "American Samoa" OR Angola OR "Antigua and Barbuda" OR Argentina OR Armenia OR Azerbaijan OR Bahrain OR Bangladesh OR Barbados OR Benin OR Belarus OR Belize OR Bhutan OR Bolivia OR Bosnia-Herzegovina OR Botswana OR Brazil OR Bulgaria OR "Burkina Faso" OR Burundi OR Cambodia OR Cameroon OR "Cape Verde" OR "Central African Republic" OR Chad OR Chile OR China OR Colombia OR Comoros OR Congo OR "Costa Rica" OR "Cote d'Ivoire" OR Croatia OR Cuba OR Cyprus OR Czechoslovakia OR "Czech Republic" OR Slovakia OR Djibouti OR "Democratic Republic of the Congo" OR Dominica OR "Dominican Republic" OR "East Timor" OR Ecuador OR Egypt OR "El Salvador" OR Eritrea OR Estonia OR Ethiopia OR Fiji OR Gabon OR Gambia OR "Georgia" OR Ghana OR Greece OR Grenada OR Guatemala OR Guinea OR "Guinea-Bissau" OR Guam OR Guyana OR Haiti OR Honduras OR Hungary OR India OR Indonesia OR Iran OR Iraq OR Jamaica OR Jordan OR Kazakhstan OR Kenya OR Korea OR Kosovo OR Kyrgyzstan OR Laos OR Latvia OR Lebanon OR Lesotho OR Liberia OR Libya OR Lithuania OR Macedonia OR Madagascar OR Malaysia OR Malawi OR Mali OR Malta OR Mauritania OR Mauritius OR Mexico OR Micronesia OR "Middle East" OR Moldova OR Mongolia OR Montenegro OR Morocco OR Mozambique OR Myanmar OR Namibia OR Nepal OR "Netherlands Antilles" OR "New Caledonia" OR Nicaragua OR Niger OR Nigeria OR Oman OR Pakistan OR Palau OR Panama OR "Papua New Guinea" OR Paraguay OR Peru OR Philippines OR Poland OR Portugal OR "Puerto Rico" OR Romania OR Russia OR Rwanda OR "Saint Kitts and Nevis" OR "Saint Lucia" OR "Saint Vincent and the Grenadines" OR Samoa OR "Saudi Arabia" OR Senegal OR Serbia OR Montenegro OR Seychelles OR "Sierra Leone" OR Slovenia OR "Sri Lanka" OR Somalia OR "South Africa" OR Sudan OR Suriname OR Swaziland OR Syria OR Tajikistan OR Tanzania OR Thailand OR Togo OR Tonga OR "Trinidad and Tobago" OR Tunisia OR Turkey OR Turkmenistan OR Uganda OR Ukraine OR Uruguay OR USSR OR Uzbekistan OR Vanuatu OR Venezuela OR Vietnam OR Yemen OR Yugoslavia OR Zambia OR Zimbabwe) (Topic)
7. 5 NOT 6
8. Limited to 2016-2024 and English only	**1972**
